# Baseline intratumoral TCRβ and TCRγ repertoires predict response to immune checkpoint inhibitors in advanced renal cell carcinoma

**DOI:** 10.64898/2026.09.01.26361892

**Authors:** Manuel Pino-González, Martín Lázaro-Quintela, Guillermo de Velasco, Marta Dueñas, Irene Alonso-Álvarez, Alejandro Francisco-Fernández, Pilar San Fraile, José Antonio Ortiz, María Gallardo-Gómez, Laura Carracedo-Videira, Sandra Gallach, Eloisa Jantus-Lewintre, Mónica Martínez-Fernández

**Affiliations:** Translational Oncology Group, Galicia Sur Health Research Institute (IIS Galicia Sur), SERGAS-UVIGO, Hospital Álvaro Cunqueiro, Carretera Clara Campoamor, 341, 36213 (Vigo), Spain; Department of Medical Oncology, Biomedical Research Institute I+12, Hospital Universitario “12 de Octubre”, 28041 (Madrid), Spain; Molecular and Traslational Oncology Division, Biomedical Innovation Unit, CIEMAT, 28040 (Madrid), Spain. Cellular and Molecular Oncology and Genitourinary Tumor Group, Biomedical Research Institute Imas12, Hospital Universitario “12 de Octubre”, 28041 (Madrid), Spain. Centro de Investigación Biomédica en Red Cáncer, 28029 (Madrid), Spain; Department of Pathology, Hospital Álvaro Cunqueiro, Carretera Clara Campoamor, 341, 36213 (Vigo), Spain. Uropathology Research Group, Galicia Sur Health Research Institute (IIS Galicia Sur), SERGAS-UVIGO, Hospital Álvaro Cunqueiro, Carretera Clara Campoamor, 341, 36213 (Vigo), Spain; Molecular Oncology Laboratory, General University Hospital of Valencia Research Foundation, 46014 (Valencia), Spain; Instituto Interuniversitario de Investigaciión de Reconocimiento Molecular y Desarrollo Tecnológico (IDM), Universitat Politècnica de València - Universitat de València, Valencia, Spain y Unidad Mixta UPV-CIPF de Investigación en Mecanismos de Enfermedades y Nanomedicina, Universitat Politècnica de València - Centro de Investigación Príncipe Felipe, Department of Biotechnology. Universitat Politècnica de València, 46022 (Valencia), Spain

**Keywords:** Renal cell carcinoma, immune checkpoint inhibitor, predictive biomarker, immunotherapy response, T cell receptor, TCR repertoire

## Abstract

Immune checkpoint inhibitors (ICIs) have transformed the treatment landscape of advanced renal cell carcinoma (RCC), yet only a subset of patients derives durable clinical benefit and no robust predictive biomarkers have been implemented in routine clinical practice. As T-cell receptor (TCR)-mediated antigen recognition underlies effective antitumor immunity, characterization of the intratumoral TCR repertoire may provide clinically relevant information for patient stratification. Here, we evaluated the predictive value of the baseline intratumoral TCRβ and TCRγ repertoires in a real-world cohort of patients with advanced RCC treated predominantly with first-line immune checkpoint inhibitor-based regimens. Responders exhibited a significantly more even TCRβ repertoire together with increased TCRγ diversity and clonal richness, whereas non-responders showed greater TCRγ clonal dominance. In addition, the preferential usage of specific TRBV genes, including *TRBV5.7*, *TRBV7.1*, and *TRBV18*, was associated with clinical response and longer progression-free survival. In this exploratory cohort, integration of the most informative TCRβ and TCRγ variables into a combined random forest model yielded an area under the receiver operating characteristic curve of 0.90 in leave-one-out cross-validation, with 100% sensitivity and 81.8% specificity at the selected threshold. To our knowledge, this is the first study to simultaneously characterize baseline intratumoral TCRβ and TCRγ repertoires in patients with advanced RCC predominantly receiving first-line immune checkpoint blockade. These findings support baseline intratumoral TCR repertoire features as candidate tissue biomarkers for pretreatment patient stratification and provide a rationale for the prospective validation of TCR repertoire profiling as a predictor of immunotherapy benefit in RCC.

## Introduction

With a rising global incidence, renal cell carcinoma (RCC) is the 15th most commonly diagnosed cancer and the 16th leading cause of cancer-related death worldwide ^1^. RCC comprises several histological subtypes, with clear-cell RCC (ccRCC) accounting for 75-80% of cases, followed by papillary (pRCC) and chromophobe subtypes ^2,3^. ccRCC is predominantly driven by inactivation of the von Hippel-Lindau (VHL) tumor suppressor gene, which ultimately results in the activation of pro-angiogenic pathways. Although most RCC cases are diagnosed at stage I, approximately 11% present with stage IV disease, while an additional 10% of patients with initially localized RCC subsequently develop metastatic disease ^3^. Immune checkpoint inhibitors (ICIs) have transformed the treatment landscape of advanced RCC and are now established as the standard first-line treatment as monotherapy, dual immunotherapy, or in combination with VEGFR-TKIs, significantly improving patient survival ^2–7^. However, these clinical benefits are tempered by significant challenges, including immune-related adverse events, the limited response rate, and the absence of reliable predictive biomarkers ^2,3^.

The predictive value of several proposed biomarkers, including PD-1/PD-L1 expression ^8–10^, tumor mutational burden (TMB) ^4,11–13^, frameshift insertion/deletion (fsINDEL) ^4,11–13^, genome instability ^13^, T cell/myeloid infiltration ratio and B cell abundance ^6,11^ and expression of human endogenous retroviruses (HERVs) ^13–16^, remains equivocal in RCC and none has been proven to predict patient response in routine clinical practice.

Tumor-infiltrating cytotoxic T cells play a key role by recognizing cancer cell-specific antigens, known as neoantigens, and mediating the adaptive antitumor immune response through their T cell receptors (TCRs) ^17^. The efficacy of ICIs ^4–7^ supports RCC as an immunogenic tumor, although the mechanisms underlying response remaining incompletely understood. Large-scale studies showed RCC to be among the most highly immune-infiltrated solid tumor types ^18–20^. Interestingly, in contrast to other cancers ^21–23^, tumor-infiltrating cytotoxic T cell abundance is inversely correlated with clinical outcomes ^6,11,23,24^.

The TCR is composed of an α and a β chain in Tαβ cells (95%) or a γ and a δ chain in Tγδ cells (5%) ^17,25–28^. During T cell differentiation in the thymus, somatic V(D)J recombination generates highly diverse CDR3 regions, which determine antigen specificity and enable the generation of a vast repertoire of unique TCRs capable of recognizing a wide range of antigens ^17,25–28^.

Because studies across solid tumors have yielded contradictory results, such as lung ^29–33^, melanoma ^34^, cervical ^8^, breast ^35^, liver ^36^ and renal cancer ^13,37–42^ the potential of the TCR repertoire as a biomarker for immunotherapy response is unclear ^28^. Yet, despite the important role of TCRγδ cells in antitumor immune responses in solid cancers ^33,35,36,43–49^, most studies have focused on TCRαβ ^8,9,13,29–32,34,37–42^. These results highlight the need to determine whether associations between the TCRβ and TCRγ repertoires and clinical outcomes are cancer-specific. Accordingly, the main objective of this study was to evaluate the potential of the tumor-infiltrating TCRβ/γ repertoire at baseline as a biomarker for predicting the response to immunotherapy in a real-world cohort of advanced RCC patients treated with ICIs in the first-line setting.

## Materials and methods

### Study design and sample collection

This study included 20 RCC patients, treated with ICI-based regimens (as monotherapy, dual immunotherapy, or in combination with targeted therapy), 18 of whom received treatment in the first-line setting, across three centers: Hospital 12 de Octubre in Madrid (n = 13), Hospital Álvaro Cunqueiro in Vigo (n = 4), and Complexo Hospitalario Universitario in Pontevedra (n = 3). The study was approved by the Galician Clinical Research Ethics Committee (CEIm-G; 2022/429) and Hospital 12 de Octubre (18/055), was conducted in accordance with the Declaration of Helsinki, and all patients provided written informed consent. The infiltrating TCR repertoire was analysed for each patient. Patients were classified as responders (R: complete response, partial response, or stable disease) and non-responders (NR: progressive disease or not evaluable because of death) ^50^ following RECIST1.1 criteria, based on the computed tomography assessment at 3 months. The most relevant clinicopathological characteristics are summarized in *Table 1*. To examine the tumor-infiltrating TCR repertoire, formalin-fixed paraffin-embedded (FFPE) tumor biopsies were obtained from all patients prior to immunotherapy. DNA was chosen over RNA for analysis due to its superior stability in FFPE samples.

**Table 1:** Clinicopathological characteristics of patients.

| Clinicopathologic characteristics | Responders (N = 9) | Non-responders (N = 11) | Cohort (N = 20) |
| --- | --- | --- | --- |
| **Age (years)** |  |  |  |
| Median (range) | 64 (28-84) | 65 (36-76) | 64.5 (28-84) |
| **Sex** |  |  |  |
| Male | 7 | 6 | 13 |
| Female | 2 | 5 | 7 |
| **Histology** |  |  |  |
| ccRCC | 6 | 9 | 15 |
| pRCC | 3 | 1 | 4 |
| Unclassified | 0 | 1 | 1 |
| **Stage at diagnosis** |  |  |  |
| II | 1 | 0 | 1 |
| III | 3 | 2 | 5 |
| IV | 5 | 9 | 14 |
| **ICIs line** |  |  |  |
| 1st | 9 | 9 | 18 |
| 2nd | 0 | 2 | 2 |
| **Treatment** |  |  |  |
| Anti-PD-1 | 3 | 4 | 7 |
| Anti-PD-1 + Anti-CTLA-4 | 3 | 5 | 8 |
| Anti-PD-L1 + TT | 1 | 2 | 3 |
| Anti-PD-1 + TT | 1 | 0 | 1 |
| Anti-PD-1 + Anti-CTLA-4 + TT | 1 | 0 | 1 |
| **Response type** |  |  |  |
| CR | 1 | 0 | 1 |
| PR | 3 | 0 | 3 |
| SD | 5 | 0 | 5 |
| PD | 0 | 9 | 9 |
| NE | 0 | 2 | 2 |
| **PFS (days)** |  |  |  |
| Median (range) | 316 (197-2314) | 83 (0-1267) | 291.5 (0-2314) |
| **OS (days)** |  |  |  |
| Median (range) | 462 (197-2314) | 233 (7-1267) | 389 (7-2314) |

ccRCC: clear cell renal cell carcinoma, CR: complete response, N: number of patients, nccRCC: non-clear cell renal cell carcinoma, NE: non-evaluable. PD: progressive disease, PFS: progression-free survival, PR: partial response, pRCC: papillary renal cell carcinoma, SD: stable disease, TT: targeted therapy, OS: overall survival.

### DNA extraction

DNA from FFPE samples was extracted using the AllPrep® DNA/RNA FFPE Kit (QIAGEN) following the manufacturer’s instructions. DNA quantification was carried out using NanoDropTM 2000c Spectrophotometer (Thermo Fisher Scientific).

### Library preparation and TCR sequencing

Twenty libraries were generated from 250 ng of genomic DNA per sample using the Oncomine™ TCR Pan-Clonality Assay (Thermo Fisher Scientific). This targeted NGS assay amplifies and sequences the TCRβ/γ CDR3 region. Libraries were pooled, loaded onto an Ion 540 chip at 25 pM, and sequenced on an Ion GeneStudio S5 Plus Series (Thermo Fisher Scientific).

### TCR sequencing data analysis

Raw sequencing data were analyzed using Ion Reporter v5.20.2.0 (Thermo Fisher Scientific). For quality control, off-target and unproductive reads were excluded from downstream analyses. Read classification is shown in *Supp. File S1*, *Table 1*. The software identified V(D)J rearrangements and calculated the main TCR repertoire metrics, including richness, convergence, diversity, and evenness. TCR richness was defined as the total number of unique TCRβ/γ nucleotide sequences (clones). TCR convergence was defined as the cumulative frequency of clones sharing the same variable gene and CDR3 amino acid sequence. Diversity (Shannon diversity index) and evenness (normalized Shannon diversity) were calculated according to Equations (1-2), where *p_i_* represents the frequency of clone *i* in a sample containing *n* unique clones. Evenness describes how uniformly clones are distributed within the TCR repertoire, ranging from 0 to 1. Lower values indicate a repertoire dominated by a small number of predominant clones, whereas values approaching 1 indicate a more uniform distribution of clone frequencies.

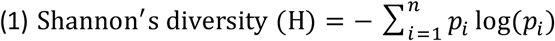

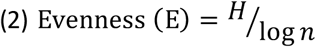

In addition, clones were classified according to their relative abundance within each repertoire using clonal-space metrics. Clonal space was quantified as the cumulative frequency of the most abundant clones and was evaluated for the top 1%, top 3%, and top 5% of the repertoire. For example, the top 1% clonal space was defined as the cumulative frequency of the top 1% most abundant clones. For survival analyses, all variables were dichotomized into “high” and “low” groups using the median value as the cutoff.

### Statistical analysis

Statistical analyses were performed using R environment (version 4.4.1). A p value < 0.05 was considered statistically significant. TCR repertoire variables were correlated with clinicopathological data (age, sex and histology), clinical response and survival. Overall survival (OS) was defined as the time from treatment initiation to death or last follow-up, and progression-free survival (PFS) was defined as time from treatment initiation to progression or death (whichever is earlier), or last follow-up. Mann-Whitney U test and Student’s t-test were used to study the relationship between TCR repertoire variables and clinical-pathological data and clinical response. The choice of appropriate test (Mann-Whitney or Student’s t) was based on normality (Shapiro-Wilk test) and variance homogeneity (Levene test). Spearman rank-correlation analysis was used for variable correlations. Contingency table analyses when comparing clinicopathological data with clinical response or clinicopathological data with TCR repertoire variables were performed using Fisher’s exact test. Kaplan-Meier survival analyses and log-rank test were used for time-dependent variables such as OS and PFS. For individual variables, receiver operating characteristic (ROC) curve analyses were performed to evaluate response prediction, and areas under the curve (AUC), sensitivity and specificity values were obtained. The Youden Index method was used to select the best cut-off values for all classification analyses^51^. A random forest classifier (1000 trees) was used to construct the combined predictive model. To enhance model diversity and reduce overfitting, the number of features sampled for splitting at each node was set to the square root of the total number of predictors. Model performance was evaluated by ROC curves derived using leave-one-out cross-validation (LOOCV) by iteratively training the model on all samples except one and testing on the excluded sample, in line with standard practice for a robust model estimation in small datasets^52^.

## Results

### Tumor-infiltrating TCRβ evenness predicts immunotherapy response

This retrospective observational study included FFPE samples from a total of 20 patients diagnosed with RCC, most with advance-stage disease (III-IV), treated with ICI-based regimens, including combination with targeted therapy (*Table 1*) mostly in the first-line setting. FFPE samples were collected before treatment (baseline) and were subjected to TCRβ/γ sequencing.

First, we evaluated the effect of immunotherapy response on patient prognosis. As expected, responders (R) had significantly longer PFS and OS than non-responders (NR) (*Fig. 1A-B*; *p* = 0.0025 and *p* = 0.0290, respectively). There was no association between any clinicopathological characteristic (age, sex or histology) and immunotherapy response. Second, we sought possible associations between the clinicopathological characteristics and TCR repertoire variables. We detected an association between histology and tumor-infiltrating TCRβ evenness (*Supp. File S1, Figure 1*; *p* = 0.0290). Sex was also associated with tumor-infiltrating TCRβ variables such as top 1% and top 3% clonal spaces (*Supp. File S1, Figure 2-3*; *p* = 0.0240 and *p* = 0.0460, respectively), and TCRγ variables such as diversity, top 1%, top 3% and top 5% clonal spaces (*Supp. File S1, Figure 4-7*; *p* = 0.0190, *p* = 0.0370, *p* = 0.0130 and *p* = 0.0140 respectively).

**Figure 1:**
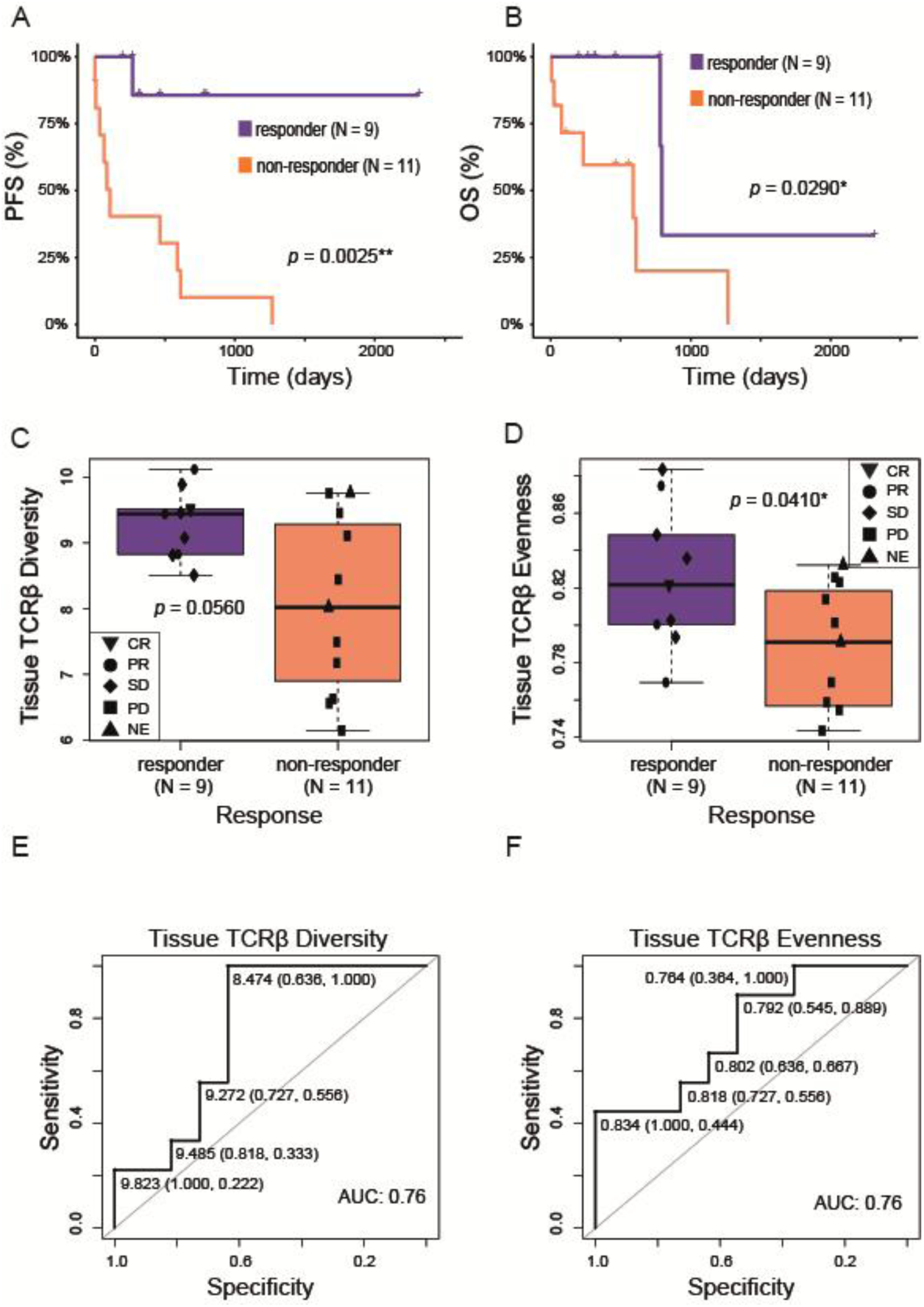
A) Responders (R) had significantly longer PFS than non-responders (NR) (p = 0.0025). P was obtained using log-rank test. B) R had significantly longer OS than NR (p = 0.0290). P was obtained using log-rank test. C) R showed a nonsignificant trend toward higher tumor-infiltrating TCRβ diversity than NR (p = 0.0560). P was obtained using Mann-Whitney test. D) R had significantly higher tumor-infiltrating TCRβ evenness than NR (p = 0.0410). P was obtained using Student’s t test. E) Tumor-infiltrating TCRβ diversity predicts response with an AUC of 0.76 (95% CI: 0.541-0.974). At a cutoff > 8.474, tumor-infiltrating TCRβ diversity yielded 100% sensitivity (95% CI: 66-100%) and 63.6% specificity (95% CI: 31-89%). F) Tumor-infiltrating TCRβ evenness predicts response with an AUC of 0.76 (95% CI: 0.550-0.964). At a cutoff > 0.834, tumor-infiltrating TCRβ evenness yielded 44.4% sensitivity (95% CI: 14-79%) and 100% specificity (95% CI: 72-100%). AUC: Area Under the Curve, CR: complete response, N: number of patients, NE: non-evaluable, OS: overall survival, PD: progressive disease, PFS: progression-free survival, PR: partial response, SD: stable disease, *: statistical significance, **: strong statistical significance (p < 0.01).

**Figure 2:**
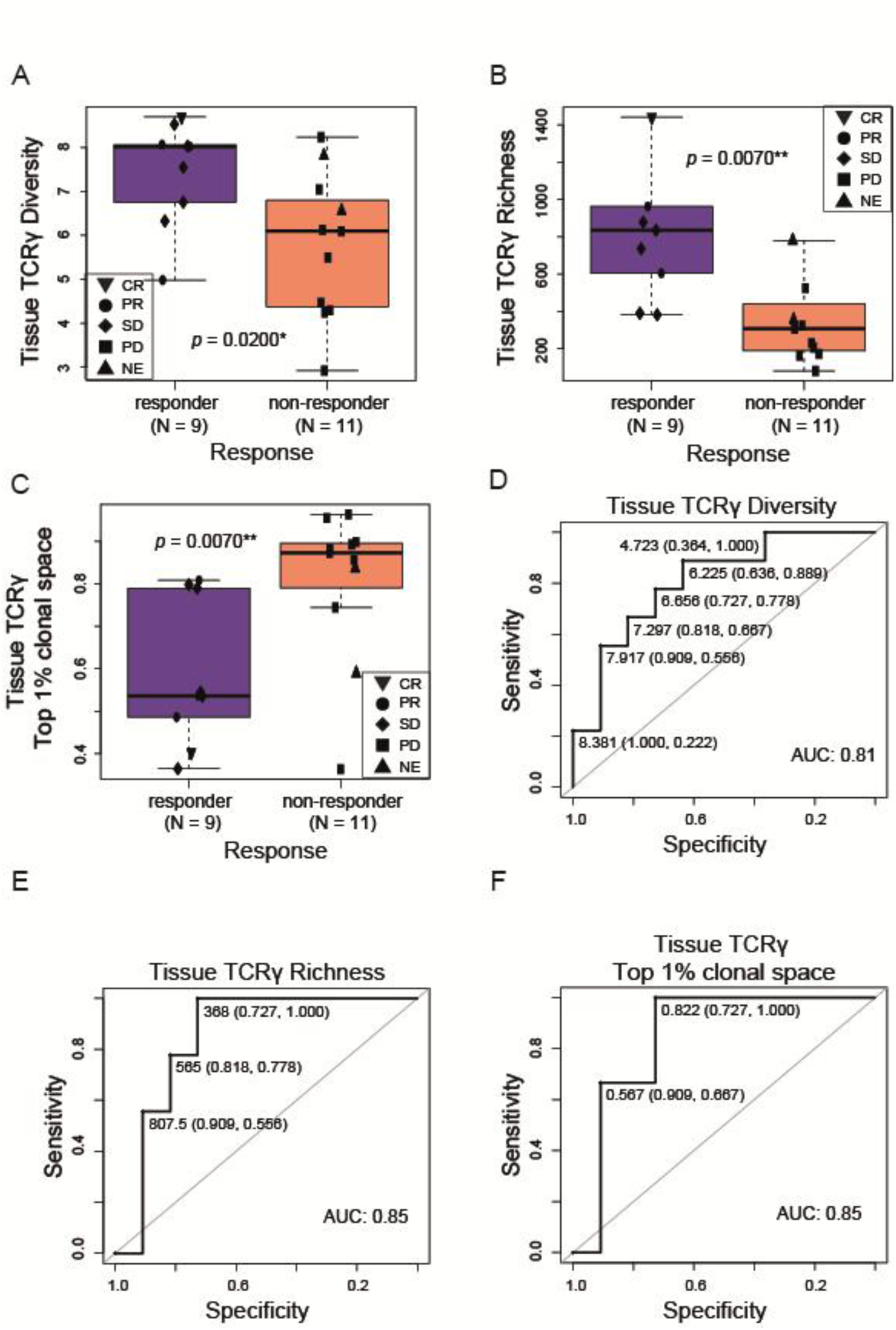
A) Responders (R) had significantly higher tumor-infiltrating TCRγ diversity than non-responders (NR) (p = 0.0200). P was obtained using Student’s t test. B) R had significantly higher tumor-infiltrating TCRγ clones than NR (p = 0.0070). P was obtained using Mann-Whitney test. C) R had significantly lower tumor-infiltrating TCRγ top 1% clonal space than NR (p = 0.0070). P was obtained using Mann-Whitney test. D) Tumor-infiltrating TCRγ diversity predicts response with an AUC of 0.81 (95% CI: 0.621-0.995). At a cutoff > 6.225, tumor-infiltrating TCRγ diversity yielded 88.8% sensitivity (95% CI: 52-100%) and 63.6% specificity (95% CI: 31-89%). E) Tumor-infiltrating TCRγ richness discriminates response with an AUC of 0.85 (95% CI: 0.664-1.000). At a cutoff > 368, tumor-infiltrating TCRγ richness yielded 100% sensitivity (95% CI: 66-100%) and 72.7% specificity (95% CI: 39-94%). F) Tumor-infiltrating TCRγ top 1% clonal space predicts response with an AUC of 0.85 (95% CI: 0.655-1.000). At a cutoff < 0.822, tumor-infiltrating TCRγ top 1% clonal space yielded 100% sensitivity (95% CI: 66-100%) and 72.7% specificity (95% CI: 39-94%). AUC: Area Under the Curve, CR: complete response, N: number of patients, NE: non-evaluable, PD: progressive disease, PR: partial response, SD: stable disease, *: statistical significance, **: strong statistical significance (p < 0.01).

**Figure 3:**
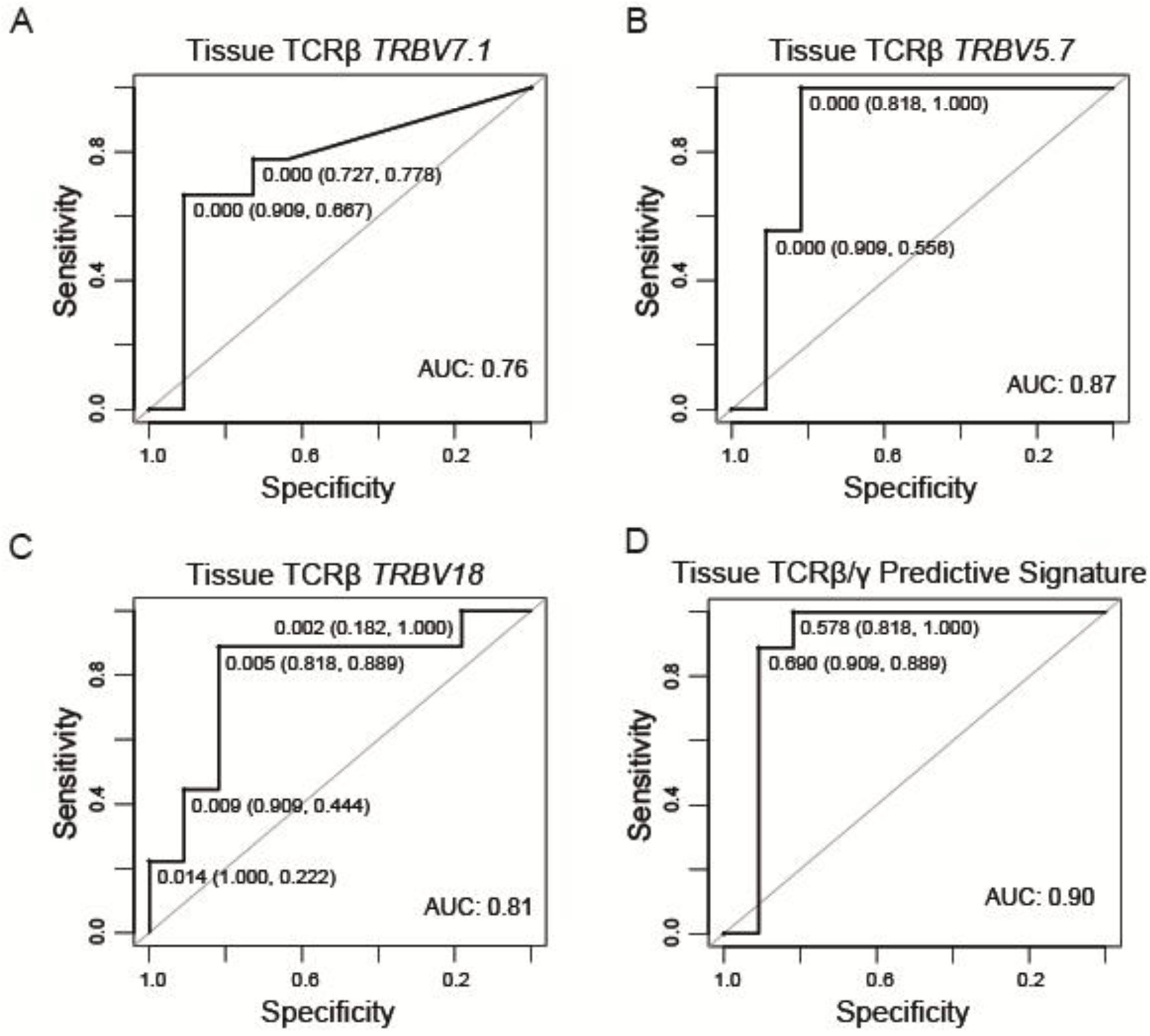
A) Tumor-infiltrating TCRβ TRBV7.1 frequency predicts response with an AUC of 0.76. (95% CI: 0.475-1.000). At a cutoff > 3.69E-6, tumor-infiltrating TCRβ TRBV7.1 frequency yielded 66.7% sensitivity (95% CI: 30-93%) and 90.9% specificity (95% CI: 59-100%). B) Tumor-infiltrating TCRβ TRBV5.7 frequency predicts response with an AUC of 0.87 (95% CI: 0.687-1.000). At a cutoff > 3.55E-6, tumor-infiltrating TCRβ TRBV5.7 frequency yielded 100% sensitivity (95% CI: 66-100%) and 81.8% specificity (95% CI: 48-98%). C) Tumor-infiltrating TCRβ TRBV18 frequency predicts response with an AUC of 0.81 (95% CI: 0.603-1.000). At a cutoff > 0.005, tumor-infiltrating TCRβ TRBV18 frequency yielded 88.9% sensitivity (95% CI: 52-100%) and 81.8% specificity (95% CI: 48-98%). D) A tumor-infiltrating predictive signature including TCRβ TRBV5.7 frequency, TCRγ richness and TCRγ top 1% clonal space predicts response with an AUC of 0.90 (95% CI: 0.721-1.000). At the selected threshold, the signature yielded 100% sensitivity (95% CI: 66-100%) and 81.8% specificity (95% CI: 48-98%). AUC: Area Under the Curve.

With respect to TCRβ repertoire, we found that R showed a non-significant trend toward higher tumor-infiltrating TCRβ diversity than NR (*Fig. 1C*; *p* = 0.0560). Indeed, following the same trend, R had significantly higher tumor-infiltrating TCRβ evenness than NR (*Fig. 1D*; *p* = 0.0410). We next evaluated the ability of these variables to discriminate responders from non-responders. Tumor-infiltrating TCRβ diversity predicted response with an AUC of 0.76 (95% CI: 0.541-0.974), where a cutoff of tumor-infiltrating TCRβ diversity > 8.474 yielded 100% sensitivity (95% CI: 66-100%) and 63.6% specificity (95% CI: 31-89%) for discriminating responders from non-responders (*Fig. 1E*). Similarly, tumor-infiltrating TCRβ evenness predicted response with an AUC of 0.76 (95% CI: 0.550-0.964), where a cutoff of tumor-infiltrating TCRβ evenness > 0.834 yielded 44.4% sensitivity (95% CI: 14-79%) and 100% specificity (95% CI: 72-100%) for discriminating responders from non-responders (*Fig. 1F*). The similarity between these two variables was expected, as they were highly positively correlated (*Supp. File S2)*.

### Tumor-infiltrating TCRγ diversity, clonal richness and top 1% clonal space are associated with immunotherapy response

With respect to TCRγ repertoire, we found that R had significantly higher tumor-infiltrating TCRγ diversity and richness than NR (*Fig. 2A-B*; *p* = 0.0200 and *p* = 0.0070, respectively). Furthermore, we also detected significantly lower tumor-infiltrating TCRγ top 1% clonal space in R than in NR (*Fig. 2C*; *p* = 0.0070). Similarly, tumor-infiltrating TCRγ top 3% and top 5% clonal spaces were significantly lower in R (*Supp. File S3, Figure 1-2*; *p* = 0.0090 and *p* = 0.0100, respectively). These results are again consistent with the correlations observed among the variables (*Supp. File S4*).

We then evaluated their ability to discriminate responders from non-responders. Tumor-infiltrating TCRγ diversity predicted response with an AUC of 0.81 (95% CI: 0.621-0.995), where a cutoff of tumor-infiltrating TCRγ diversity > 6.225 yielded 88.8% sensitivity (95% CI: 52-100%) and 63.6% specificity (95% CI: 31-89%) (*Fig. 2D*). Tumor-infiltrating TCRγ richness discriminated response with an AUC of 0.85 (95% CI: 0.664-1.000), where a cutoff of tumor-infiltrating TCRγ richness > 368 yielded 100% sensitivity (95% CI: 66-100%) and 72.7% specificity (95% CI: 39-94%) (*Fig. 2E*). Tumor-infiltrating TCRγ top 1% clonal space predicted response with an AUC of 0.85 (95% CI: 0.655-1.000), where a cutoff of tumor-infiltrating TCRγ top 1% clonal space < 0.822 yielded 100% sensitivity (95% CI: 66-100%) and 72.7% specificity (95% CI: 39-94%).

Moreover, patients with high tumor-infiltrating TCRγ richness (> 457.5) had significantly longer PFS than patients with low (≤ 457.5) tumor-infiltrating TCRγ clones (*Supp. File S3, Figure 3*; *p* = 0.0110). Conversely, patients with low tumor-infiltrating TCRγ top 1% clonal space (< 0.7934) had significantly longer PFS than patients with high (≥ 0.7934) tumor-infiltrating TCRγ top 1% clonal space (*Supp. File S3, Figure 4*; *p* = 0.0190).

### Tumor-infiltrating TCRβ TRBV and TRBJ gene usage is associated with immunotherapy response

Next, we examined associations between TRBV/TRGV and TRBJ/TRGJ gene frequencies and the immunotherapy response (*Supp. File S3, Figure 5-6*). Regarding TCRγ, there was no association between TRGV and TRGJ gene frequencies and clinical response. Interestingly, we found several tumor-infiltrating TCRβ gene frequencies that were significantly higher in R than in NR (*Supp. File S3, Table 1*). Among these genes, only *TRBV7.1*, *TRBV5.7* and *TRBV18* were associated with PFS, with higher frequencies of each associated with longer PFS (*Supp. File S3, Figure 7-9*; *p* = 0.0270, *p* = 0.0022 and *p* = 0.0340, respectively). We then evaluated their ability to discriminate responders from non-responders: (1) TCRβ *TRBV7.1* frequency predicted response with an AUC of 0.76 (95% CI: 0.475-1.000), where a cutoff > 3.69E-6 yielded 66.7% sensitivity (95% CI: 30-93%) and 90.9% specificity (95% CI: 59-100%) (*Fig. 3A*). (2) TCRβ *TRBV5.7* frequency predicted response with an AUC of 0.87 (95% CI: 0.687-1.000), where a cutoff > 3.55E-6 yielded 100% sensitivity (95% CI: 66-100%) and 81.8% specificity (95% CI: 48-98%) (*Fig. 3B*). (3) TCRβ *TRBV18* frequency predicted response with an AUC of 0.81 (95% CI: 0.603-1.000), where a cutoff > 0.005 yielded 88.9% sensitivity (95% CI: 52-100%) and 81.8% specificity (95% CI: 48-98%) (*Fig. 3C*).

### Combined TCRβ/γ signature to predict immunotherapy response

Finally, we developed a Random Forest predictive signature for immunotherapy response. From TCRβ variables we included *TRBV5.7* frequency because it was associated with PFS and had the highest ROC AUC. For the same reason, among TCRγ variables, we selected richness and top 1% clonal space. The combined TCRβ/γ signature achieved a cross-validated AUC of 0.90 (95% CI: 0.721-1.000) for discriminating clinical response. A positive result from this signature (probability of being classified as responder ≥ 0.578) yielded 100% sensitivity (95% CI: 66-100%) and 81.8% (95% CI: 48-98%) specificity (*Fig. 3D*).

## Discussion

Since its establishment as the standard of care for advanced RCC, ICIs have drastically improved clinical outcomes. However, no validated biomarker is currently available to guide treatment selection. Although findings have been inconsistent, the TCR repertoire has emerged as a potential predictive biomarker of response to ICIs across several solid tumors^8,9,13,29–42^.

To our knowledge, this represents the first characterization of the baseline tumor-infiltrating TCRβ/γ repertoire in patients predominantly receiving first-line ICI-based therapy. In fact, most TCR repertoire studies in solid tumors have focused on TCRαβ cells because of the broader understanding of their molecular regulation and function ^8,9,13,29–32,34,37–42^. However, increasing evidence over the last decade has highlighted the important role of TCRγδ cells in antitumor immunity, contributing to both systemic and local immunosurveillance across many solid cancers ^35,36,43–45^, including RCC ^43,46–49^. Indeed, the baseline circulating abundance of γδ T cells has been associated with improved survival in both localized ^53^ and advanced RCC ^54^, and a γδ T cell-associated prognostic signature has been shown to predict survival in patients with ccRCC^55^. Nevertheless, studies characterizing the TCRγ repertoire, particularly those investigating its relationship with clinical outcomes, remain scarce.

We confirmed that responder patients (R) achieved improved PFS and OS, while ICIs response showed no significant association with age, sex, or histological subtype, highlighting the need for reliable predictive biomarkers to guide treatment selection at baseline. Interestingly, some TCRβ/γ repertoire features appeared to be influenced by histology or sex; however, none of these clinicopathological variables were predictive of clinical response. Previous findings have been inconsistent: while Xu *et al.* also found lower tumor-infiltrating TCRβ evenness in ccRCC compared to nccRCC tumors ^39^, Gadot *et al.* did not find such an association ^37^. Although sex-related differences in the immune microenvironment of RCC have been reported ^56^, no studies have specifically examined associations between sex and TCR repertoire metrics in RCC patients.

Our results showed that R exhibited higher tumor-infiltrating TCRβ evenness at baseline, with a moderate capacity to predict clinical response. The question of whether tumor-specific T cells activated by ICIs pre-exist in the tumor ^13,57,58^ or are replaced by new T cell clones recruited to the tumor microenvironment ^59,60^ also remains under debate. Au *et al.* observed both an expansion of novel CD8+ T cell clones and maintenance of previously expanded CD8+ T cell clones after anti-PD-1 treatment in advanced ccRCC, with only the latter appearing to be directly associated with clinical outcomes ^13^. Moreover, responders harboured significantly higher tumor-infiltrating T cells both at baseline and post-treatment, with enrichment of ‘‘Immune-activation’’ and ‘‘TCR signaling’’ pathways ^13^. One possible explanation is that a more even baseline TCR repertoire provides a broader repertoire of tumor-reactive T-cell clones that can be reinvigorated by immune checkpoint blockade, thereby enhancing tumor recognition and antitumor immunity. Ross-Macdonald *et al.* also observed a trend of higher baseline tumor-infiltrating TCRβ evenness in responders with advanced ccRCC treated with anti-PD-1 ^40^. Conversely, Au *et al.* and Casarrubios *et al.* reported lower baseline tumor-infiltrating TCRβ evenness in responders with advanced ccRCC treated with anti-PD-1 ^13^ and NSCLC treated with neoadjuvant chemoimmunotherapy ^32^, suggesting a pre-existing intratumoral clonal expansion that is maintained after therapy. Similar results were reported in a small cohort by Gadot *et al.*, with non-recurring advanced ccRCC patients having lower baseline tumor-infiltrating TCRβ evenness. Interestingly, they also showed a tendency towards a higher richness in non-recurring patients ^37^.

Studies in lung ^29,30^, cervical ^8^, pancreatic ^9^ and renal ^41^ cancer have shown that higher baseline circulating TCRβ diversity predicts better prognosis and many others have also studied how it changes during treatment, with inconsistent results. Guo *et al.* found that circulating TCRβ diversity was associated with high numbers of CD4+ and CD8+ T cells, whereas lower diversity was associated with more neutrophils or protumour regulatory T cells, in advanced RCC ^41^. Conversely, Han *et al.* ^30^, Hopkins *et al.* ^9^ and Kato *et al.* ^38^ reported that decreases in circulating TCRβ diversity or evenness were associated with improved survival in lung, pancreatic and renal cancer, respectively. Yet, Carlisle *et al.* reported no changes in circulating TCRβ diversity during treatment in advanced ccRCC treated with ICIs ^42^. Nevertheless, we believe the study of the TCR repertoire as a biomarker for ICIs response should be evaluated prior first-line treatment to have a real impact on daily clinical practice. It is important to note that the prognostic value of the TCR repertoire may also depend on the type of immune checkpoint inhibitor and combination administered, as reported in lung ^31^ and melanoma ^34^.

We also found that specific TCRβ genes were associated with response to immunotherapy. Interestingly, all of them showed higher frequencies in R. Furthermore, *TRBV7.1*, *TRBV5.7*, and *TRBV18* were also associated with survival benefit, with higher frequencies of these genes corresponding to longer PFS. The association between treatment response and increased frequencies of specific tumor-infiltrating TRBV genes may reflect preferential expansion of clonotypes using these V genes in response to tumor-associated antigens. However, antigen specificity cannot be inferred from bulk TCR sequencing alone.

With respect to TCRγ, R showed greater tumor-infiltrating TCRγ diversity and clonal richness, together with a reduced top 1% clonal space at baseline, all showing good discrimination for clinical response in this cohort. The latter two parameters were also associated with longer PFS. As observed for TCRβ, these findings may reflect a more balanced TCR repertoire and a broader pool of potentially tumor-reactive T-cell clones, although antigen specificity cannot be determined from these data. To the best of our knowledge, this is the first report linking intratumoral TCRγ repertoire metrics with immunotherapy response in advanced RCC. As reported in our previous study in NSCLC ^33^, TCRγ gene usage was not associated with treatment response, possibly reflecting the more constrained TCRγ repertoire ^27^.

Accordingly, we integrated the top predictive variables from each repertoire to construct a combined TCRβ/γ signature using a Random Forest classifier. This model showed high apparent discrimination, with 100% sensitivity, 81.8% specificity, and a cross-validated AUC of 0.90, with all responders correctly classified at the selected threshold in this cohort. Nonetheless, approximately 18.2% of non-responders would be misclassified, potentially resulting in overtreatment. From a translational standpoint, this tissue-based approach warrants direct comparison with established biomarkers in independent cohorts. However, due to the unavailability of PD-1/PD-L1 expression data in our cohort, a direct comparison could not be performed. In addition, finding minimally invasive circulating biomarkers that are easily measurable throughout therapy would facilitate their implementation in daily clinical practice ^38,41^.

In conclusion, this study provides the first comprehensive characterization of the baseline intratumoral TCRβ and TCRγ repertoires in patients with advanced RCC predominantly receiving first-line immune checkpoint inhibitor-based therapy. Despite RCC being one of the most immunogenic tumor types and among the earliest malignancies to benefit from immune checkpoint blockade, the predictive value of the intratumoral TCR repertoire (particularly the TCRγ repertoire) has remained largely unexplored. We identified multiple baseline TCRβ and TCRγ features associated with both clinical response and progression-free survival and developed a combined predictive signature integrating the most informative variables from both repertoires. Given the exploratory nature of the study, the small cohort size, and the lack of an independent validation cohort, prospective validation in larger patient series is required. Our findings were generated using routinely available pretreatment FFPE tumor samples collected across three referral hospitals in a real-world clinical setting, highlighting the feasibility and translational potential of TCR repertoire profiling as candidate biomarker for pretreatment patient stratification in advanced RCC.

## Acknowledgements

The authors thank all enrolled patients and their families. Samples were collected and stored by the Galicia Sur Health Research Institute (IIS Galicia Sur) Biobank (registry B.0000802).

## Author contributions

MPG contributed to conceptualization, performed formal analysis, curated the data, prepared the figures, and wrote the original draft of the manuscript. MLQ contributed to conceptualization, enrolled patients, curated the clinical data, and critically revised the manuscript. GdV contributed patient samples and critically revised the manuscript. MD and AFF contributed patient samples and clinical data curation and critically revised the manuscript. IAA performed and optimized DNA extraction and library preparation. PSF and JAO reviewed and selected the FFPE tumor samples. MGG and LCV contributed to data validation and interpretation. SG and EJL developed the sequencing methodology, performed TCR sequencing, and critically revised the manuscript. MMF conceived and designed the study, supervised the project, secured funding, administered the project, contributed to methodology, wrote and critically revised the manuscript. All authors reviewed, edited, and approved the final version of the manuscript.

## Data availability

The datasets supporting the findings of this study are available from the corresponding authors upon reasonable request.

