## Supplementary File S1 for "Baseline intratumoral TCRβ and TCRγ repertoires predict response to immune checkpoint inhibitors in advanced renal cell carcinoma"

*Table 1: Description of the type of reads based on their quality. INDEL, Insertion Deletion.*

| Thype of read | Description |
|---|---|
| **Off-target/low-quality** | Reads that are of low quality or represent the product of an off-target amplification. |
| **Unproductive** | Reads that have uncorrectable sequencing or PCR errors that lead the rearrangement to have out-of-frame variable and joining genes or a premature stop codon. |
| **Rescued productive** | Reads that have an in-frame variable and joining gene, and no stop codons after INDEL error correction. |
| **Productive** | Reads that have an in-frame variable and joining gene, and no stop codons. |

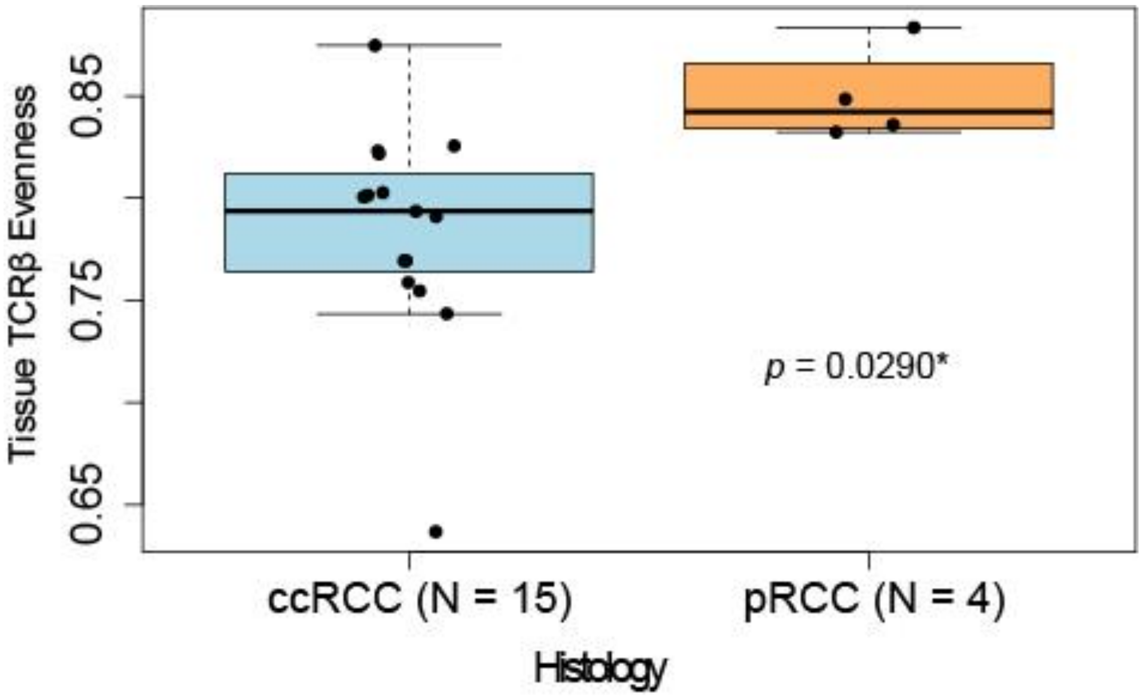


Figure 1: There is an association between histology and tumor-infiltrating TCRβ evenness,

where ccRCC have statistically significant lower tumor-infiltrating TCRβ evenness than pRCC (p = 0.0290). P was obtained using Student's t test.

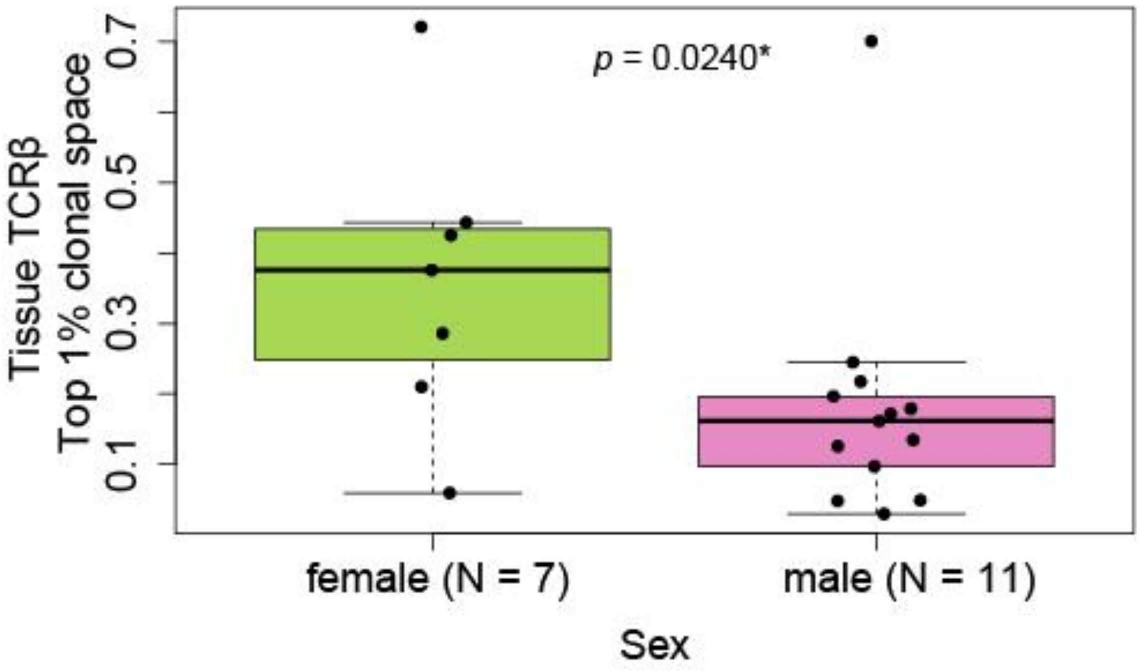


Figure 2: There is an association between sex and tumor-infiltrating TCRβ top 1% clonal space, where females have statistically significant higher tumor-infiltrating TCRβ top 1% clonal space than males (*p* = 0.0240). *P* was obtained using Mann-Whitney test.

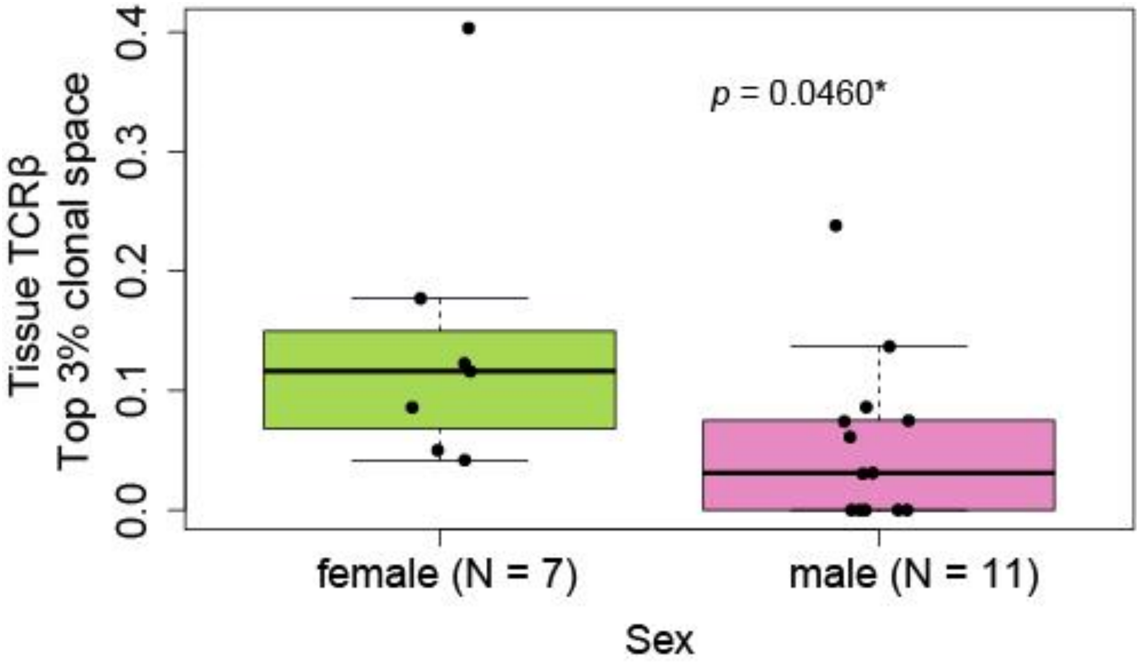


Figure 3: There is an association between sex and tumor-infiltrating TCRβ top 3% clonal space, where females have statistically significant higher tumor-infiltrating TCRβ top 3% clonal space than males (*p* = 0.0460). *P* was obtained using Mann-Whitney test.

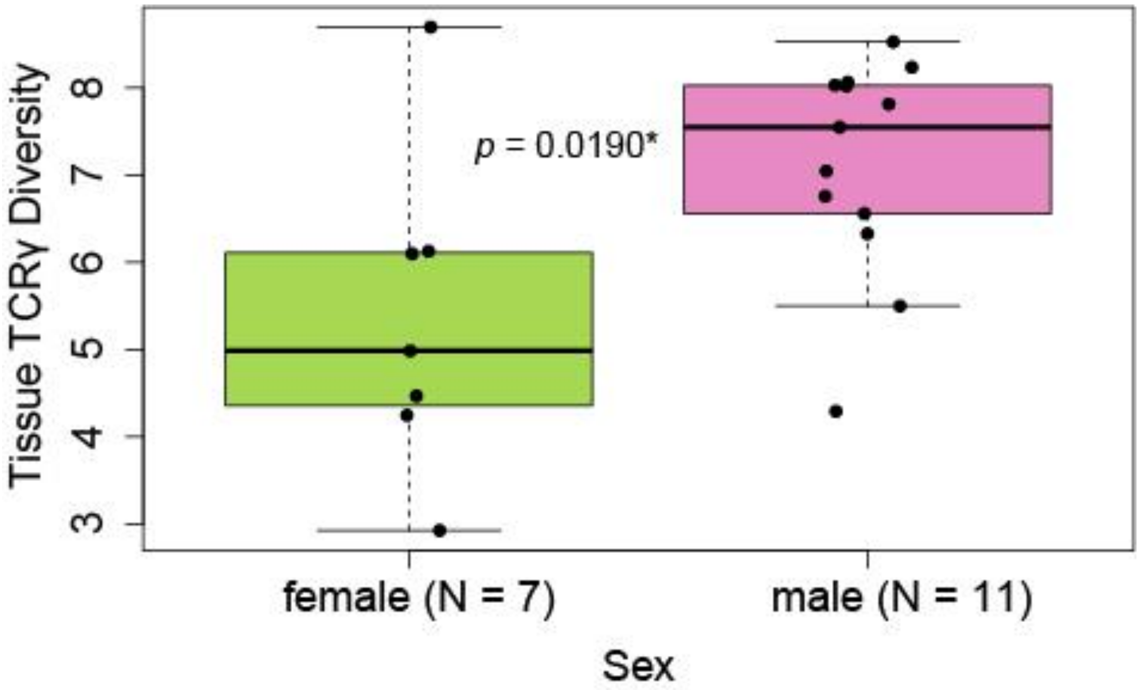


Figure 4: There is an association between sex and tumor-infiltrating TCRγ diversity, where females have statistically significant lower tumor-infiltrating TCRγ diversity than males (*p* = 0.0190). *P* was obtained using Student's t test.

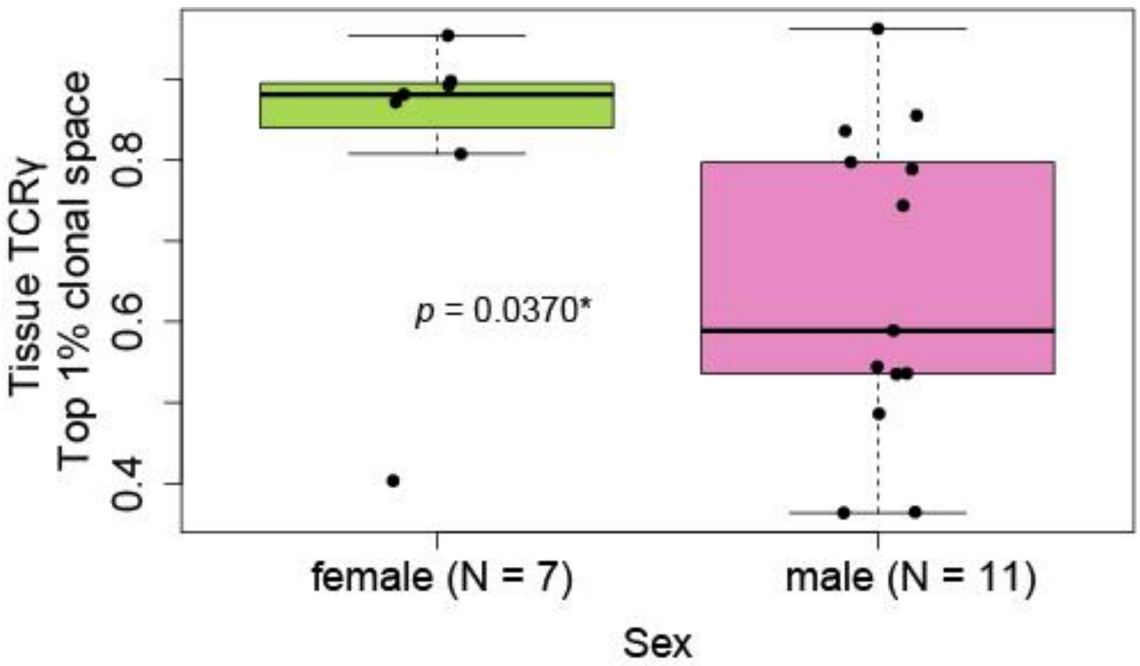


Figure 5: There is an association between sex and tumor-infiltrating TCRγ top 1% clonal space, where females have statistically significant lower tumor-infiltrating TCRγ top 1% clonal space than males (*p* = 0.0370). *P* was obtained using Mann-Whitney test.

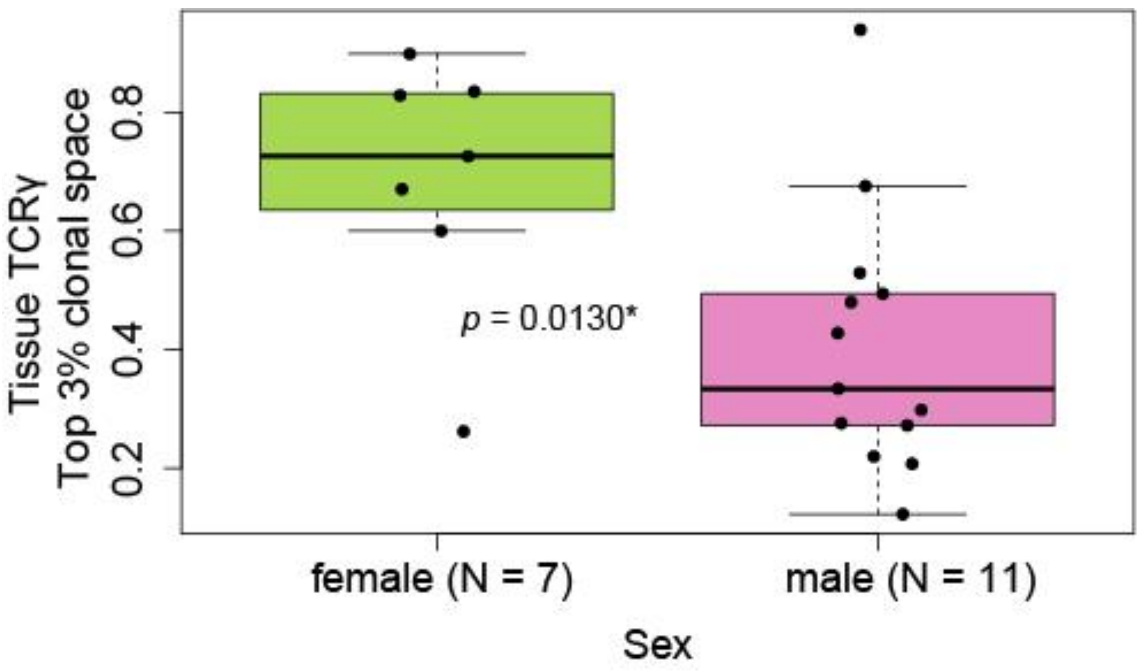


Figure 6: There is an association between sex and tumor-infiltrating TCRγ top 3% clonal space, where females have statistically significant lower tumor-infiltrating TCRγ top 3% clonal space than males (*p* = 0.0130). *P* was obtained using Student's t test.

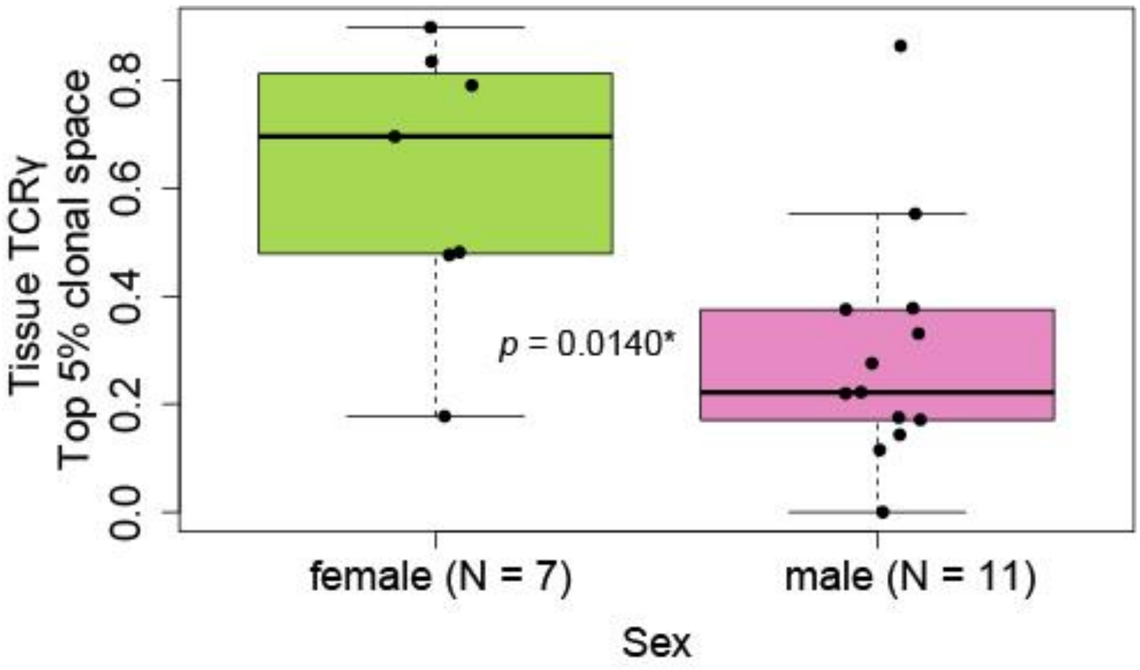


Figure 7: There is an association between sex and tumor-infiltrating TCRγ top 5% clonal space, where females have statistically significant lower tumor-infiltrating TCRγ top 5% clonal space than males (*p* = 0.0140). *P* was obtained using Mann-Whitney test.
