## Supplementary File S2 for "Baseline intratumoral TCRβ and TCRγ repertoires predict response to immune checkpoint inhibitors in advanced renal cell carcinoma"

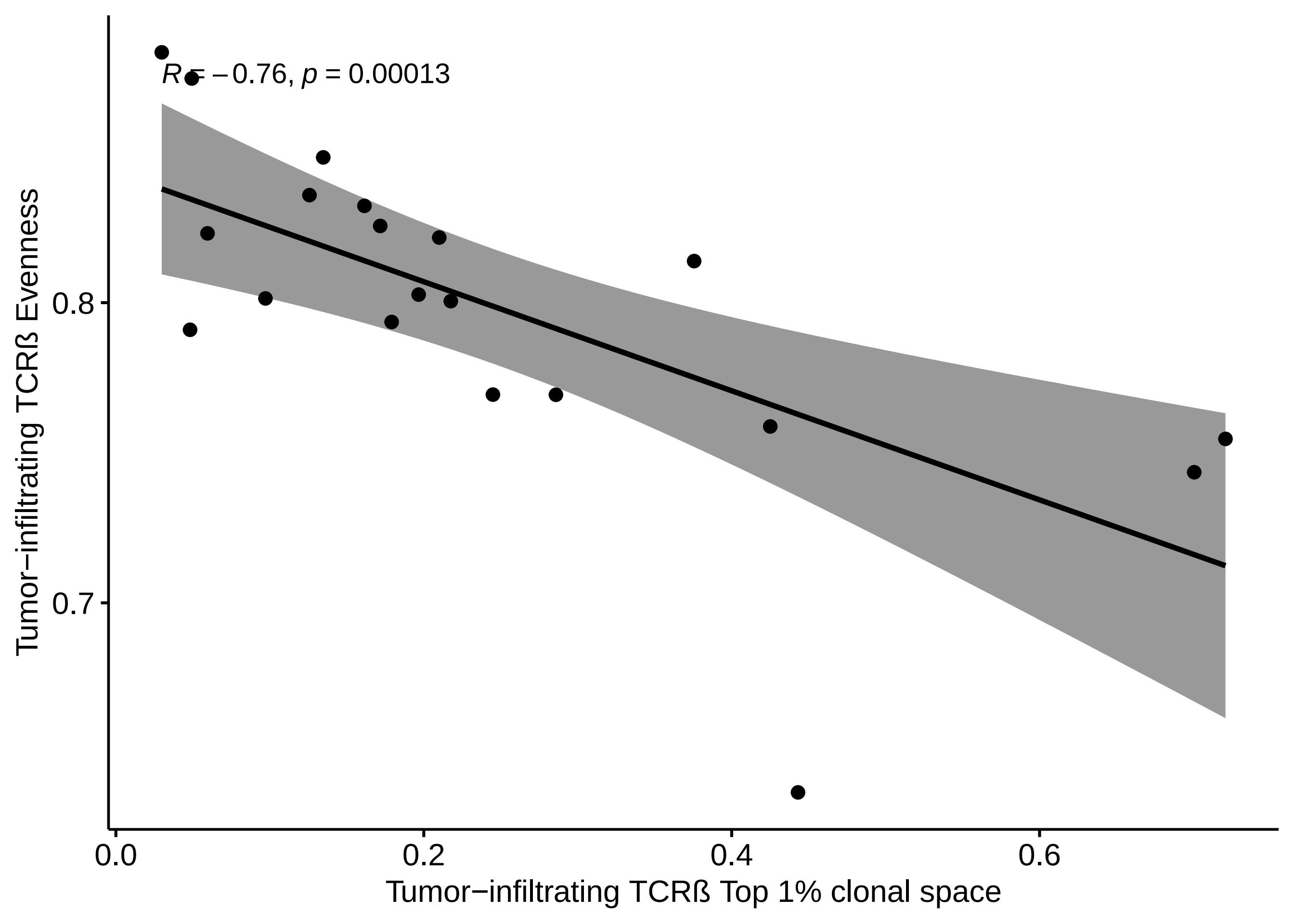

R = −0.76, p = 0.00013
Tumor−infiltrating TCRß Evenness
0.8
0.7
0.0
0.2
0.4
0.6
Tumor−infiltrating TCRß Top 1% clonal space

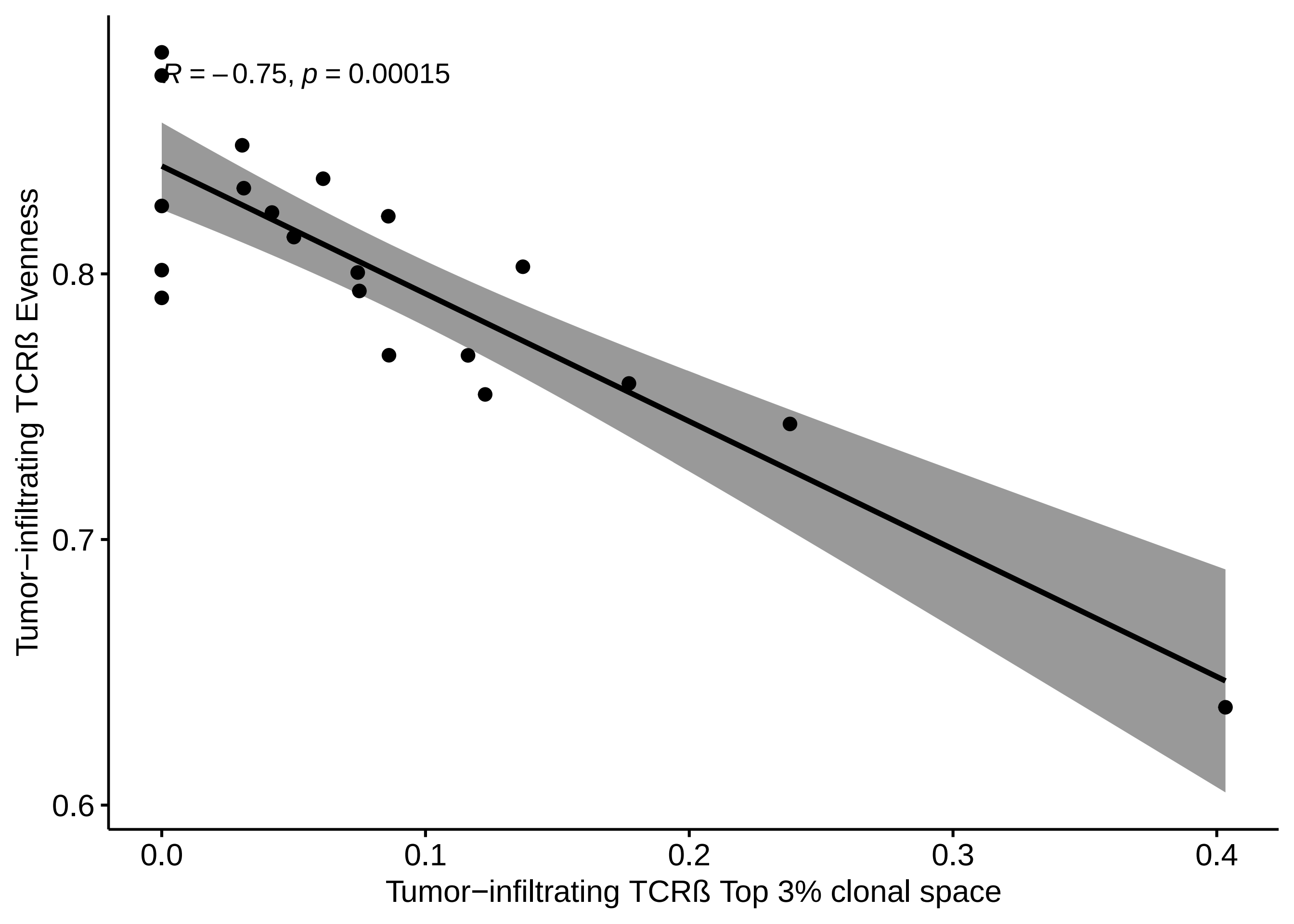

R = −0.75, p = 0.00015
0.6
0.7
0.8
0.0
0.1
0.2
0.3
0.4
Tumor−infiltrating TCRß Evenness
Tumor−infiltrating TCRß Top 3% clonal space

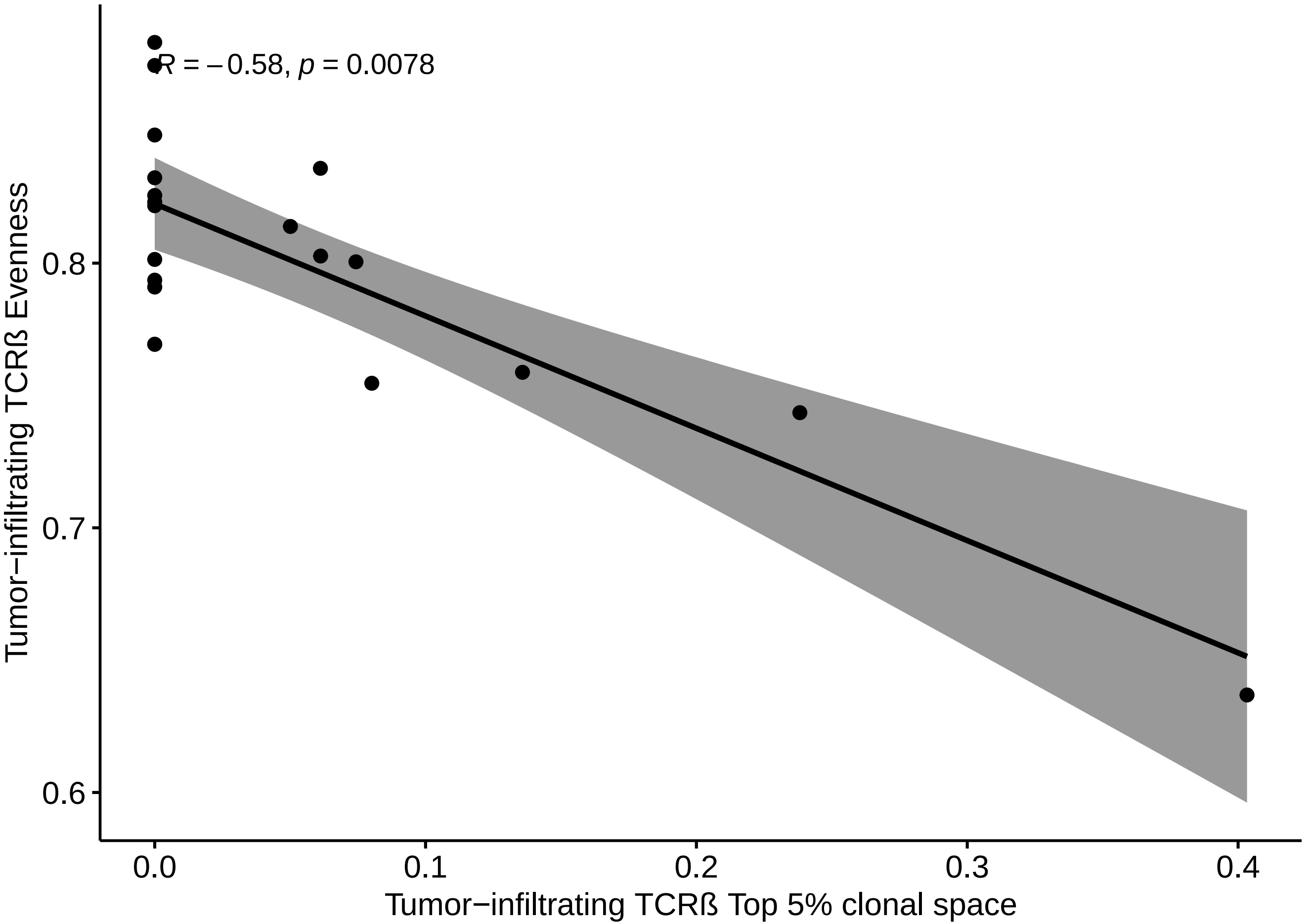

R = −0.58, p = 0.0078
Tumor−infiltrating TCRß Evenness
0.8
0.7
0.6
0.0
0.1
0.2
0.3
0.4
Tumor−infiltrating TCRß Top 5% clonal space

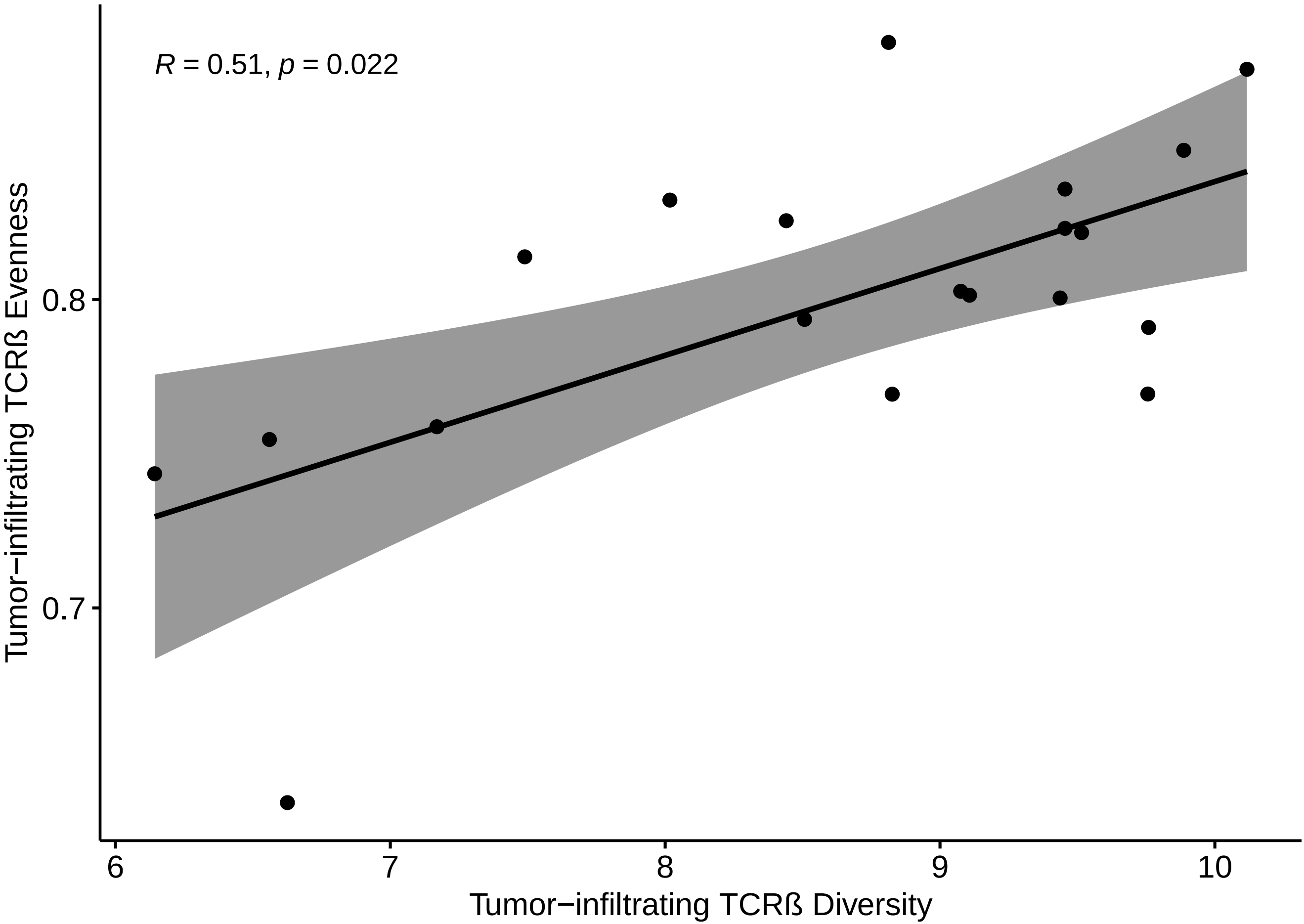

R = 0.51, p = 0.022
Tumor−infiltrating TCRß Evenness
0.8
0.7
6
7
8
9
10
Tumor−infiltrating TCRß Diversity

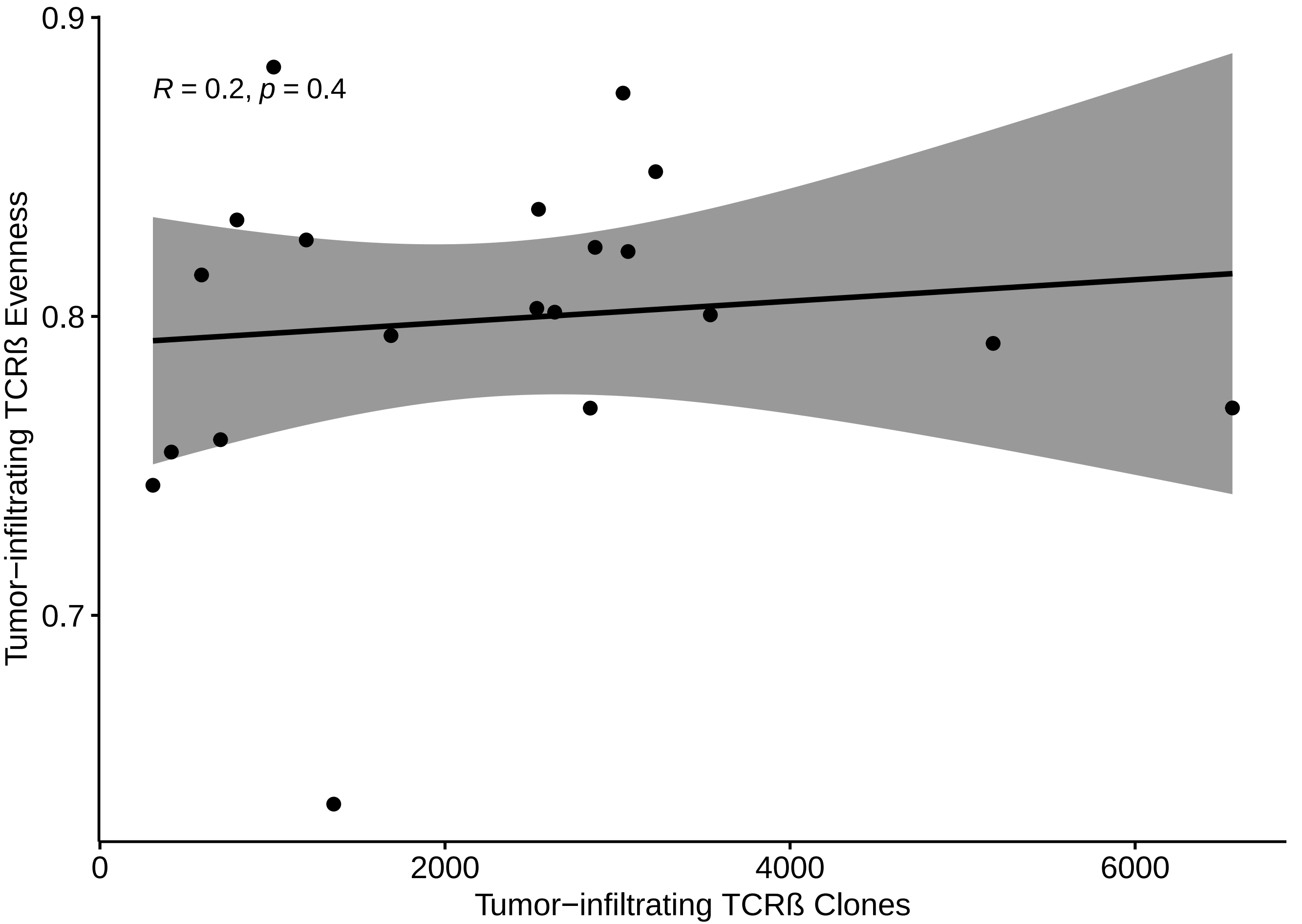

R = 0.2, p = 0.4
0.9
0.8
0.7
0
2000
4000
6000
Tumor−infiltrating TCRß Evenness
Tumor−infiltrating TCRß Clones

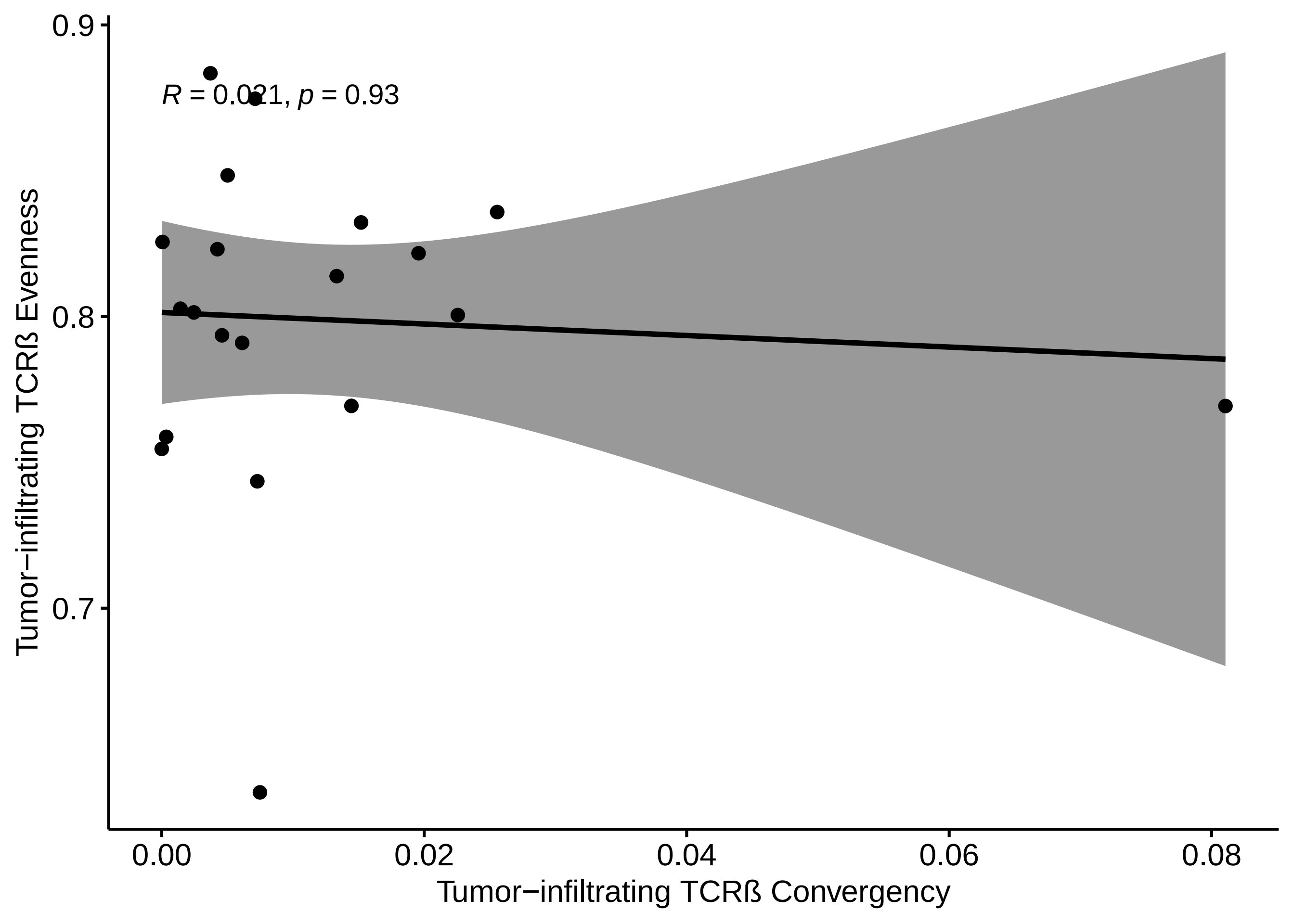
R = 0.021, p = 0.93
0.9
0.8
0.7
0.00
0.02
0.04
0.06
0.08
Tumor−infiltrating TCRß Evenness
Tumor−infiltrating TCRß Convergency

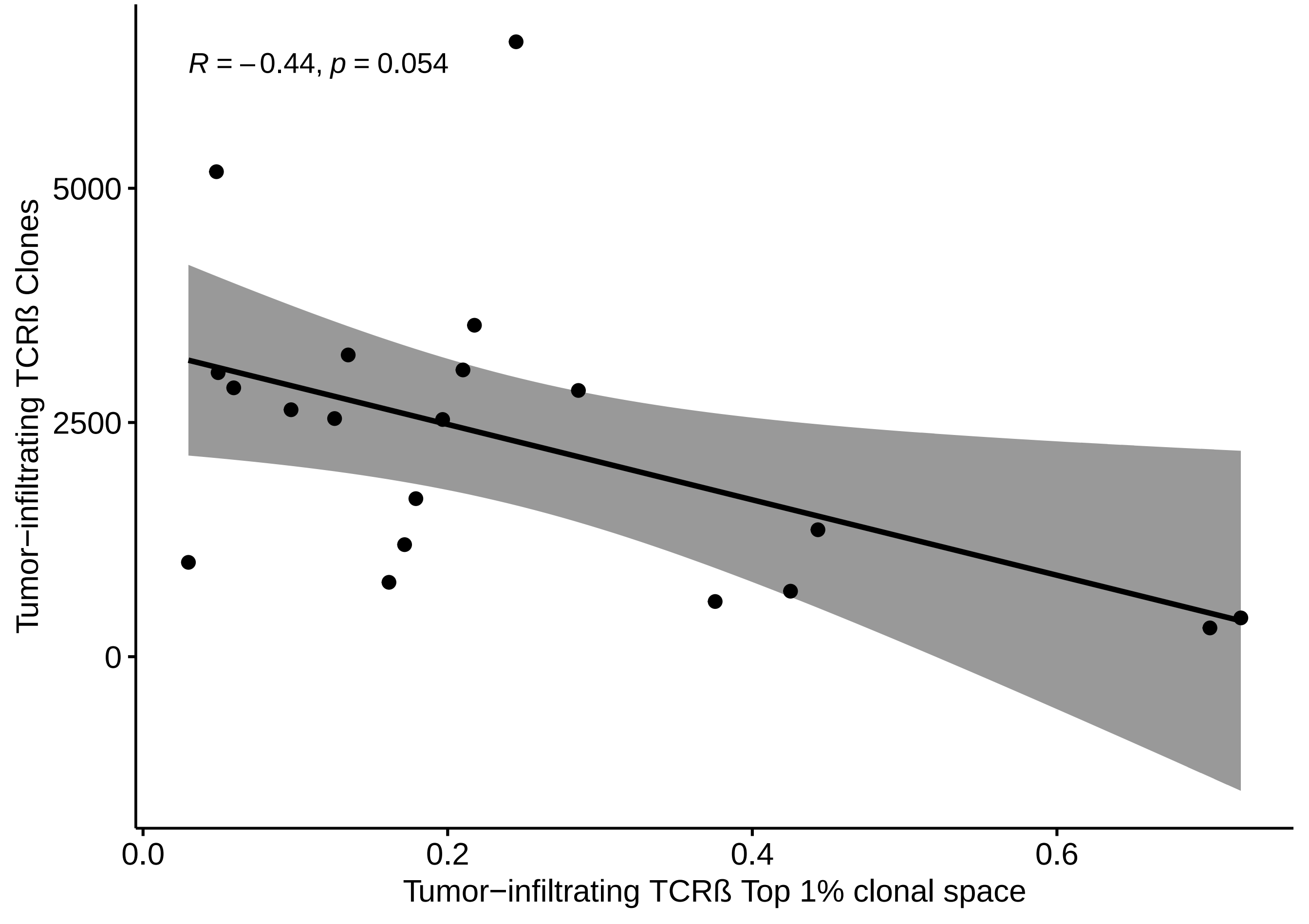

R = −0.44, p = 0.054
5000
2500
0
0.0
0.2
0.4
0.6
Tumor−infiltrating TCRß Clones
Tumor−infiltrating TCRß Top 1% clonal space

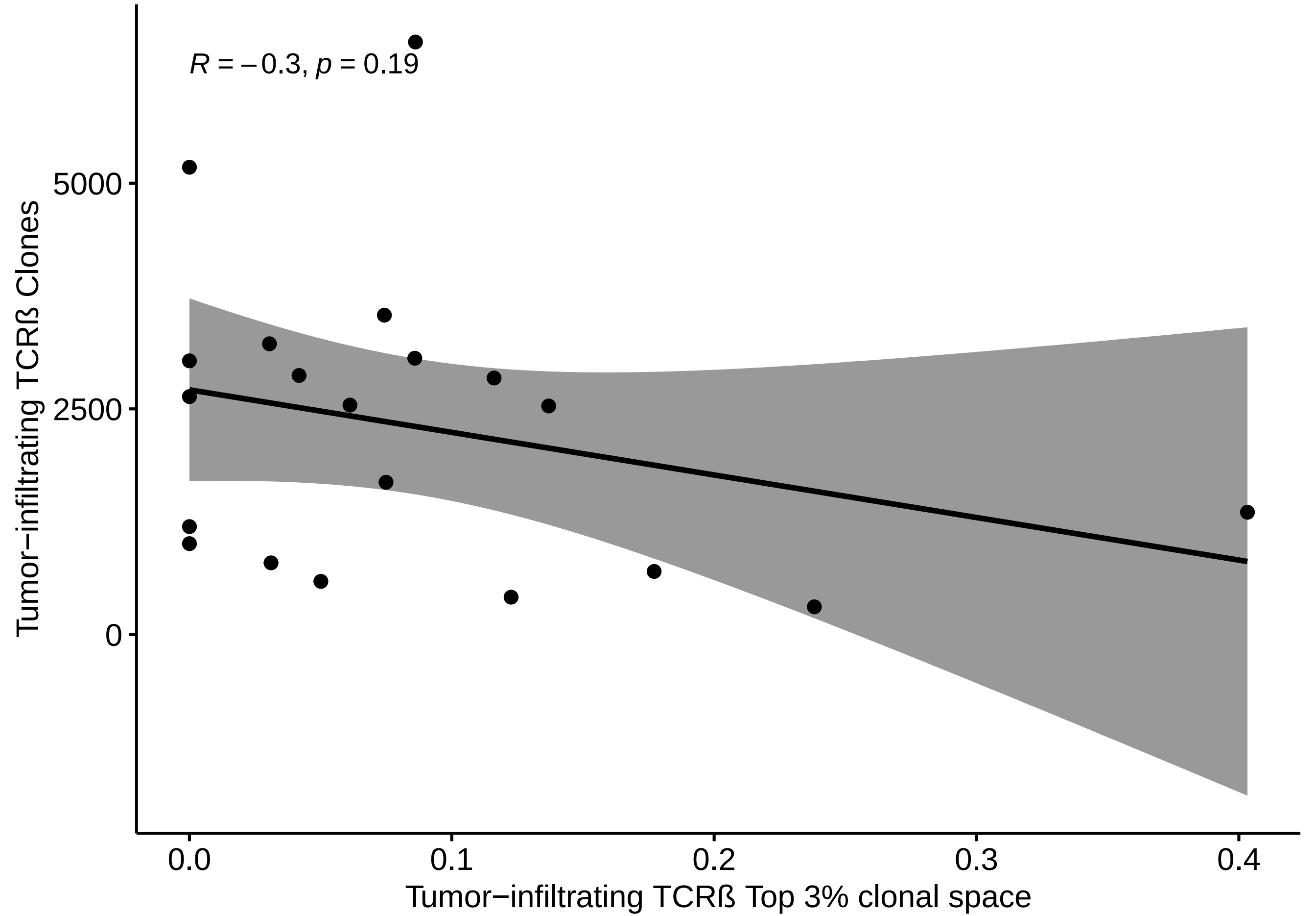
R = −0.3, p = 0.19
Tumor−infiltrating TCRß Clones
5000
2500
0
0.0
0.1
0.2
0.3
0.4
Tumor−infiltrating TCRß Top 3% clonal space

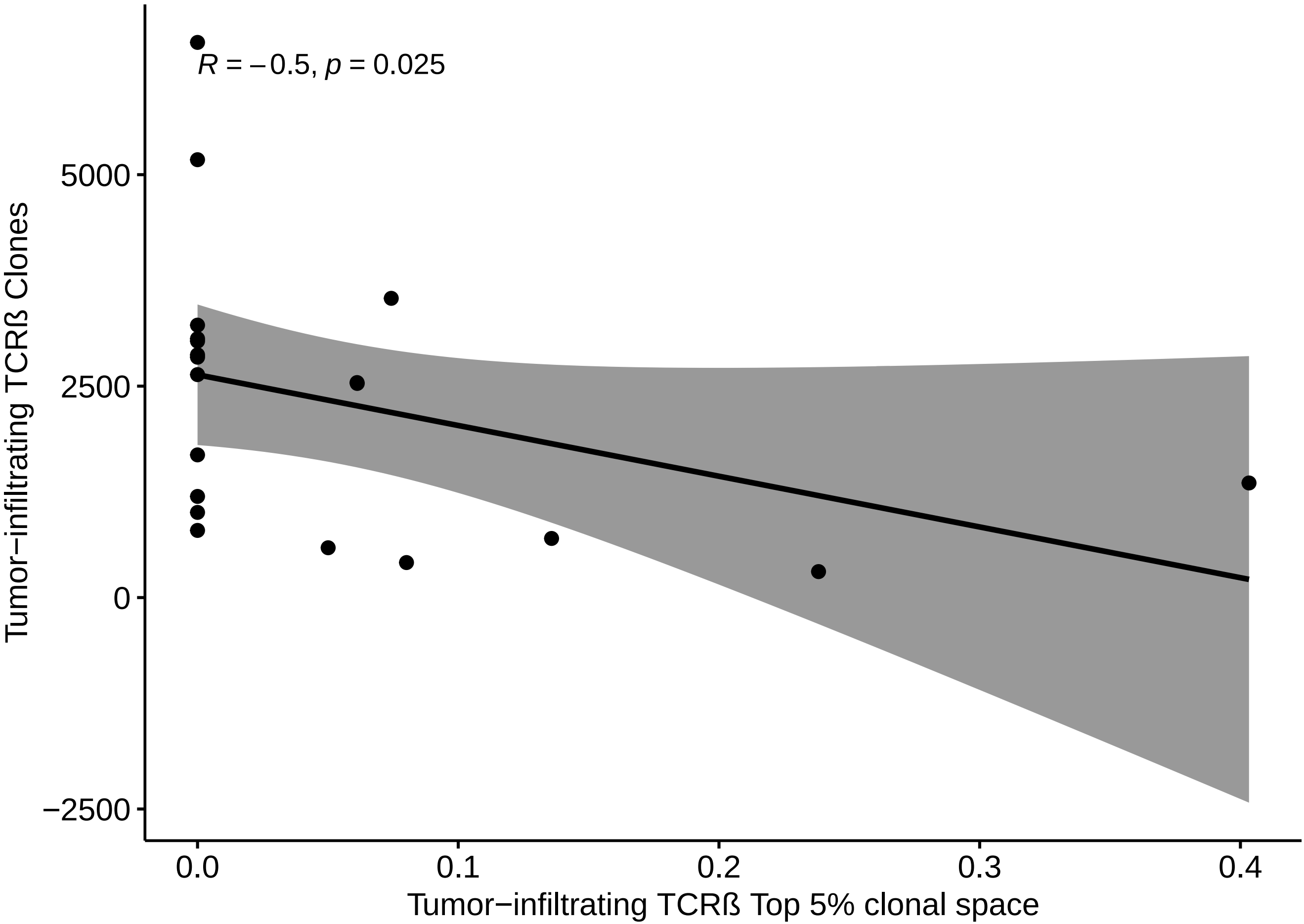
R = −0.5, p = 0.025
Tumor−infiltrating TCRß Clones
5000
2500
0
−2500
0.0
0.1
0.2
0.3
0.4
Tumor−infiltrating TCRß Top 5% clonal space

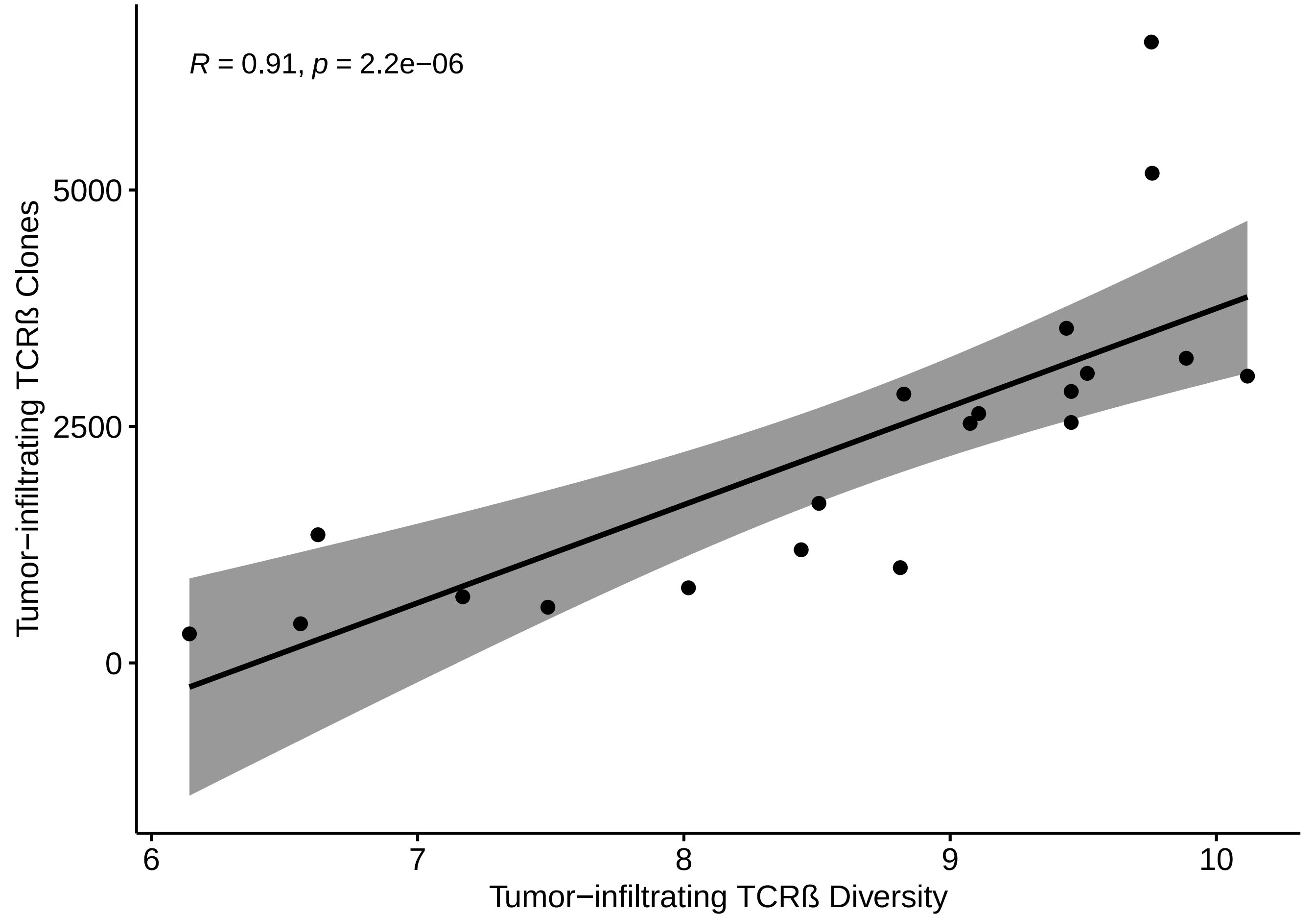

R = 0.91, p = 2.2e−06
Tumor−infiltrating TCRß Clones
5000
2500
0
6
7
8
9
10
Tumor−infiltrating TCRß Diversity

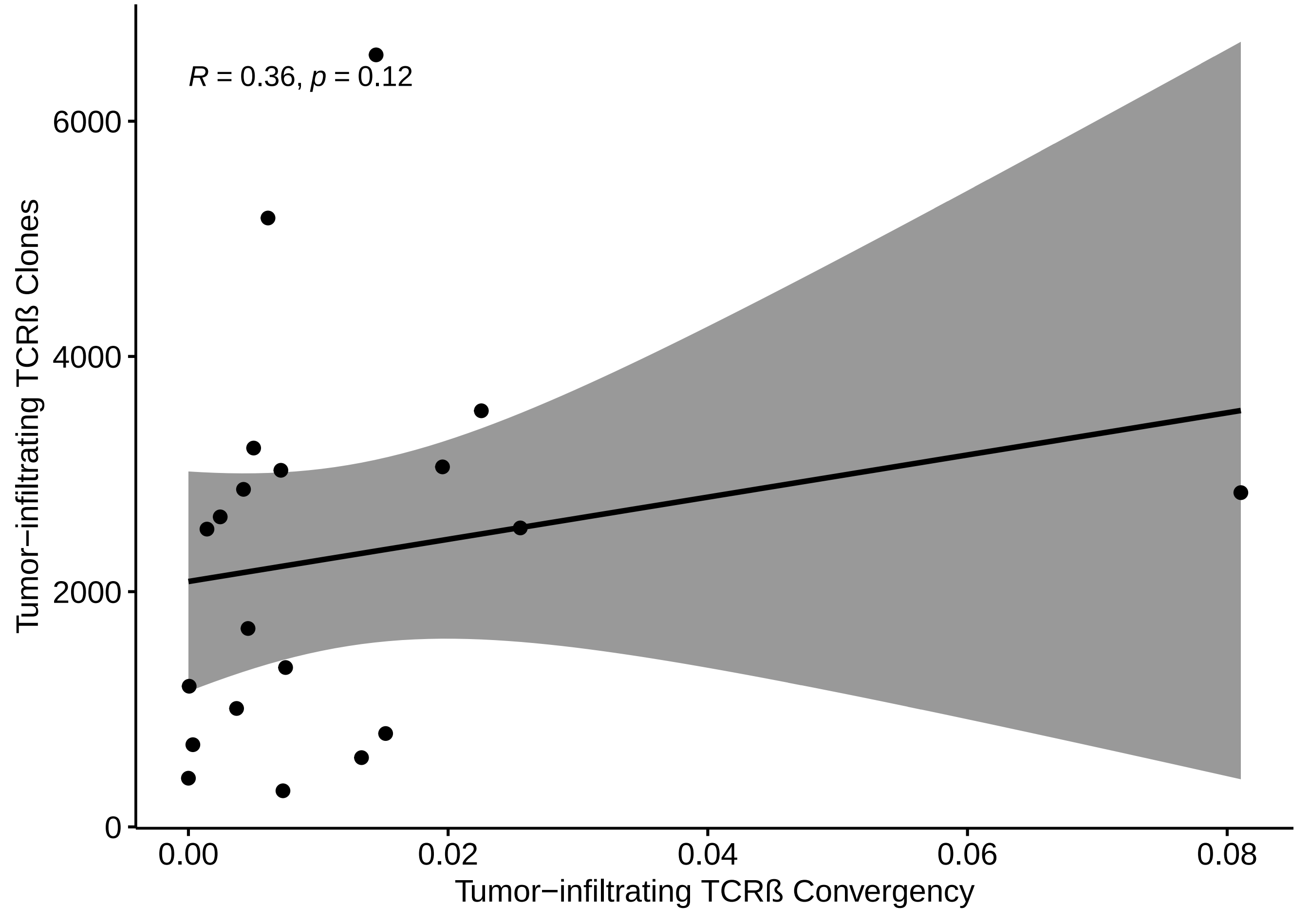

R = 0.36, p = 0.12
Tumor−infiltrating TCRß Clones
Tumor−infiltrating TCRß Convergency
0
2000
4000
6000
0.00
0.02
0.04
0.06
0.08

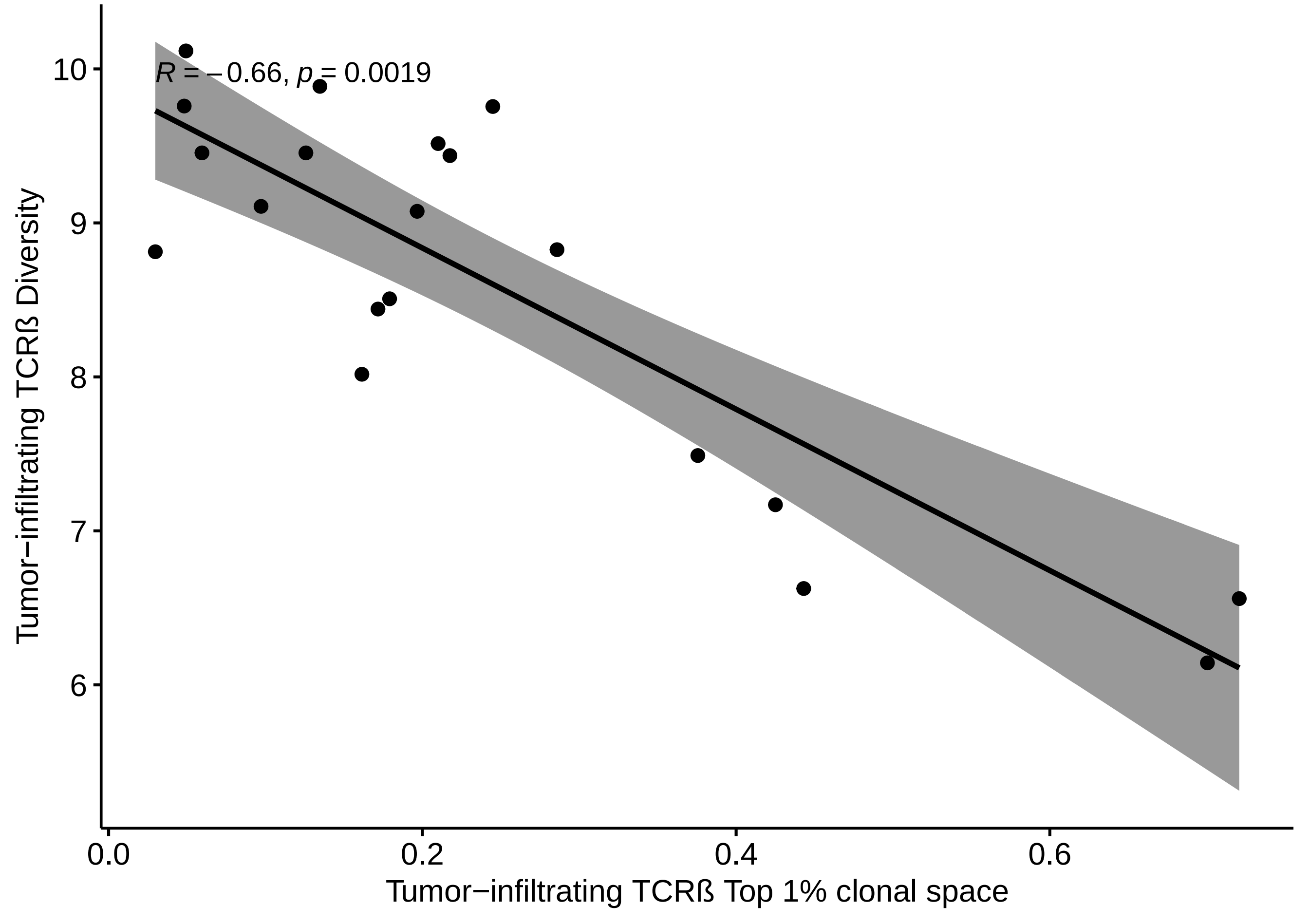
R = −0.66, p = 0.0019
Tumor−infiltrating TCRß Diversity
10
9
8
7
6
0.0
0.2
0.4
0.6
Tumor−infiltrating TCRß Top 1% clonal space

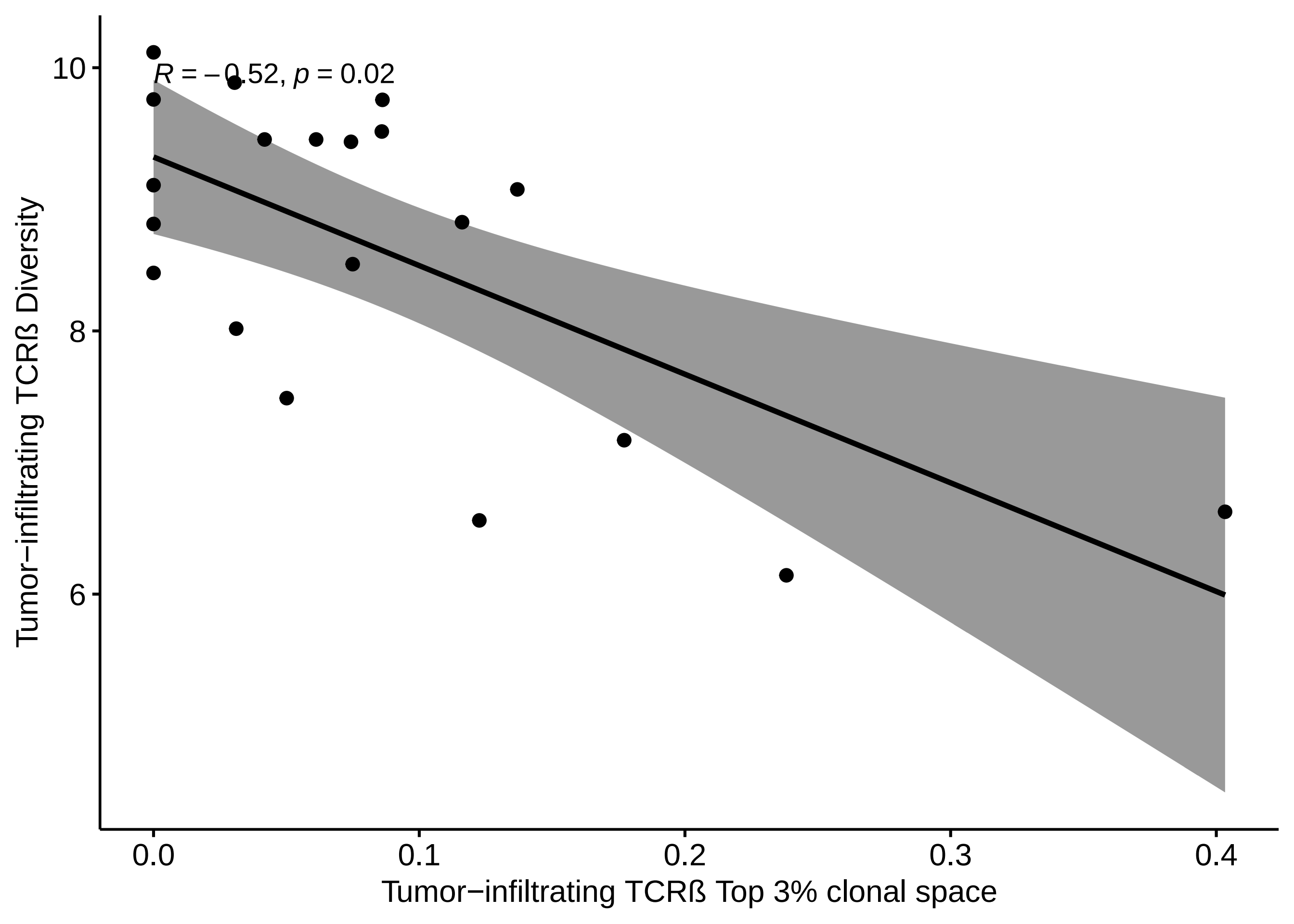
R = −0.52, p = 0.02
10
8
6
Tumor−infiltrating TCRß Diversity
0.0
0.1
0.2
0.3
0.4
Tumor−infiltrating TCRß Top 3% clonal space

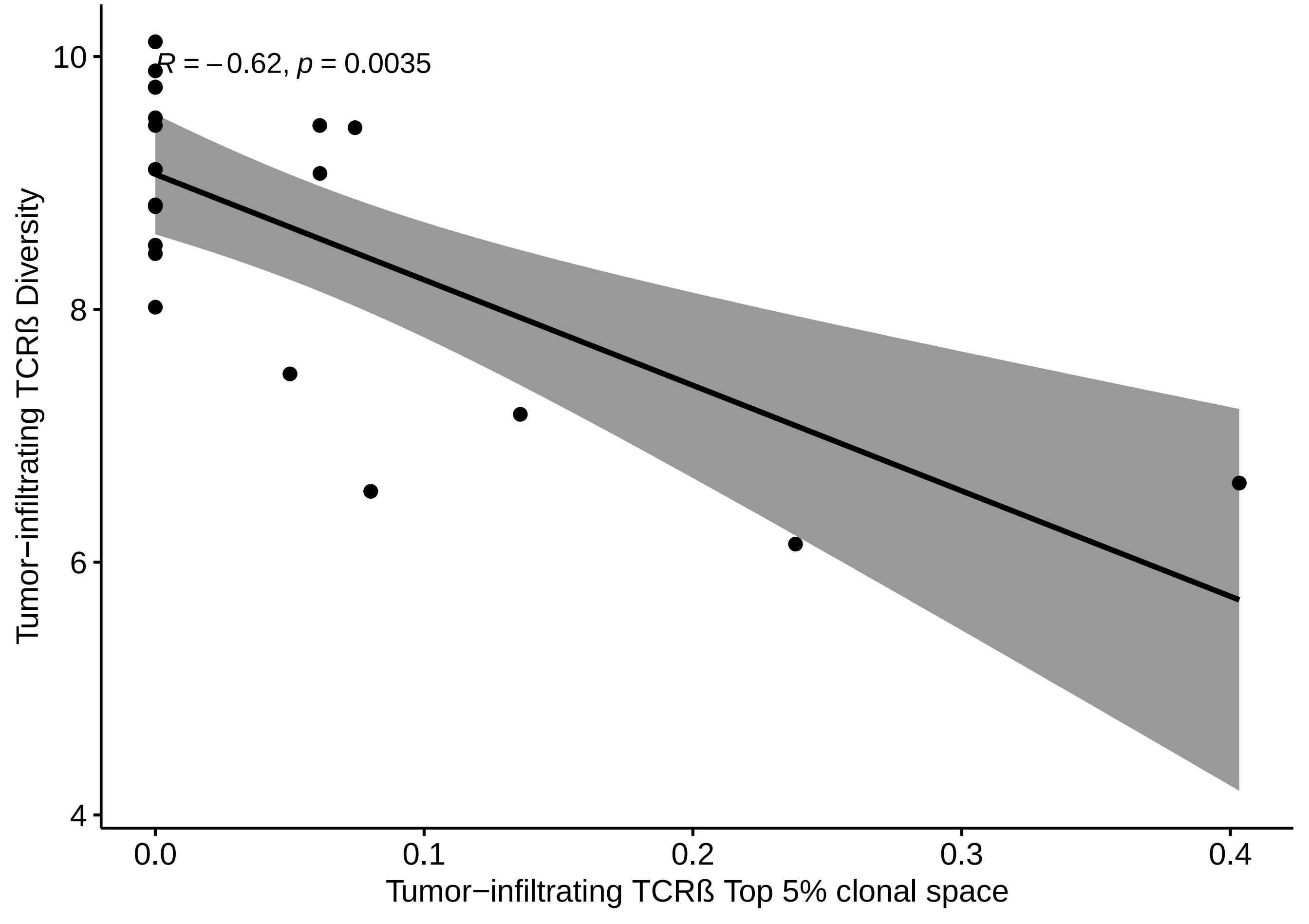

R = −0.62, p = 0.0035
10
8
6
4
0.0
0.1
0.2
0.3
0.4
Tumor−infiltrating TCRß Diversity
Tumor−infiltrating TCRß Top 5% clonal space

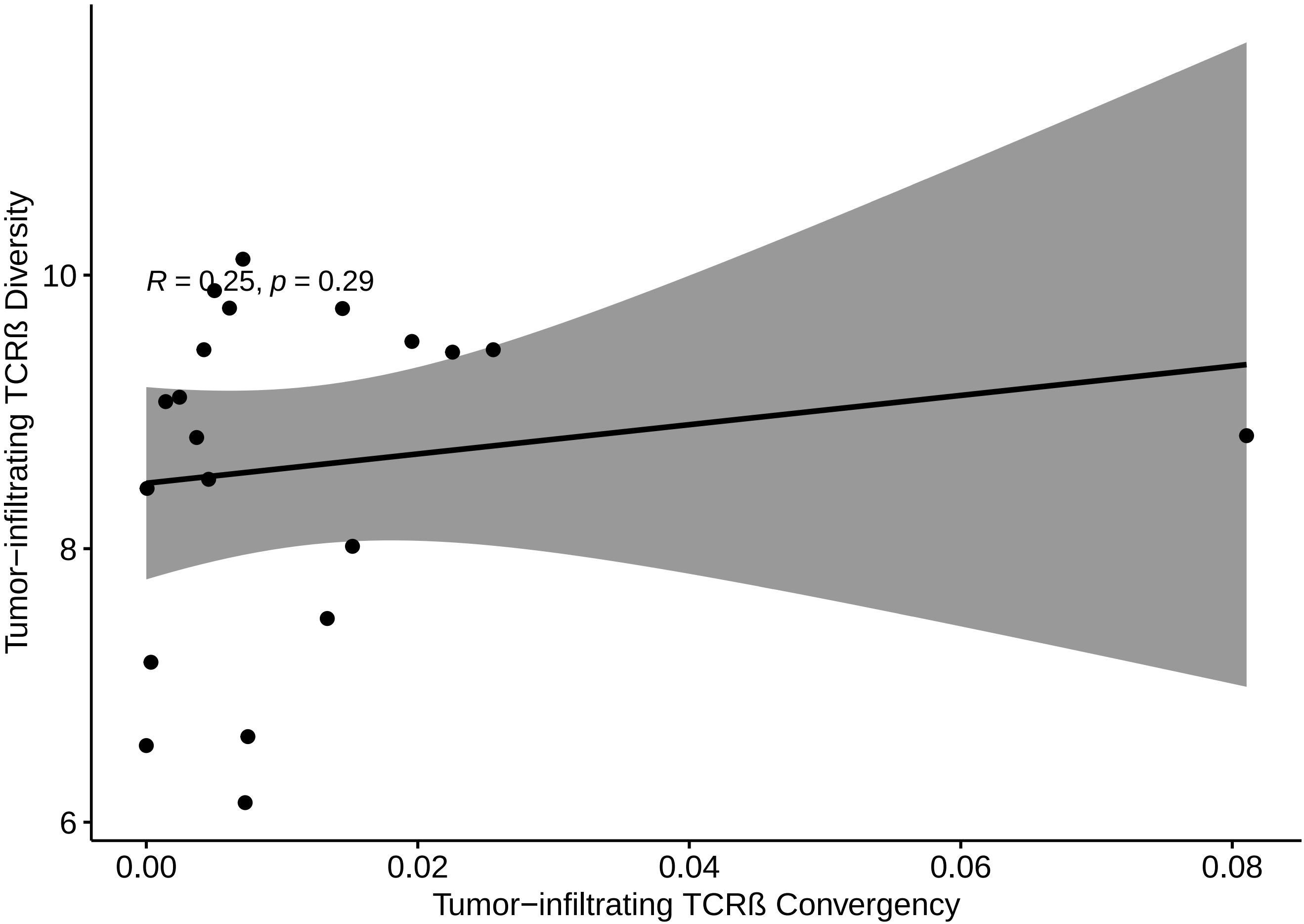

R = 0.25, p = 0.29
Tumor−infiltrating TCRß Diversity
10
8
6
0.00
0.02
0.04
0.06
0.08
Tumor−infiltrating TCRß Convergency

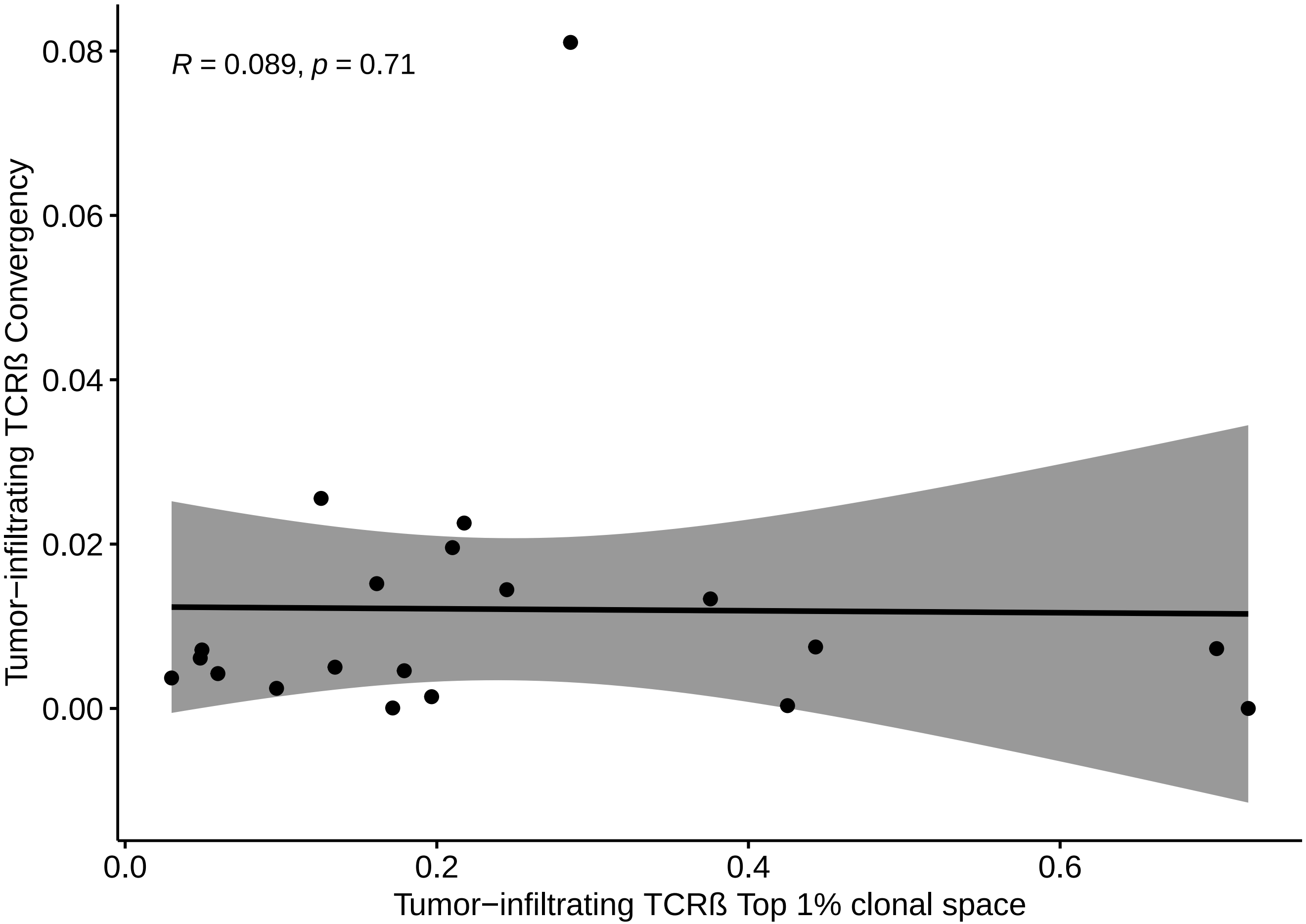

R = 0.089, p = 0.71
0.08
0.06
0.04
0.02
0.00
Tumor−infiltrating TCRß Convergency
0.0
0.2
0.4
0.6
Tumor−infiltrating TCRß Top 1% clonal space

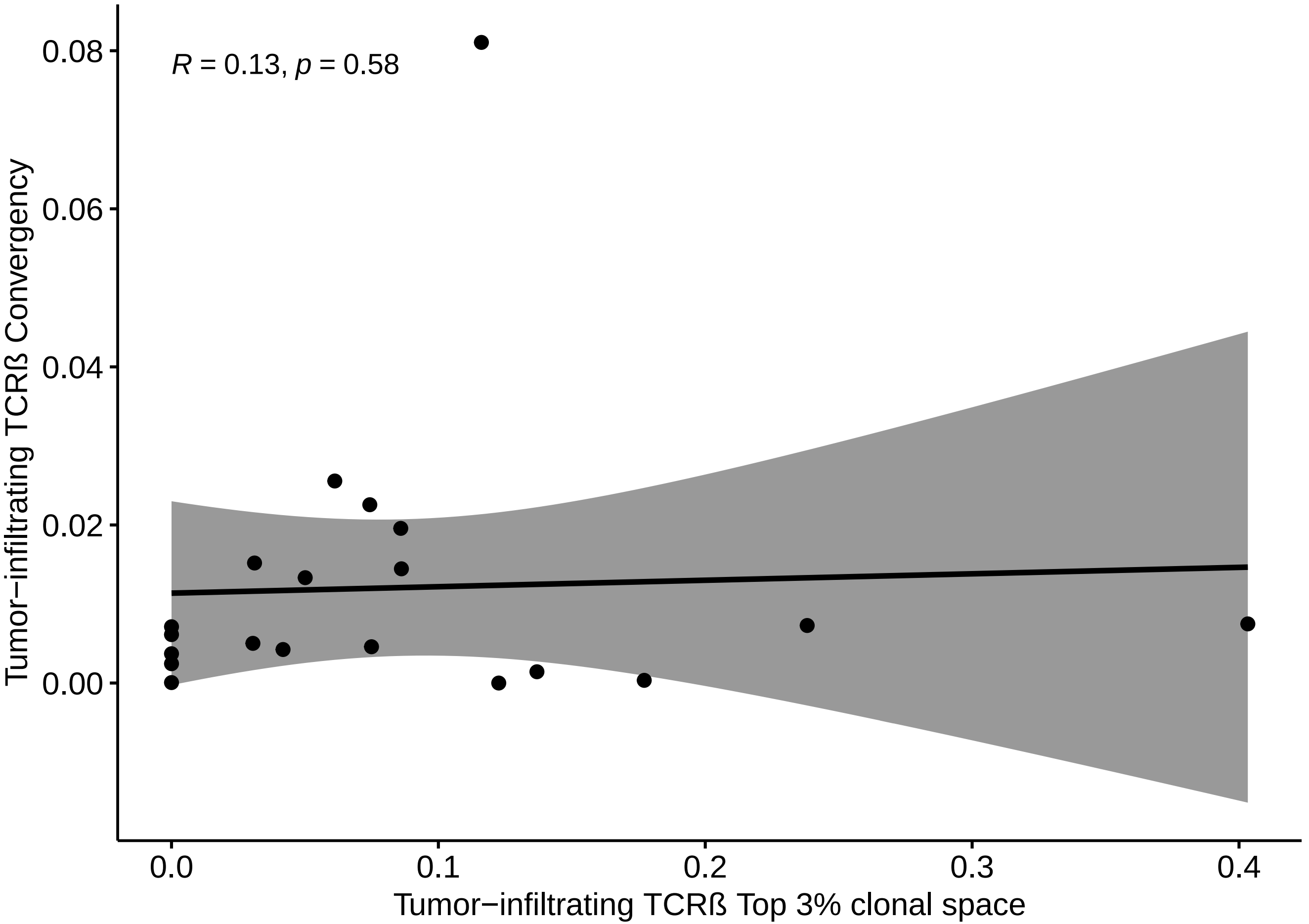
R = 0.13, p = 0.58
Tumor−infiltrating TCRß Convergency
0.08
0.06
0.04
0.02
0.00
0.0
0.1
0.2
0.3
0.4
Tumor−infiltrating TCRß Top 3% clonal space

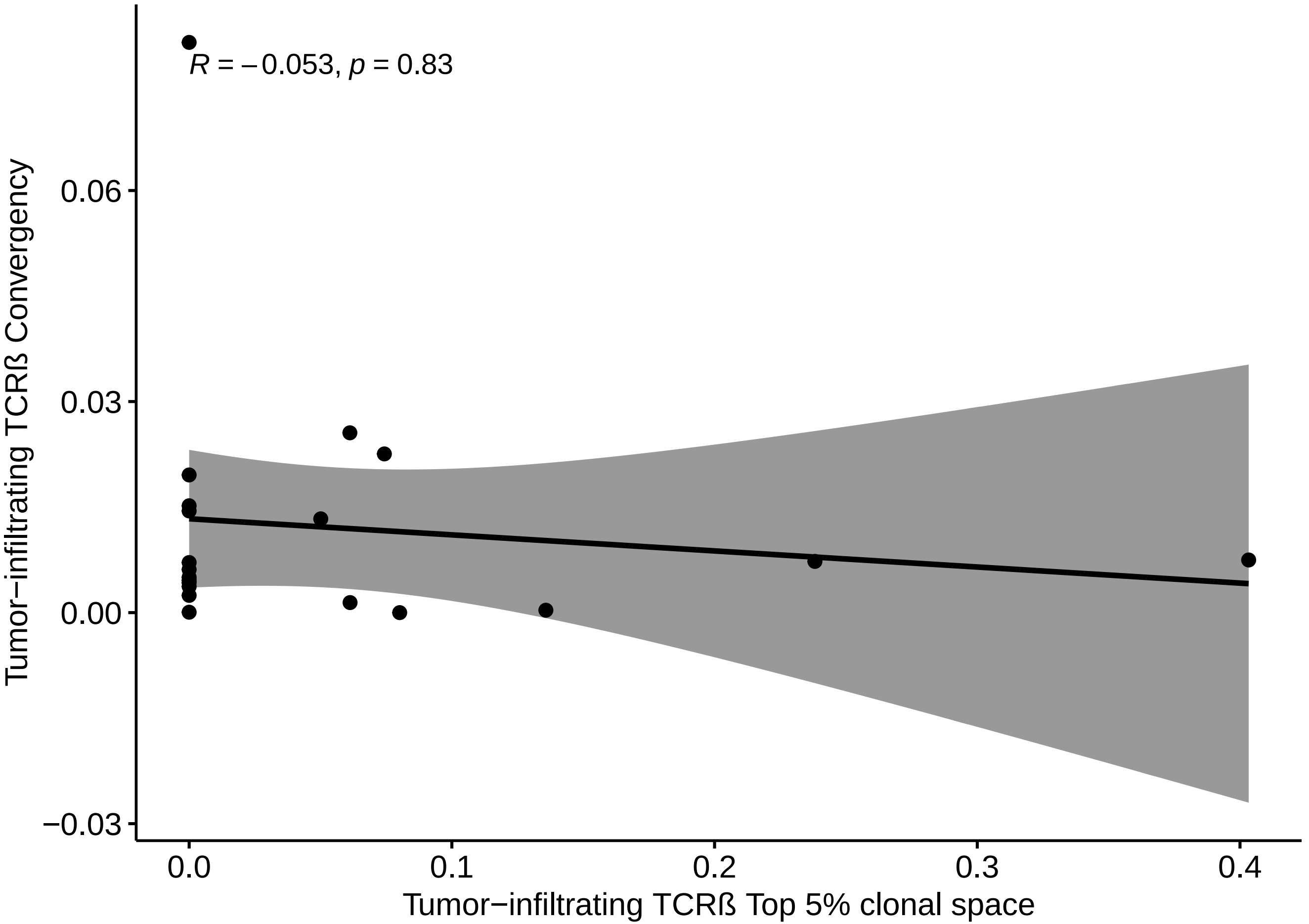
R = −0.053, p = 0.83
0.06
0.03
0.00
−0.03
0.0
0.1
0.2
0.3
0.4
Tumor−infiltrating TCRß Convergency
Tumor−infiltrating TCRß Top 5% clonal space
