## Supplementary File S3 for "Baseline intratumoral TCRβ and TCRγ repertoires predict response to immune checkpoint inhibitors in advanced renal cell carcinoma"

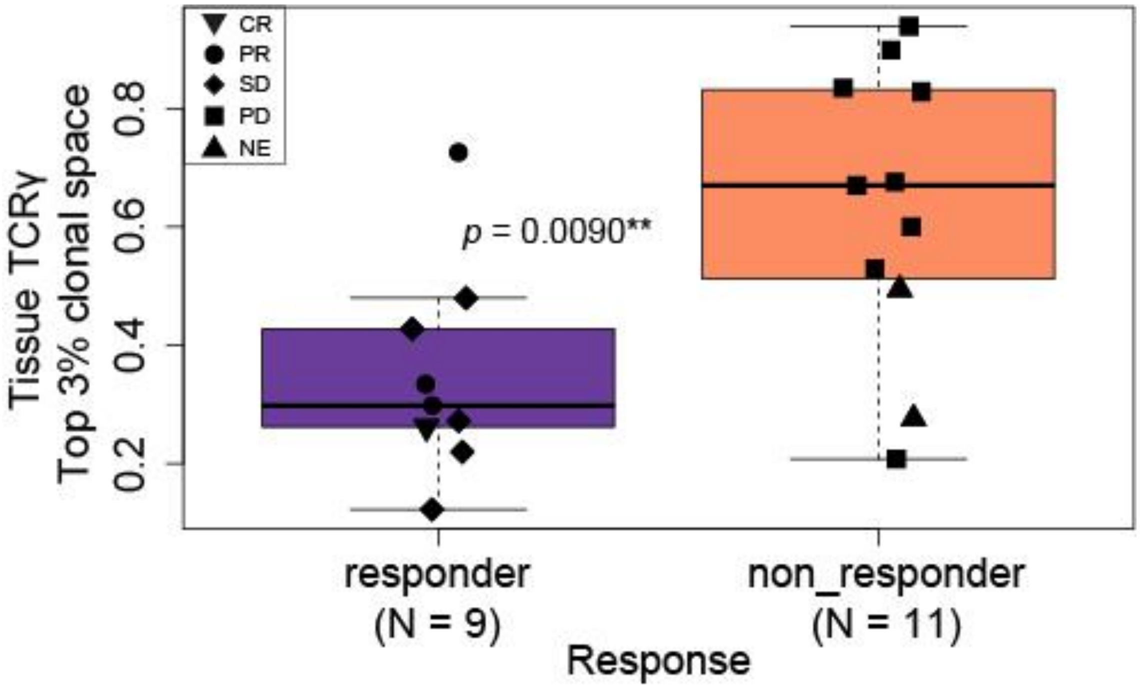


Figure 1: Responders have statistically significant lower tumor-infiltrating TCRγ top 3% clonal space than non-responders (*p* = 0.0090). *P* was obtained using Student's t test.

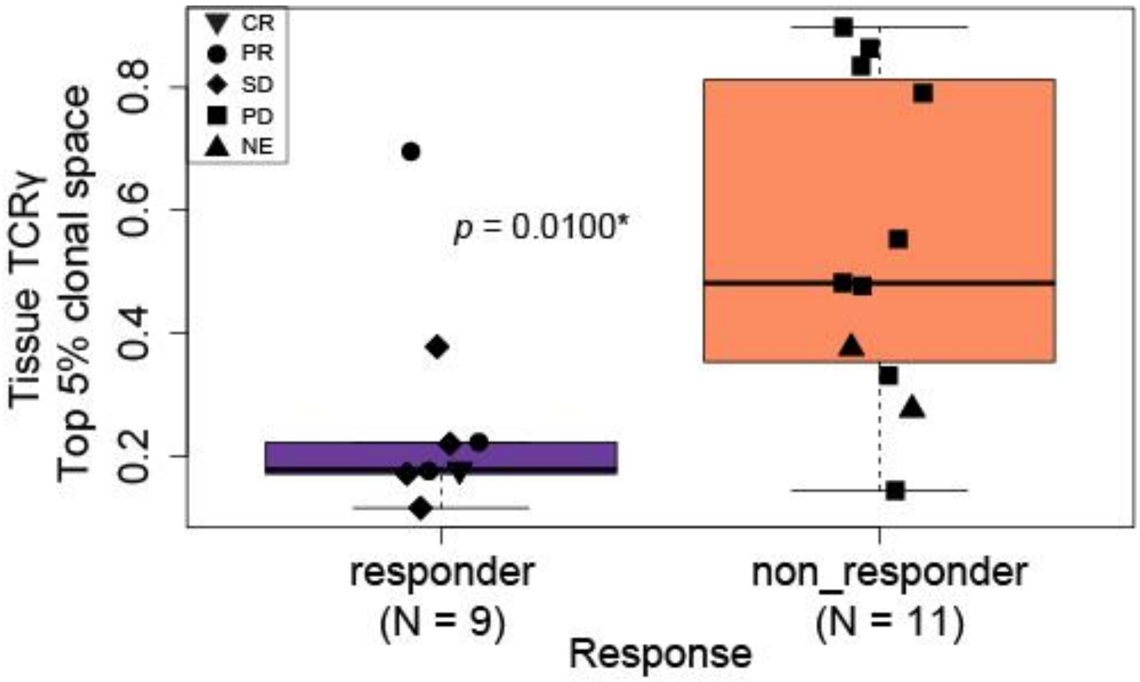


Figure 2: Responders have statistically significant lower tumor-infiltrating TCRγ top 5% clonal space than non-responders (*p* = 0.0100). *P* was obtained using Mann-Whitney test.

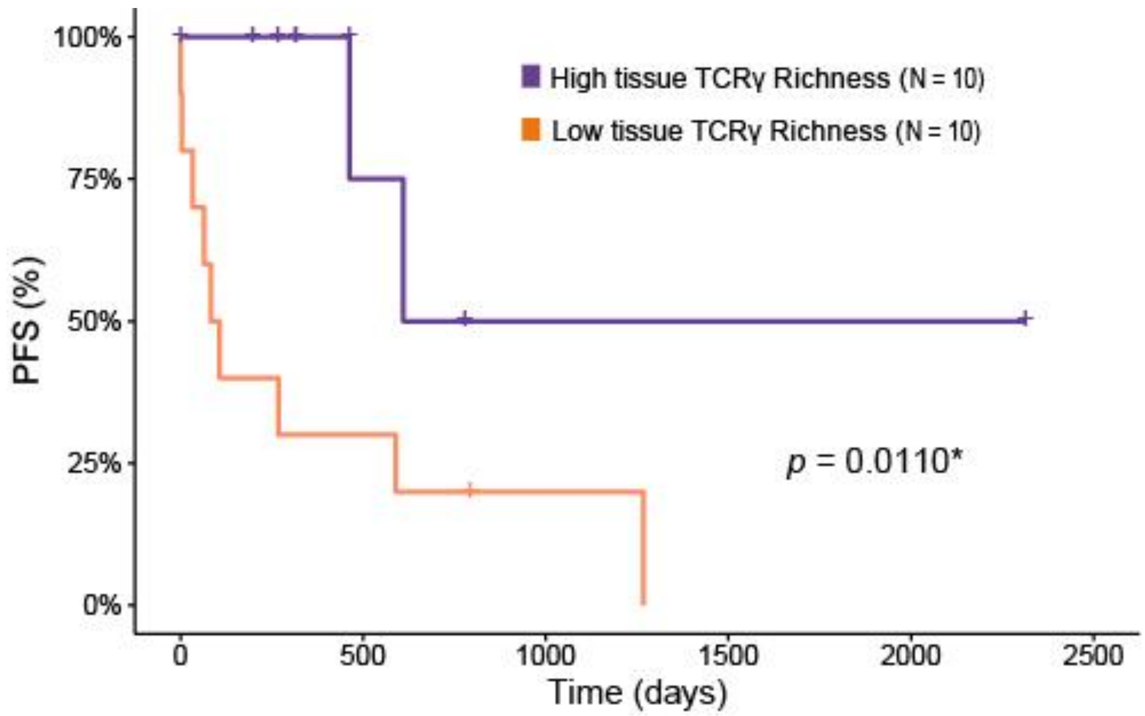


Figure 3: Patients with high tumor-infiltrating TCRγ richness have statistically significant longer PFS than patients with low tumor-infiltrating TCRγ richness (*p* = 0.0110). *P* was obtained using Log-Rank test.

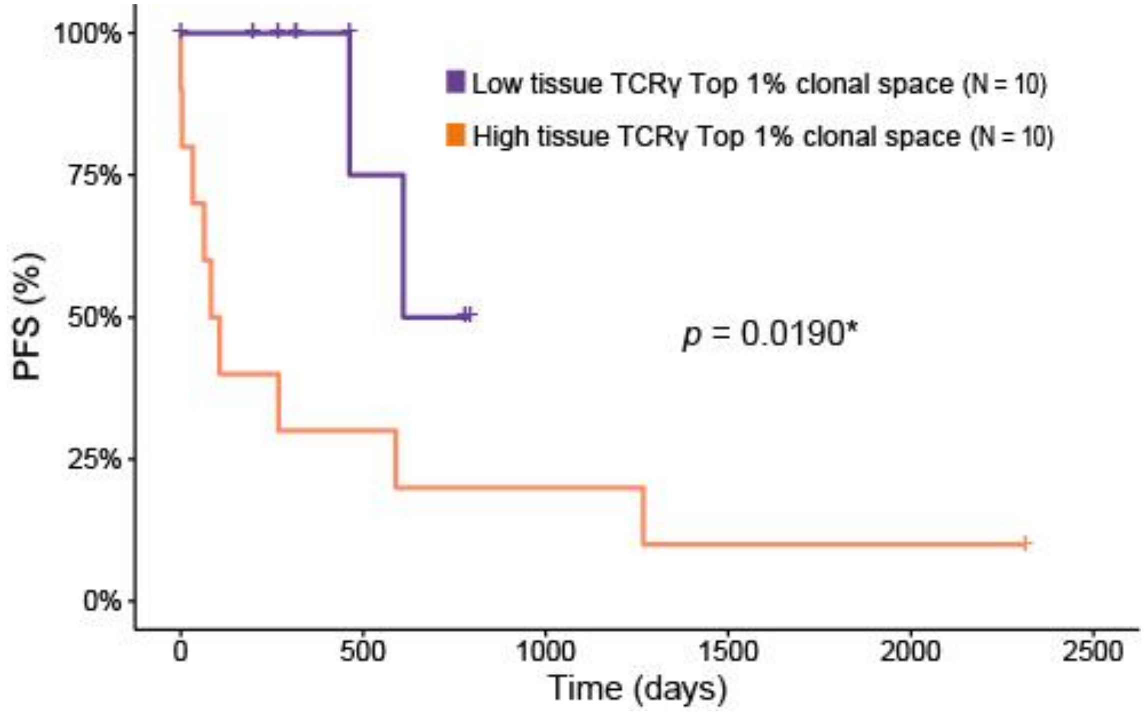


Figure 4: Patients with low tumor-infiltrating TCRγ top 1% clonal space have statistically significant longer PFS than patients with high tumor-infiltrating TCRγ top 1% clonal space (*p* = 0.0190). *P* was obtained using Log-Rank test.

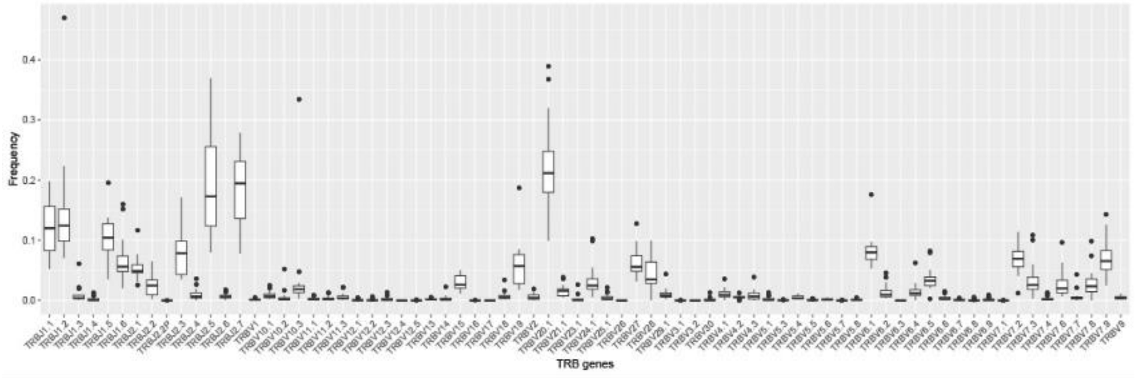


Figure 5: Boxplot showing the distribution of TRBV and TRBJ gene frequencies across the cohort.

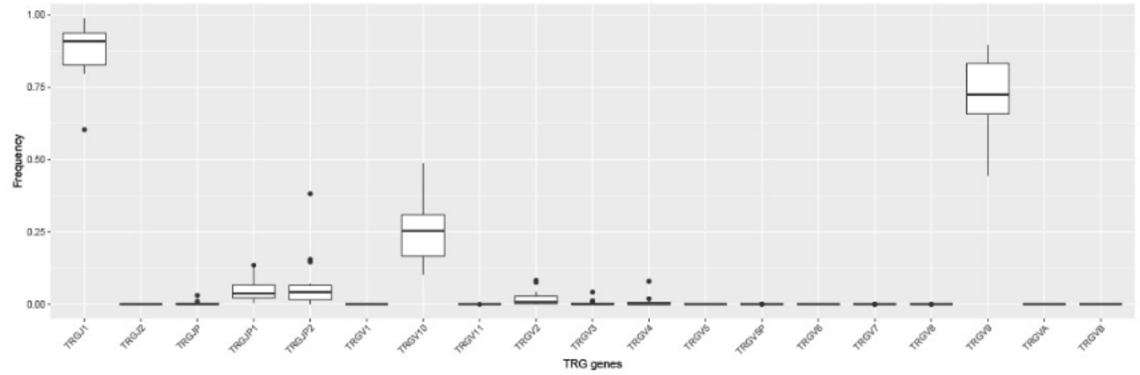


Figure 6: Boxplot showing the distribution of TRGV and TRGJ gene frequencies across the cohort.

Table 1: Tumor-infiltrating TCRβ TRBV and TRBJ genes frequency in responder (R) compared to non-responders (NR).

| Variable | Median | | *p*-value |
|---|---|---|---|
| | Responder (N = 9) | Non-responder (N = 11) | |
| ***TRBV2*** | 0.00832 | 0.00210 | 0.0200* |
| ***TRBV7.1*** | 0.00000406 | 0.00 | 0.0460* |
| ***TRBV4.2*** | 0.00622 | 0.00475 | 0.0330* |
| ***TRBV9*** | 0.00581 | 0.0301 | 0.0330* |
| ***TRBV12.2*** | 0.000306 | 0.0000305 | 0.0330* |
| ***TRBV6.5*** | 0.0417 | 0.0257 | 0.0100* |
| ***TRBV5.7*** | 0.0000182 | 0.00 | 0.0050** |
| ***TRBV16*** | 0.0000219 | 0.00 | 0.0210* |
| ***TRBV18*** | 0.00617 | 0.00327 | 0.0200* |
| ***TRBV23.1*** | 0.0014 | 0.000204 | 0.0200* |
| ***TRBV30*** | 0.00165 | 0.000139 | 0.0030** |
| ***TRBJ1.3*** | 0.0093 | 0.00348 | 0.0120* |
| ***TRBJ2.2*** | 0.0331 | 0.0111 | 0.0310* |
| ***TRBJ2.4*** | 0.0118 | 0.00409 | 0.0250* |

*P* was obtained using Mann-Whitney or Student's t test. N: number of patients, *: statistically significance, **: strongly statistically significance.

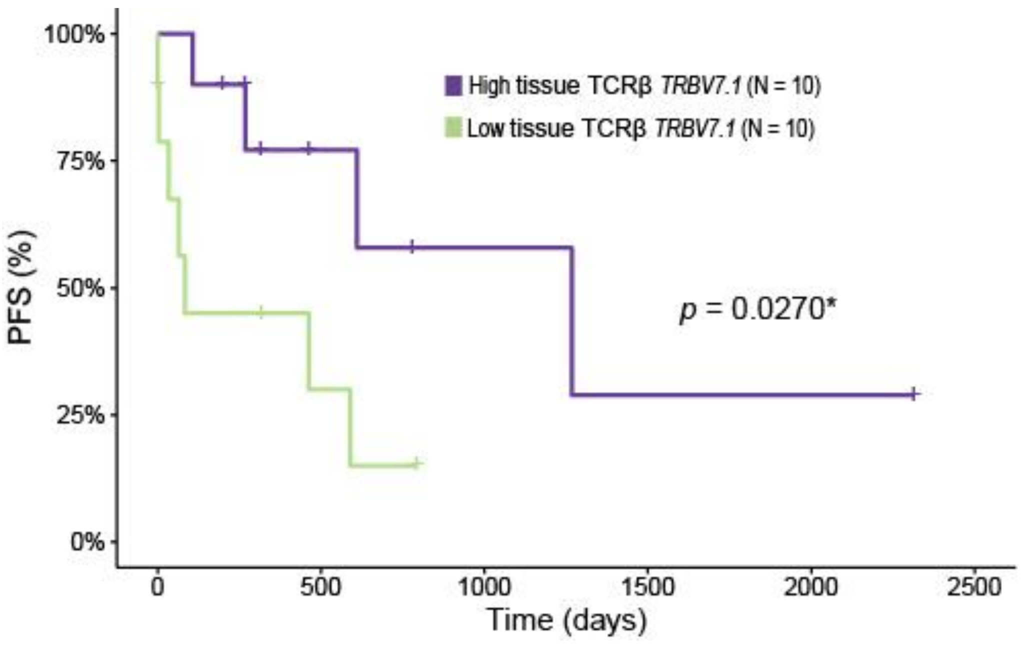


Figure 7: Patients with high tumor-infiltrating TCRβ *TRBV7.1* frequency have statistically significant longer PFS than patients with low tumor-infiltrating TCRβ *TRBV7.1* frequency ($p$ = 0.0270). $P$ was obtained using Log-Rank test.

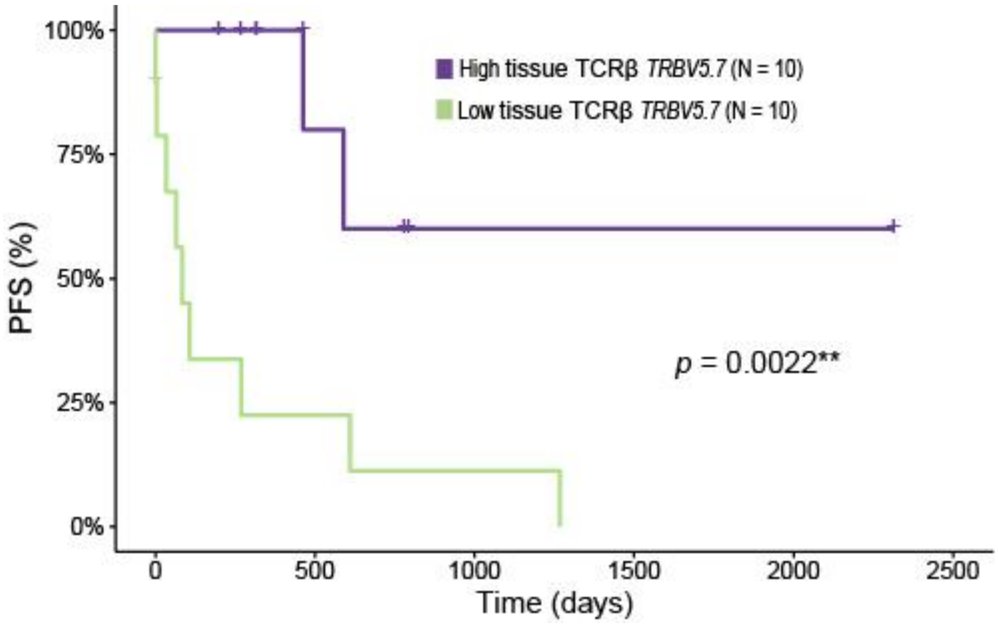


Figure 8: Patients with high tumor-infiltrating TCRβ *TRBV5.7* frequency have statistically significant longer PFS than patients with low tumor-infiltrating TCRβ *TRBV5.7* frequency ($p$ = 0.0022). *P* was obtained using Log-Rank test.

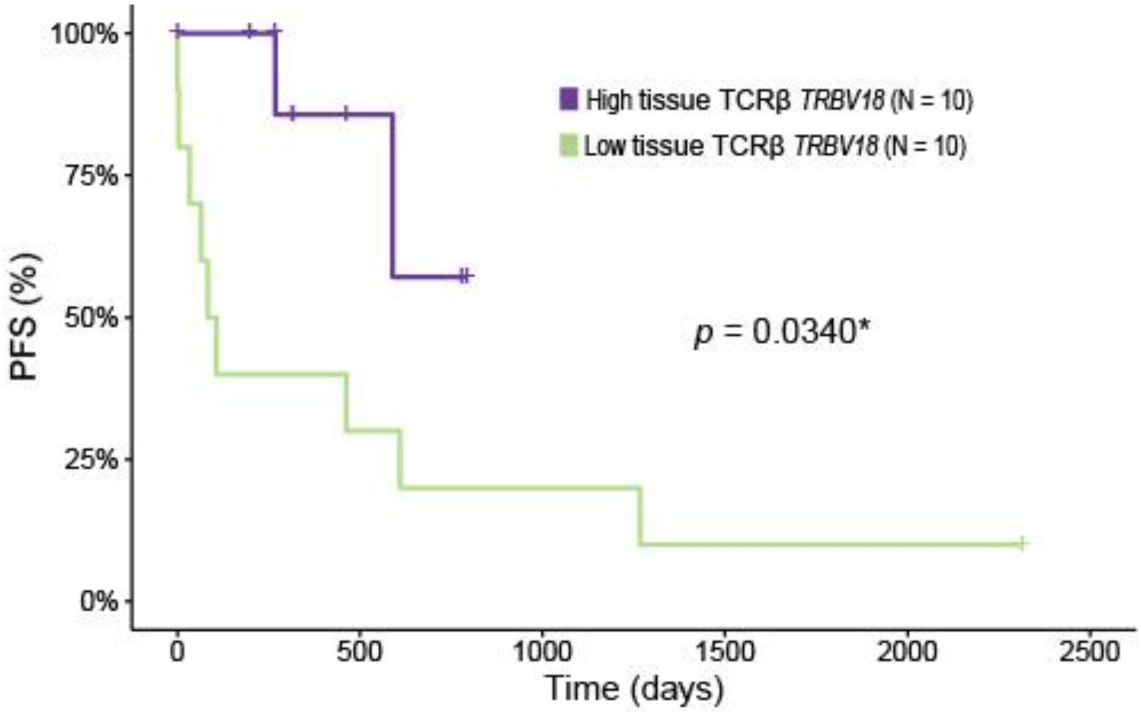


Figure 9: Patients with high tumor-infiltrating TCRβ *TRBV18* frequency have statistically significant longer PFS than patients with low tumor-infiltrating TCRβ *TRBV18* frequency ($p$ = 0.0340). *P* was obtained using Log-Rank test.
