## Supplementary File S4 for "Baseline intratumoral TCRβ and TCRγ repertoires predict response to immune checkpoint inhibitors in advanced renal cell carcinoma"

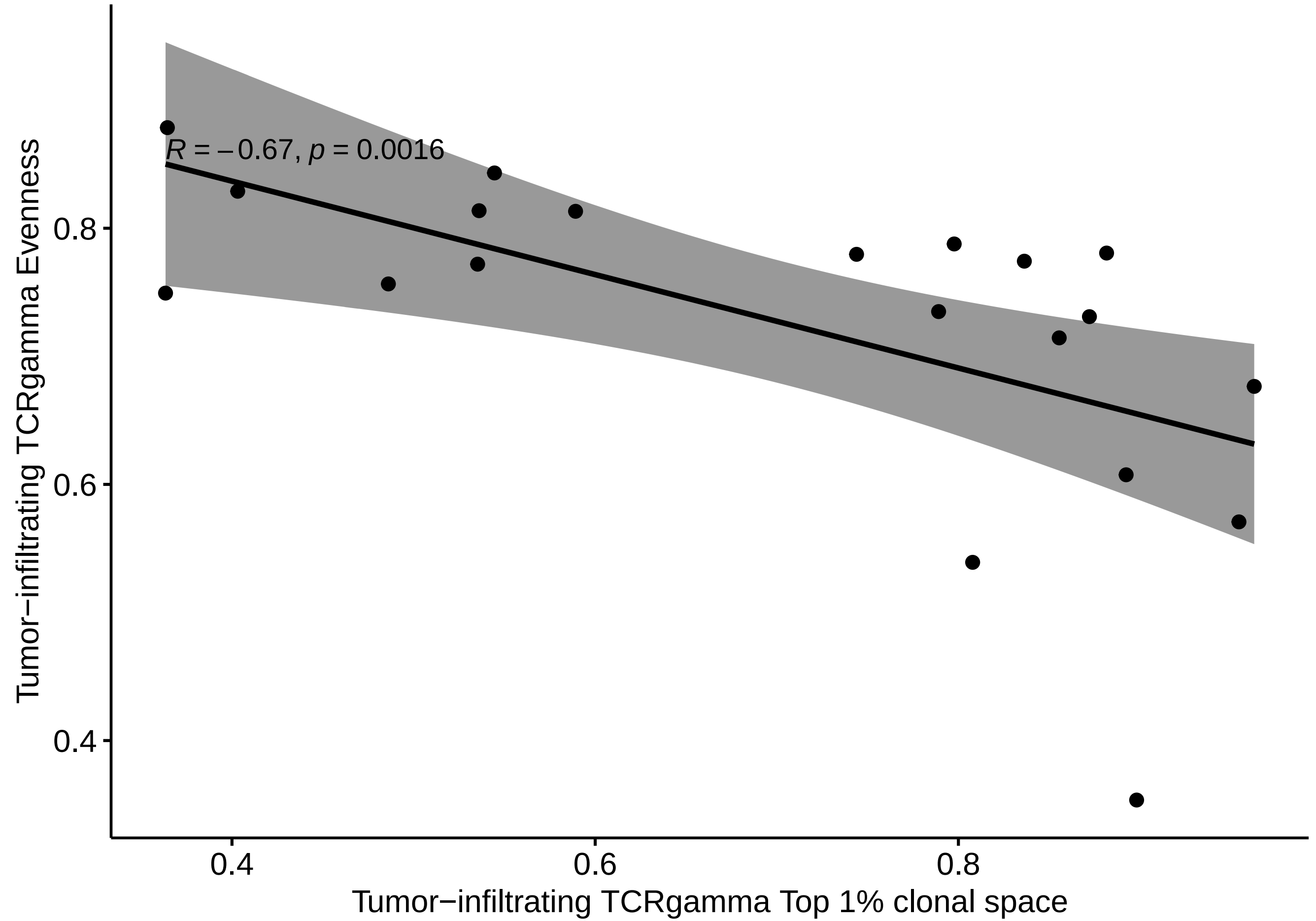

R = −0.67, p = 0.0016
0.8
0.6
0.4
Tumor−infiltrating TCRgamma Evenness
0.4
0.6
0.8
Tumor−infiltrating TCRgamma Top 1% clonal space

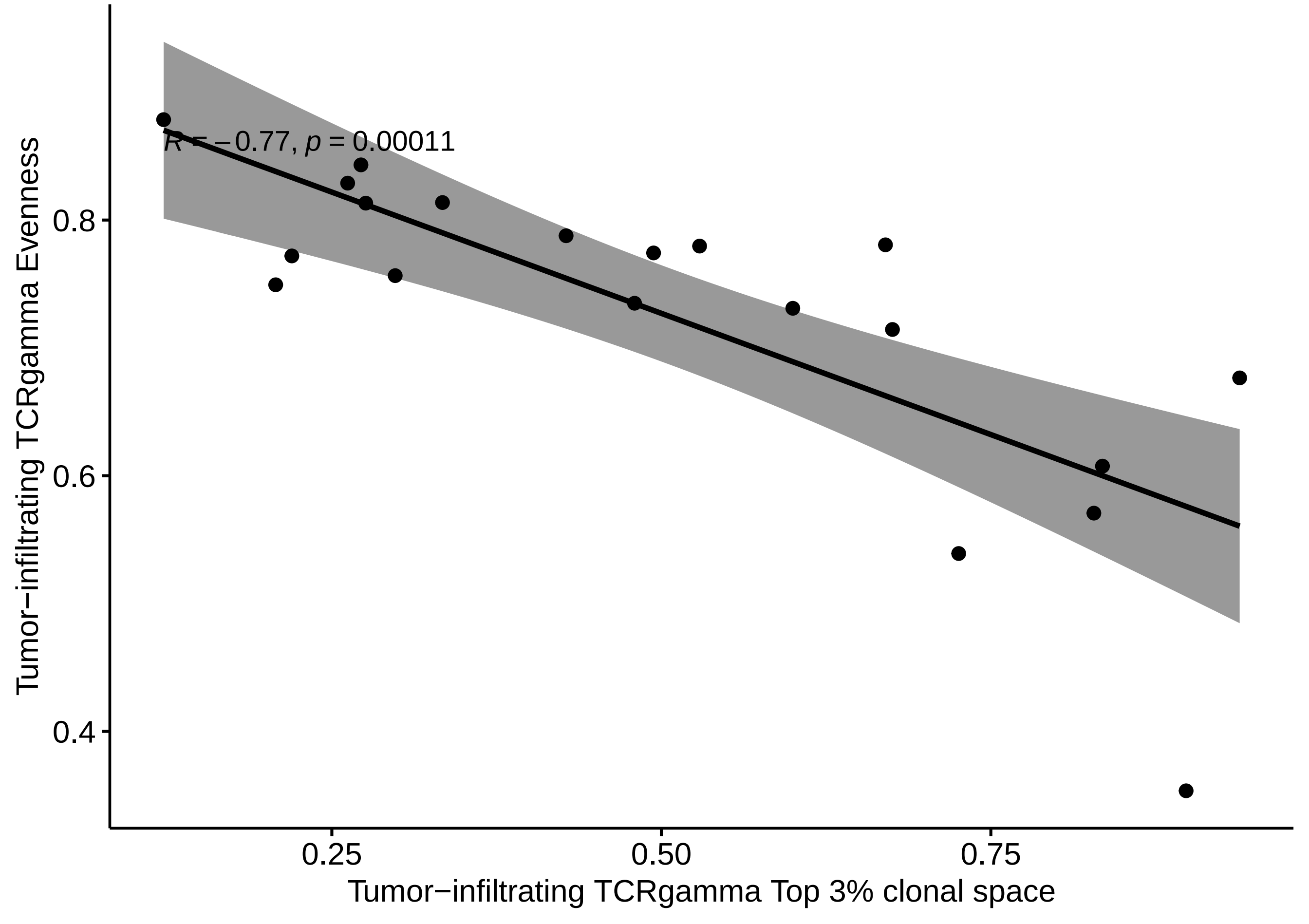

R = −0.77, p = 0.00011
Tumor−infiltrating TCRgamma Evenness
0.8
0.6
0.4
0.25
0.50
0.75
Tumor−infiltrating TCRgamma Top 3% clonal space

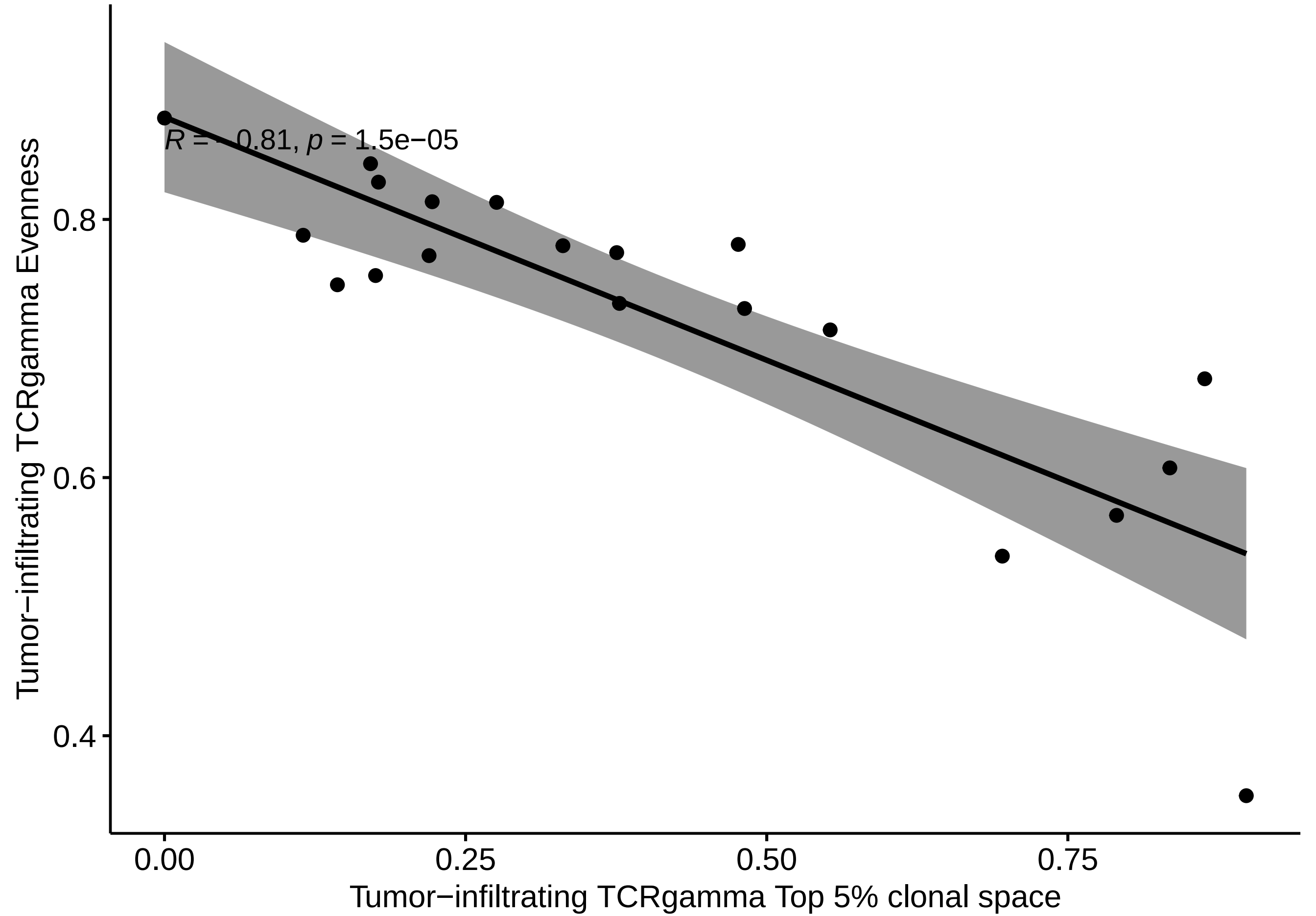
R = −0.81, p = 1.5e−05
Tumor−infiltrating TCRgamma Evenness
0.8
0.6
0.4
0.00
0.25
0.50
0.75
Tumor−infiltrating TCRgamma Top 5% clonal space

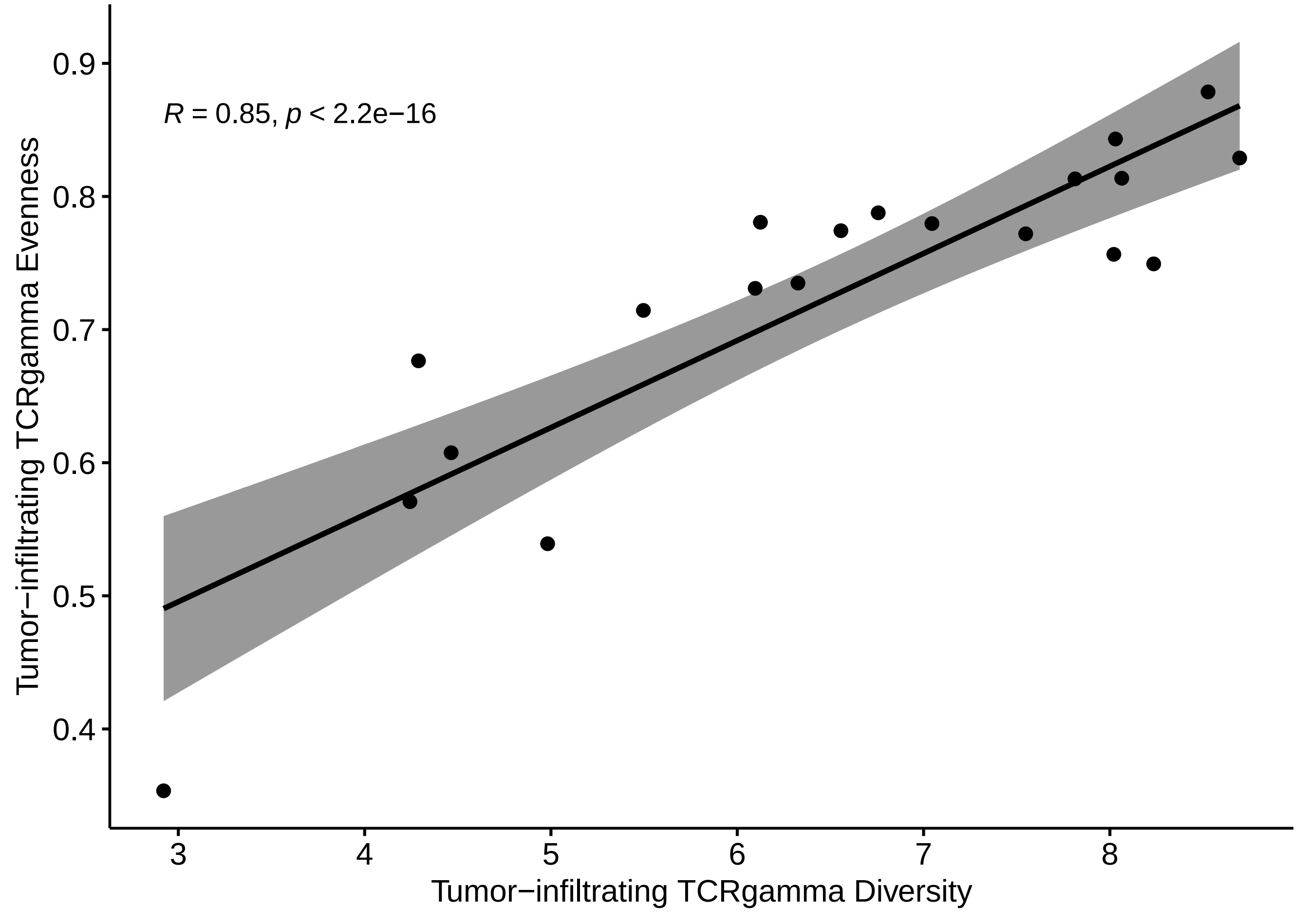
R = 0.85, p < 2.2e−16
Tumor−infiltrating TCRgamma Evenness
0.9
0.8
0.7
0.6
0.5
0.4
3
4
5
6
7
8
Tumor−infiltrating TCRgamma Diversity

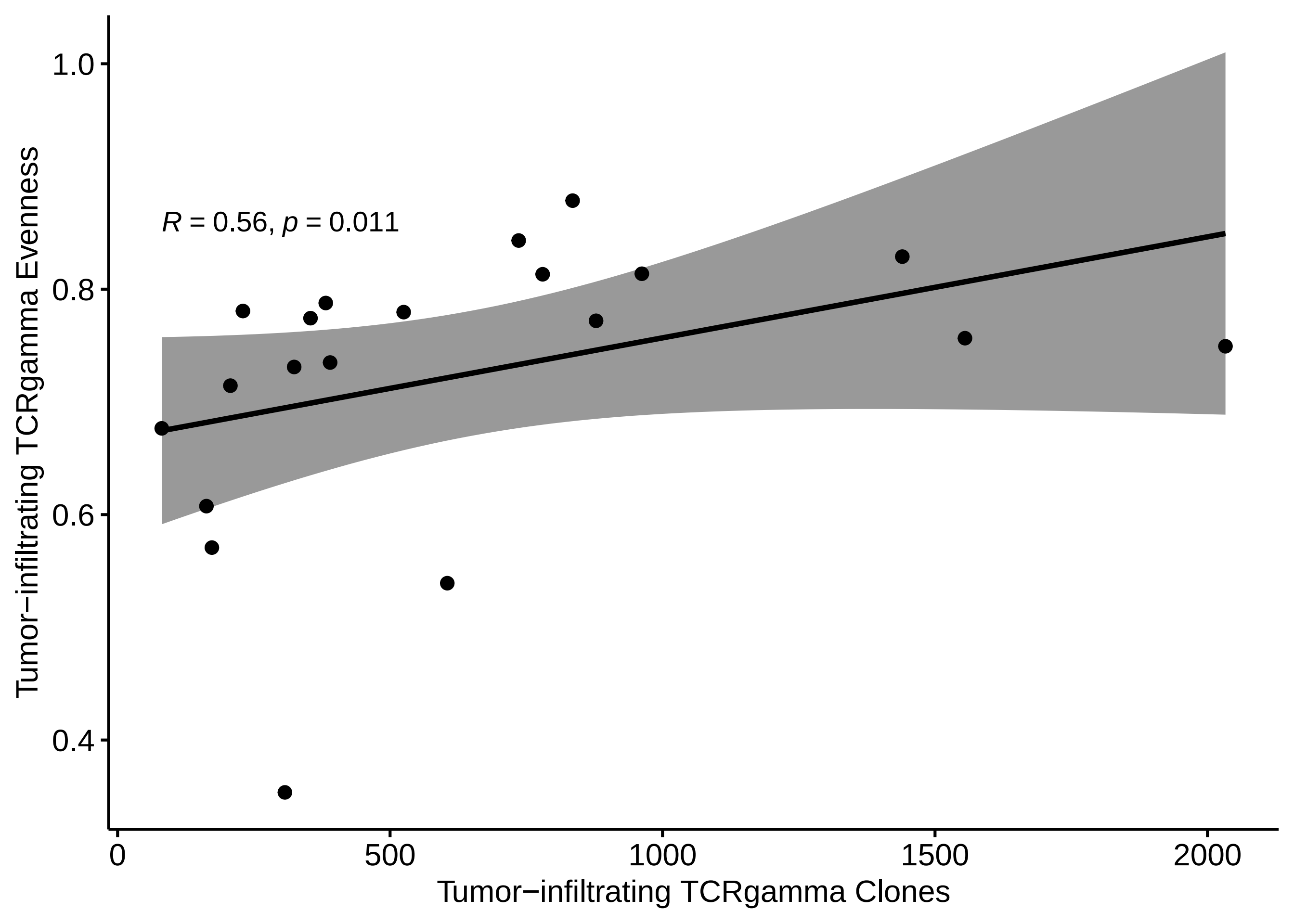

R = 0.56, p = 0.011
1.0
0.8
0.6
0.4
Tumor−infiltrating TCRgamma Evenness
0
500
1000
1500
2000
Tumor−infiltrating TCRgamma Clones

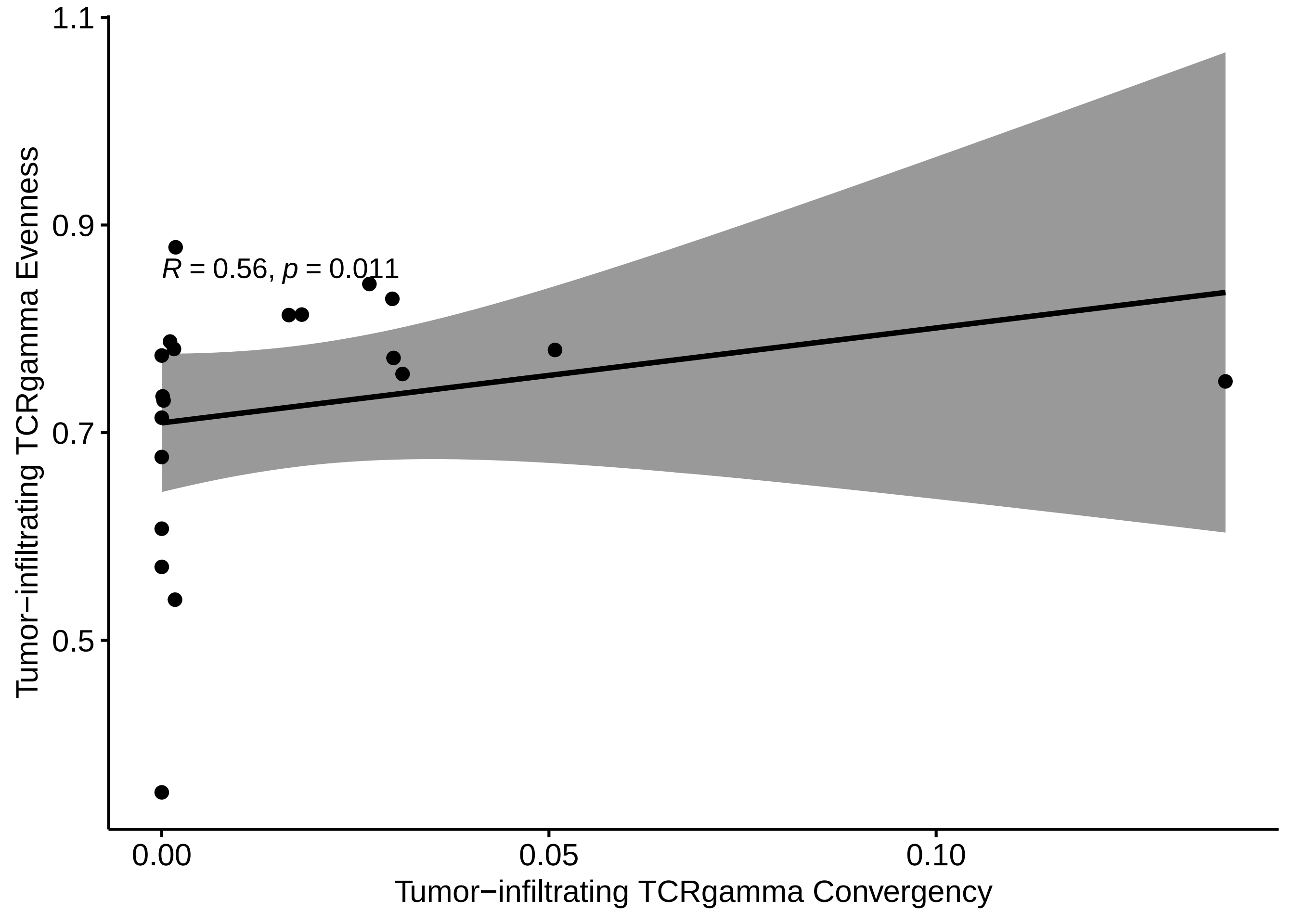

R = 0.56, p = 0.011
Tumor−infiltrating TCRgamma Evenness
1.1
0.9
0.7
0.5
0.00
0.05
0.10
Tumor−infiltrating TCRgamma Convergency

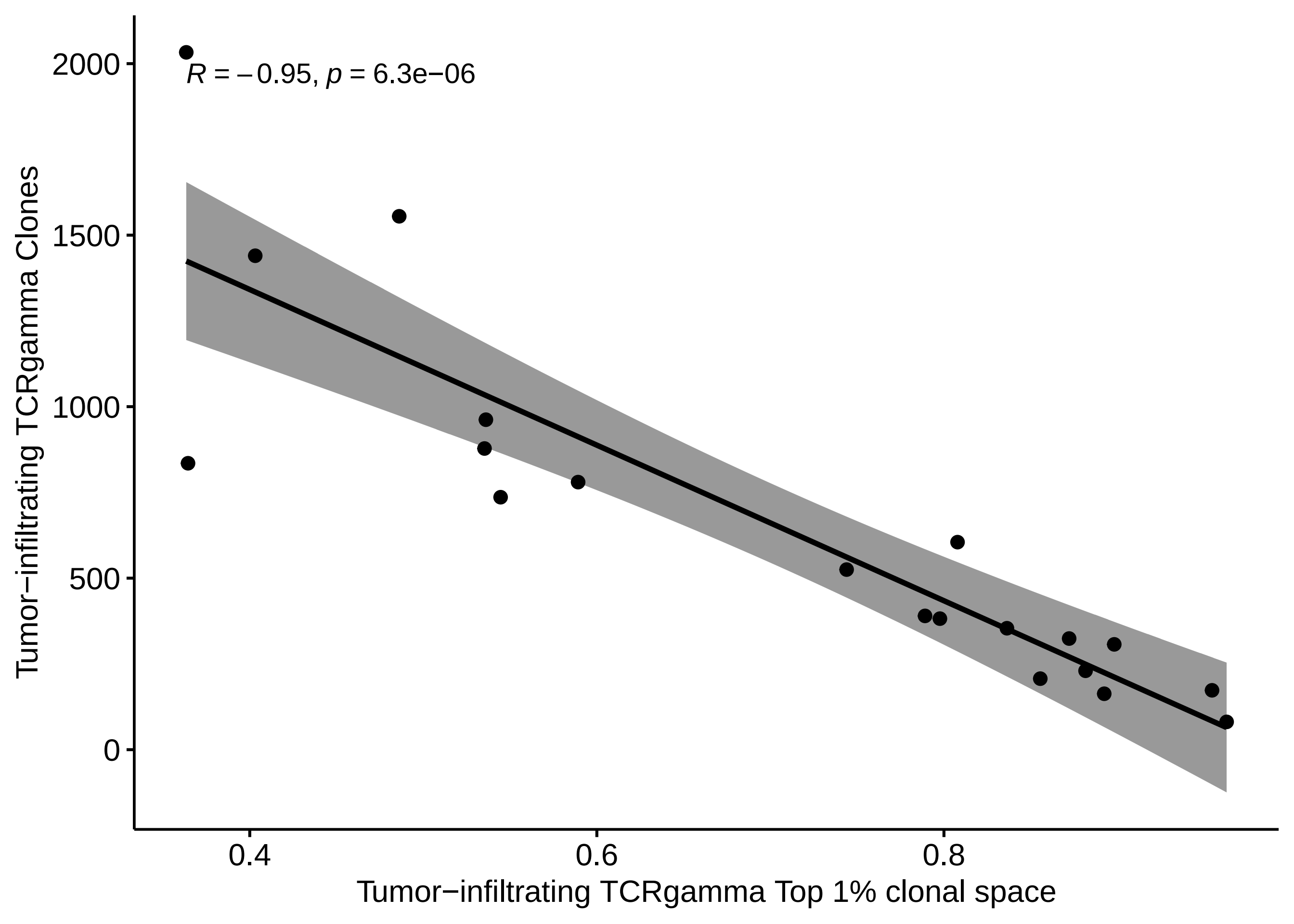
R = −0.95, p = 6.3e−06
Tumor−infiltrating TCRgamma Clones
2000
1500
1000
500
0
0.4
0.6
0.8
Tumor−infiltrating TCRgamma Top 1% clonal space

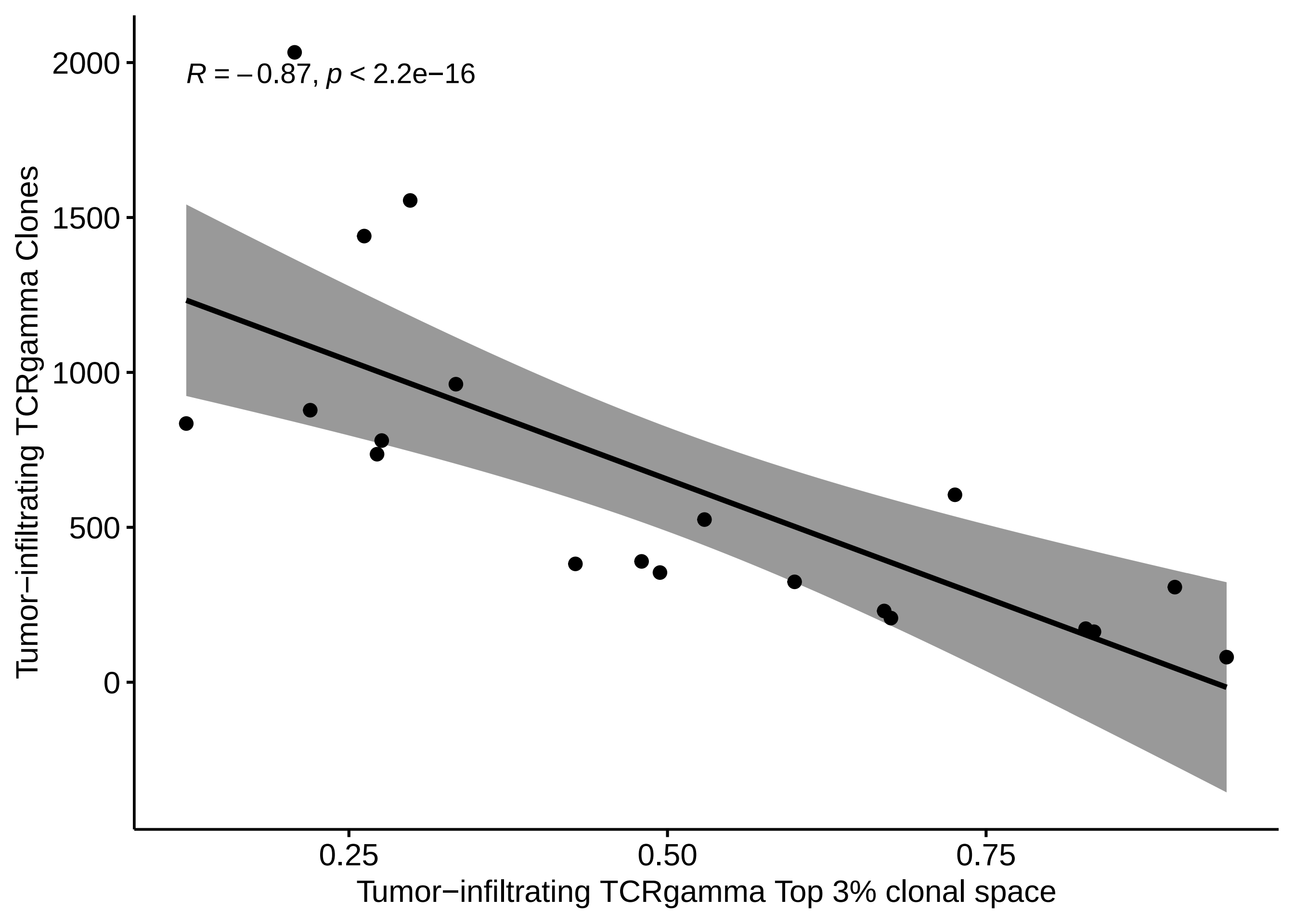
R = −0.87, p < 2.2e−16
2000
1500
1000
500
0
0.25
0.50
0.75
Tumor−infiltrating TCRgamma Clones
Tumor−infiltrating TCRgamma Top 3% clonal space

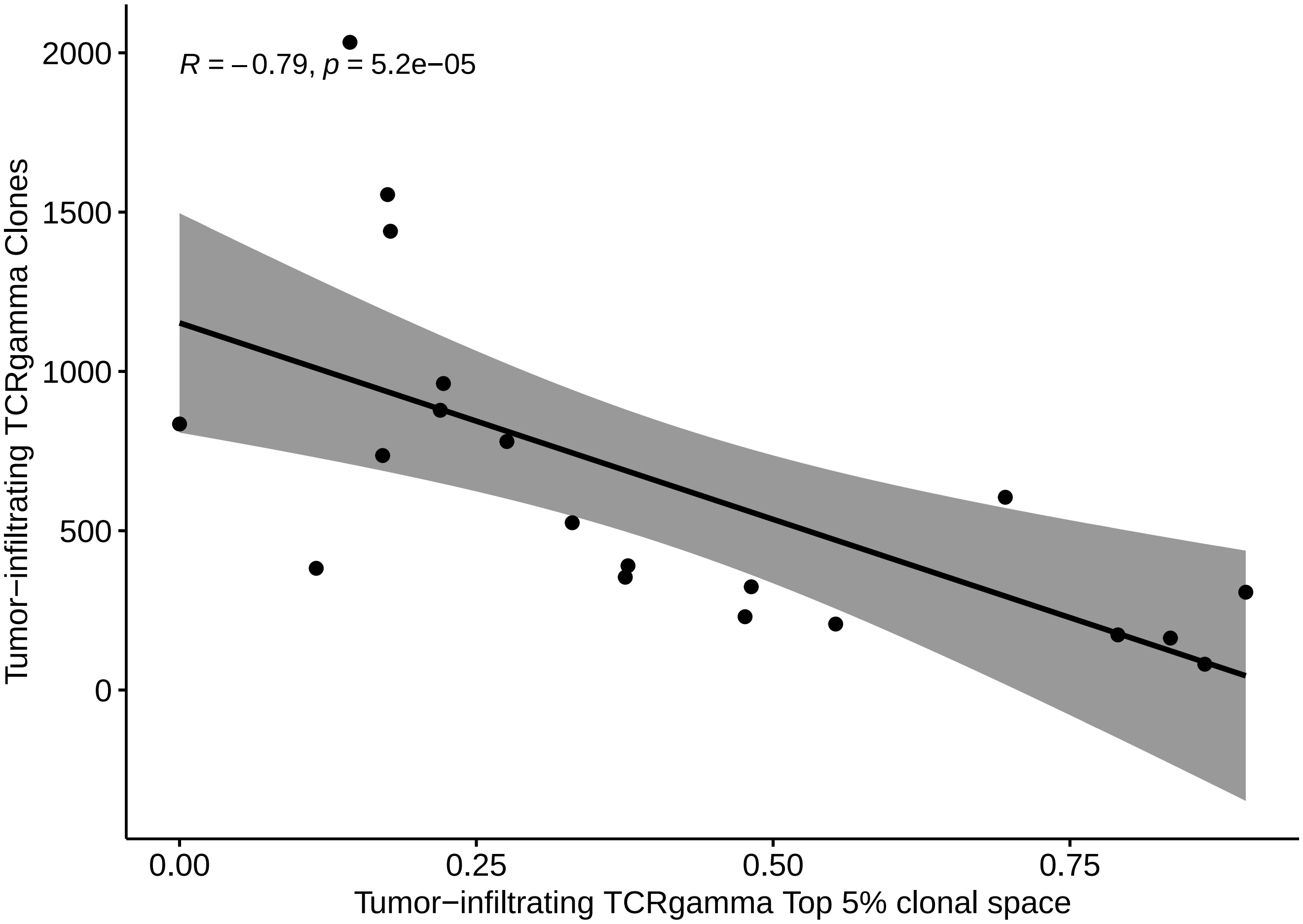

R = −0.79, p = 5.2e−05
2000
1500
1000
500
0
Tumor−infiltrating TCRgamma Clones
0.00
0.25
0.50
0.75
Tumor−infiltrating TCRgamma Top 5% clonal space

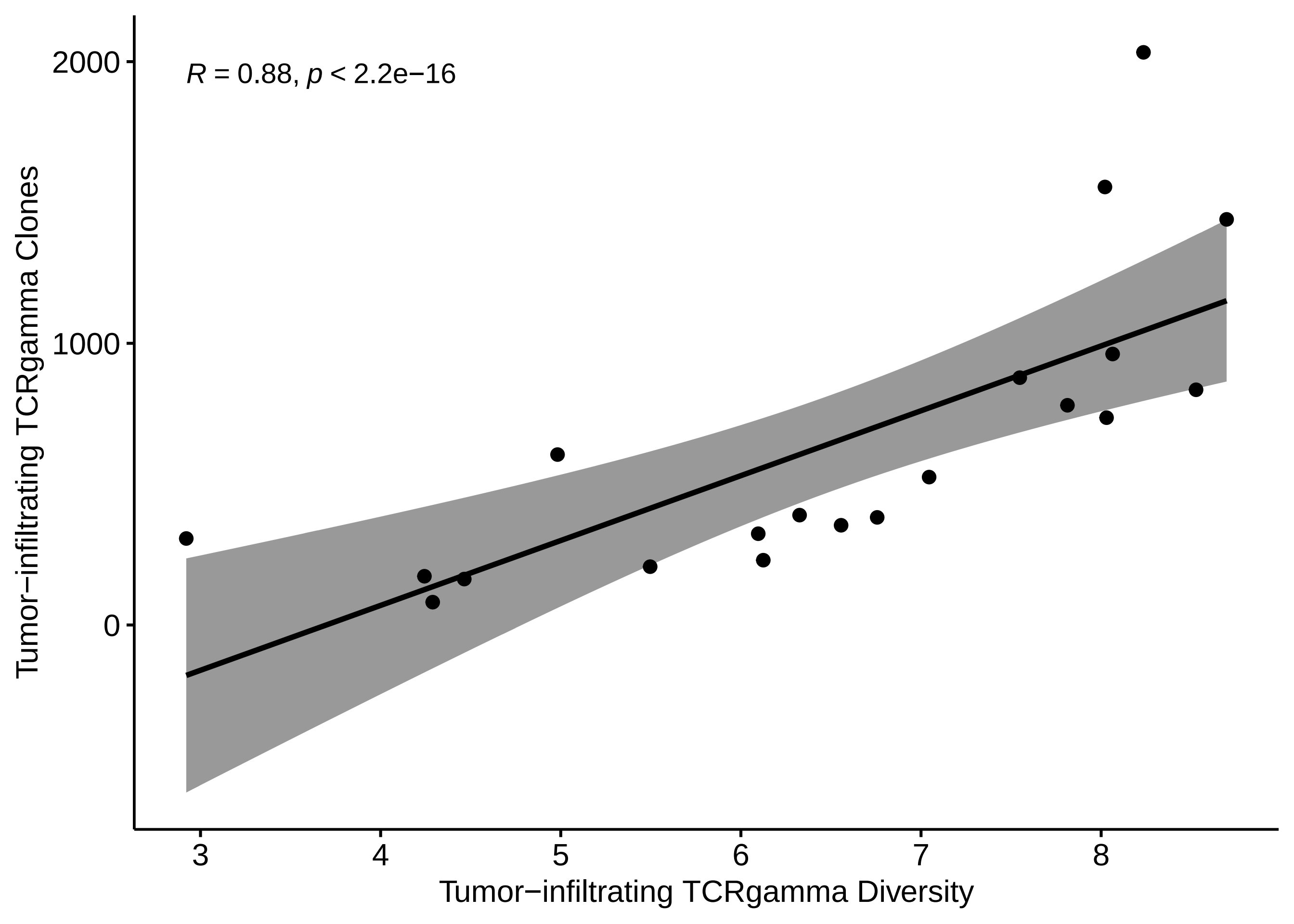

R = 0.88, p < 2.2e−16
Tumor−infiltrating TCRgamma Clones
2000
1000
0
3
4
5
6
7
8
Tumor−infiltrating TCRgamma Diversity

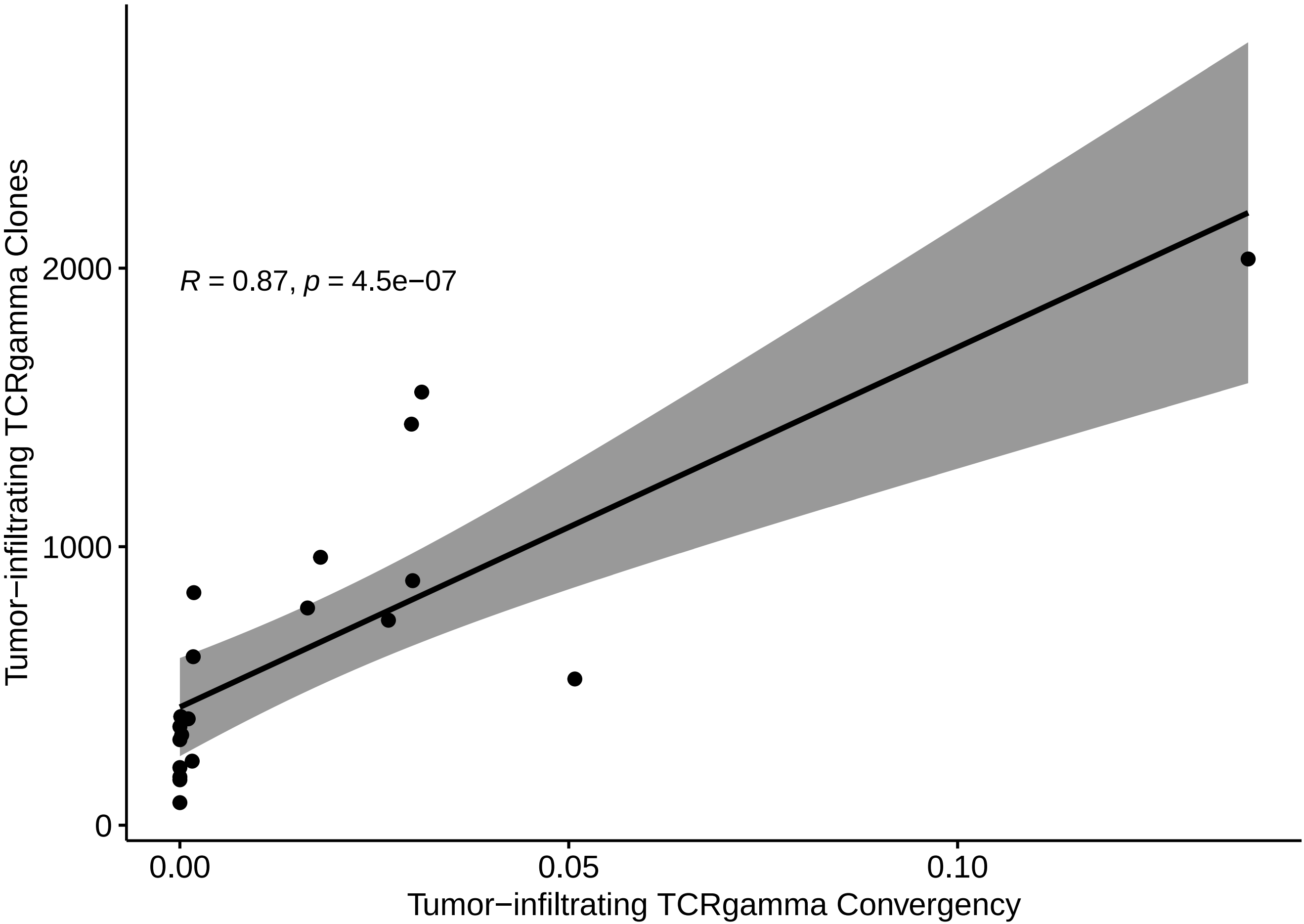
Tumor−infiltrating TCRgamma Clones
2000
1000
0
R = 0.87, p = 4.5e−07
0.00
0.05
0.10
Tumor−infiltrating TCRgamma Convergency

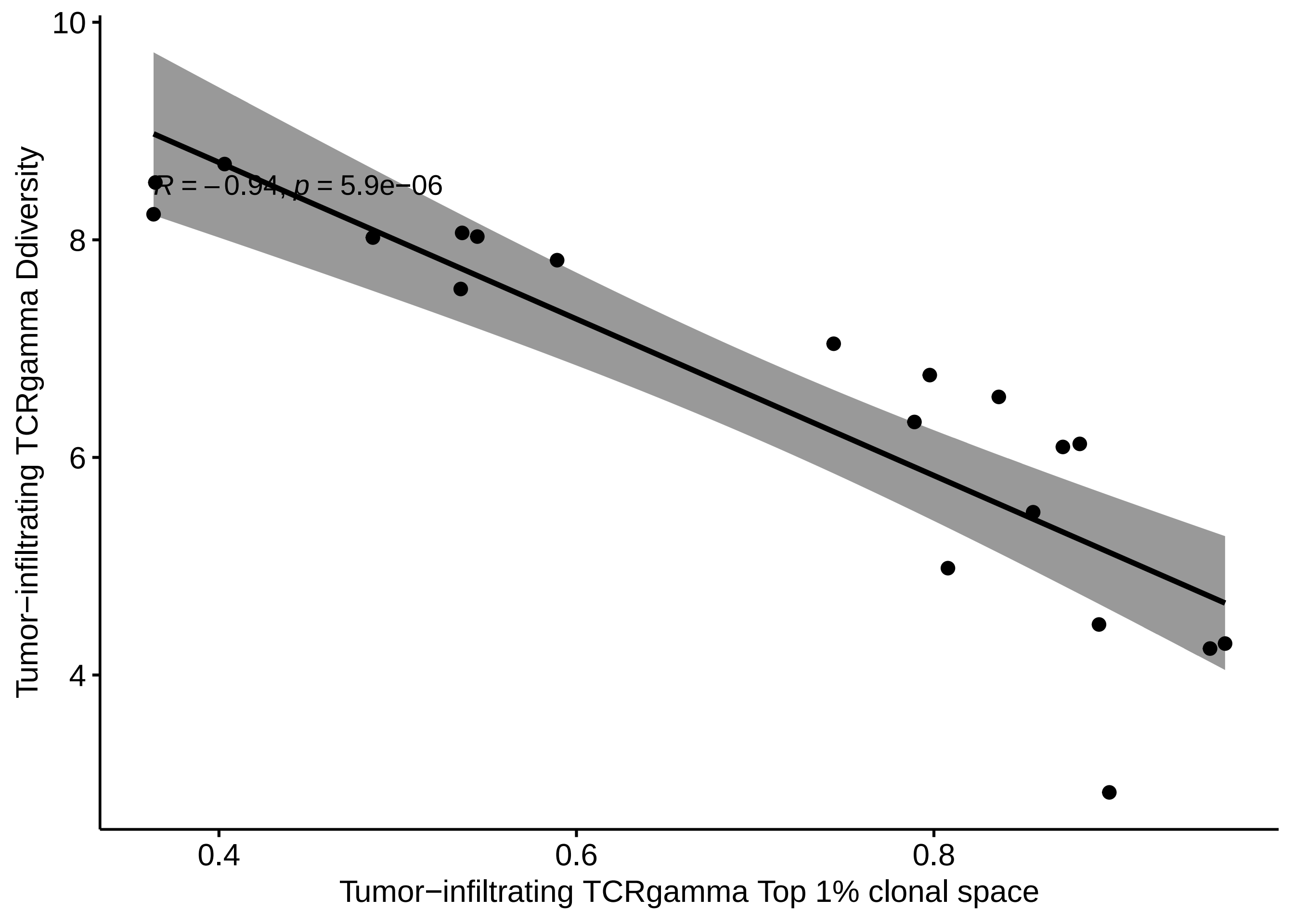
R = −0.94, p = 5.9e−06
Tumor−infiltrating TCRgamma Ddiversity
10
8
6
4
0.4
0.6
0.8
Tumor−infiltrating TCRgamma Top 1% clonal space

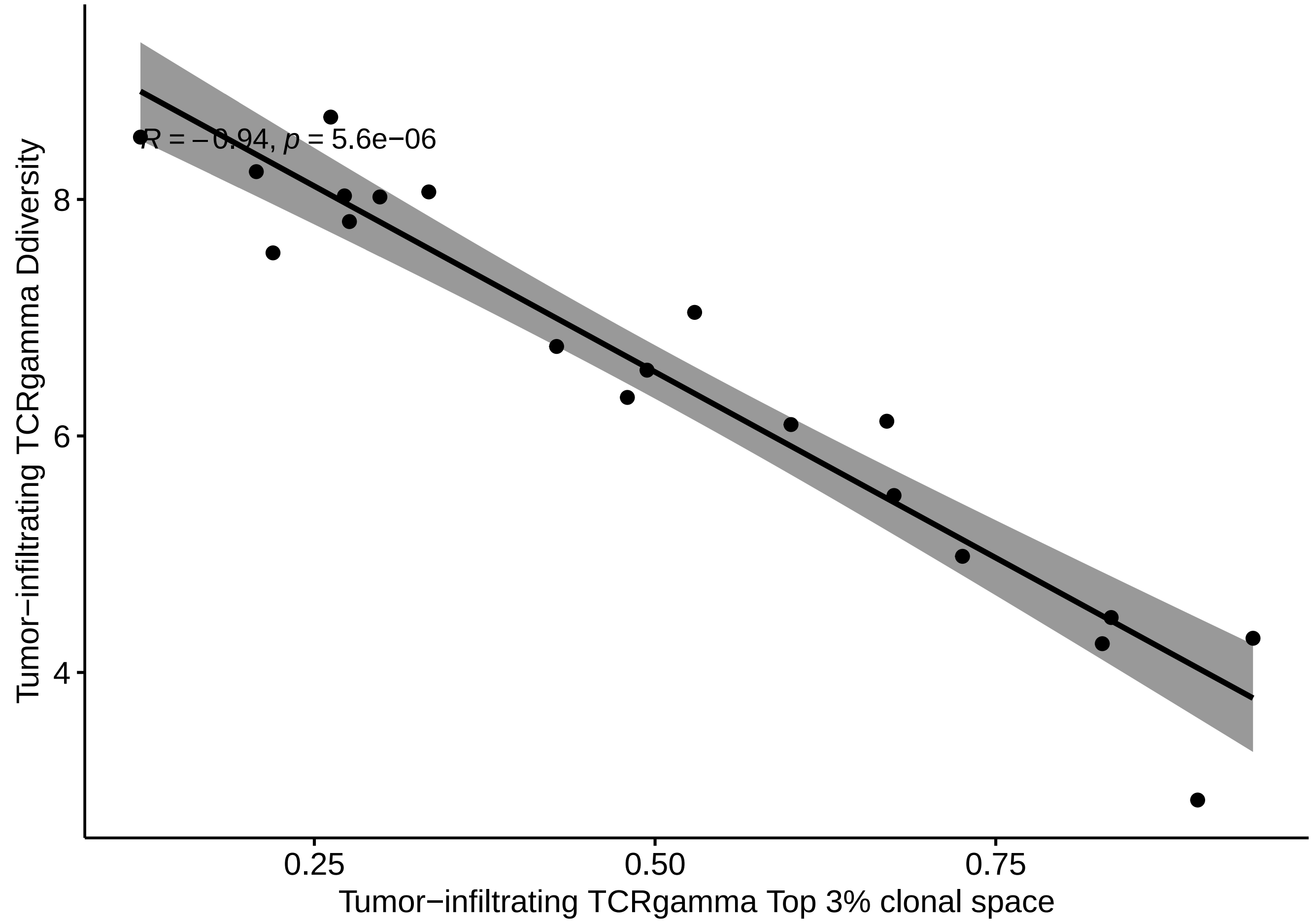
R = −0.94, p = 5.6e−06
Tumor−infiltrating TCRgamma Ddiversity
8
6
4
0.25
0.50
0.75
Tumor−infiltrating TCRgamma Top 3% clonal space

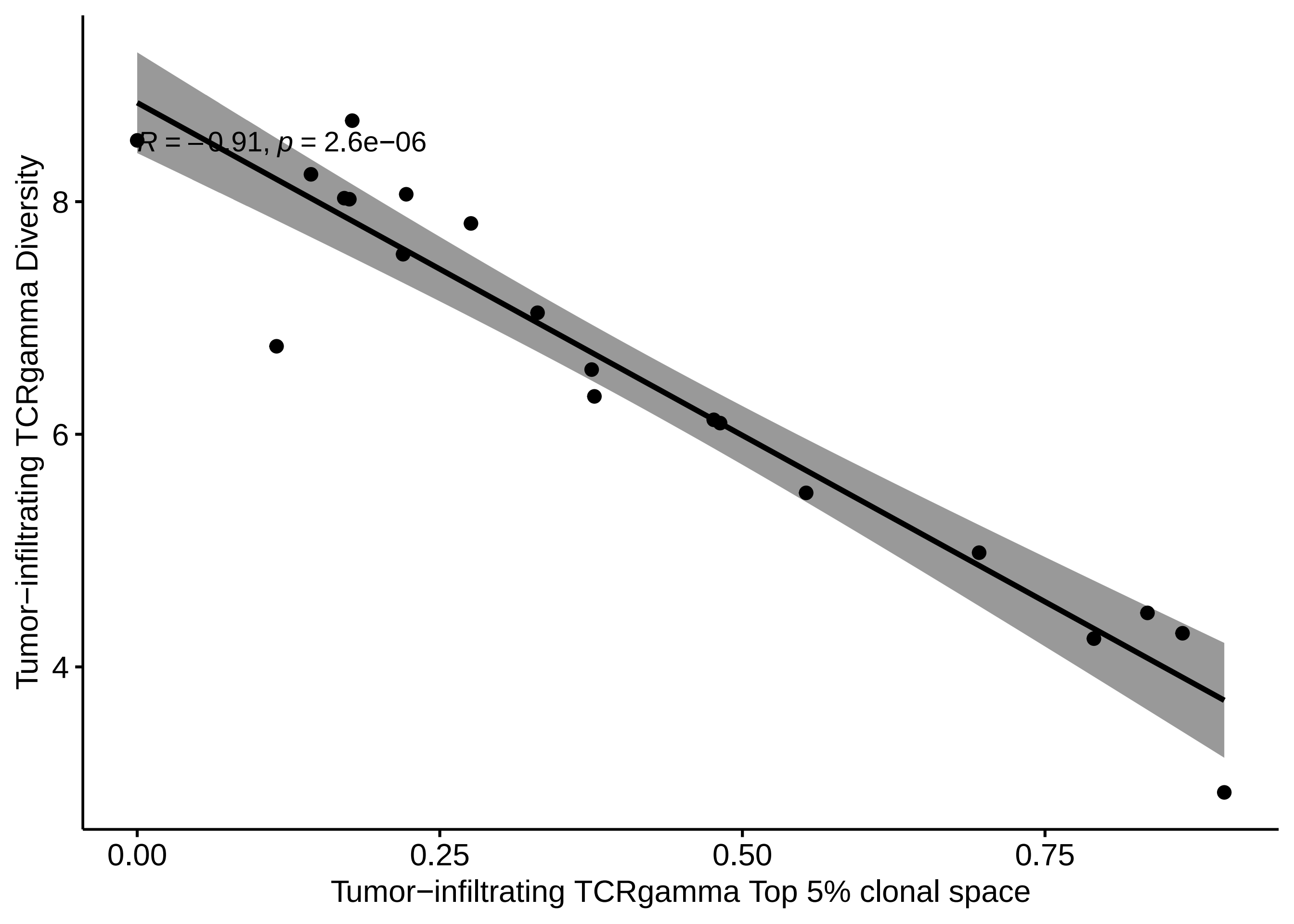

R = −0.91, p = 2.6e−06
Tumor−infiltrating TCRgamma Diversity
8
6
4
0.00
0.25
0.50
0.75
Tumor−infiltrating TCRgamma Top 5% clonal space

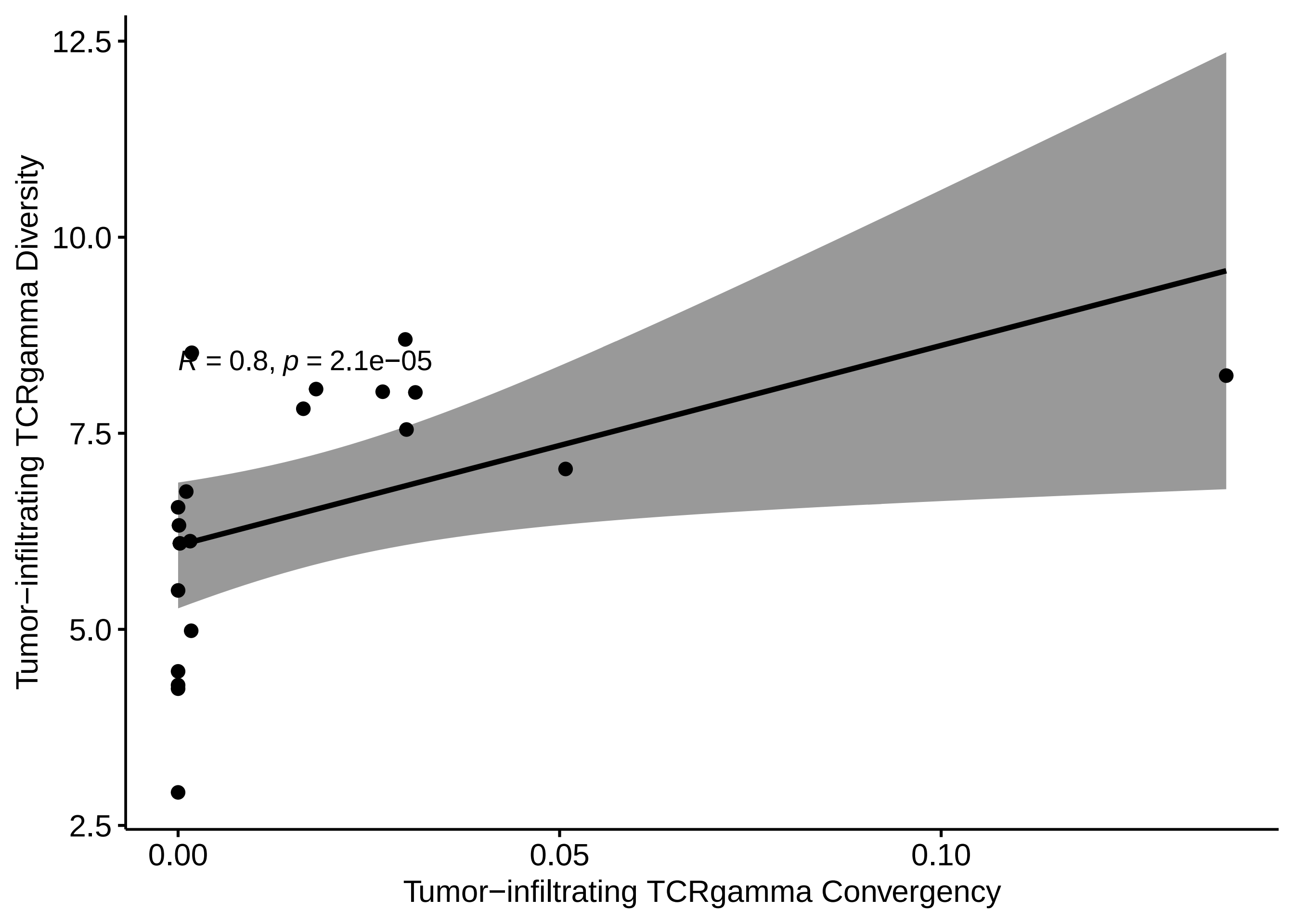
R = 0.8, p = 2.1e−05
12.5
10.0
7.5
5.0
2.5
0.00
0.05
0.10
Tumor−infiltrating TCRgamma Diversity
Tumor−infiltrating TCRgamma Convergency

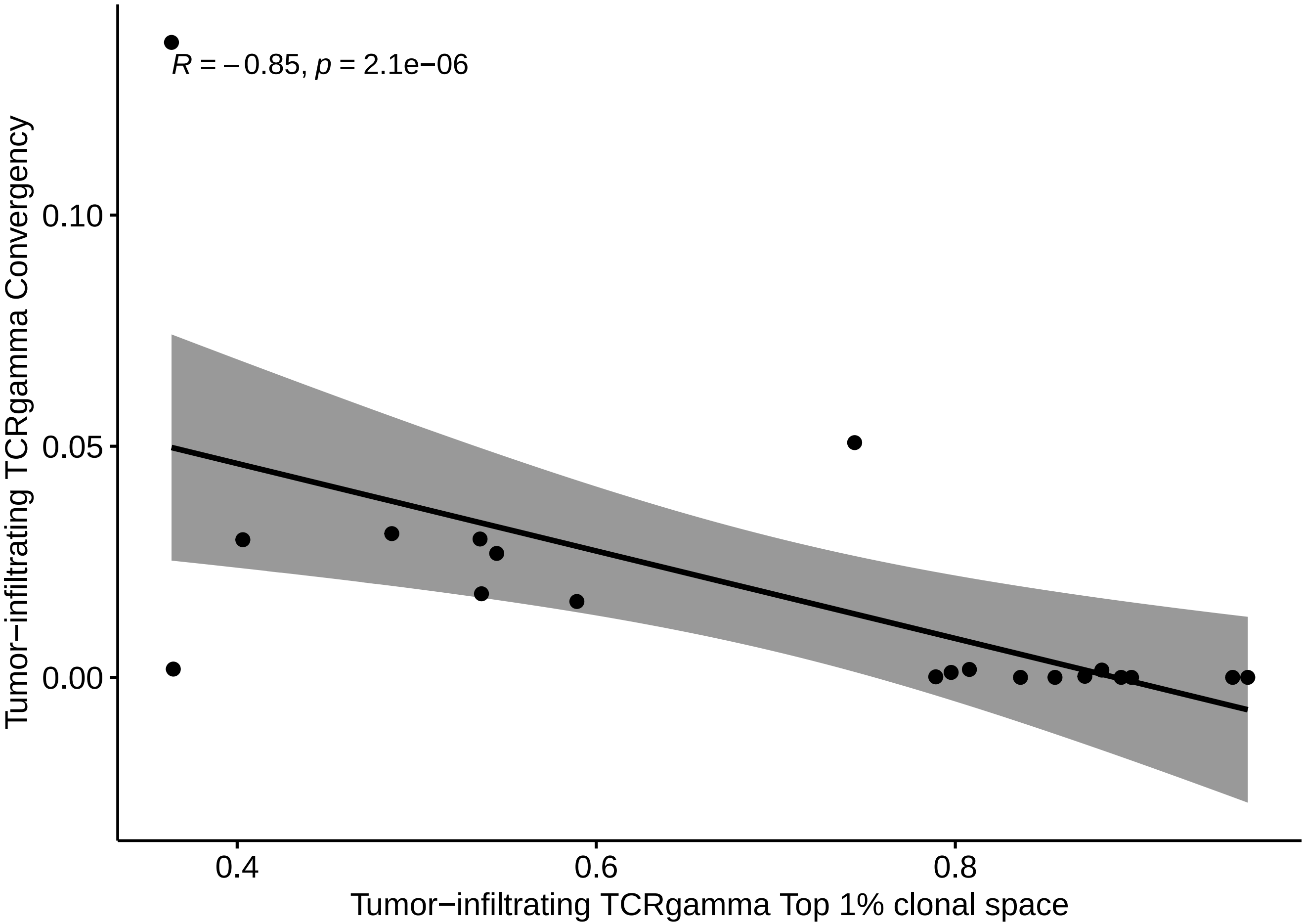
R = −0.85, p = 2.1e−06
0.10
0.05
0.00
0.4
0.6
0.8
Tumor−infiltrating TCRgamma Convergency
Tumor−infiltrating TCRgamma Top 1% clonal space

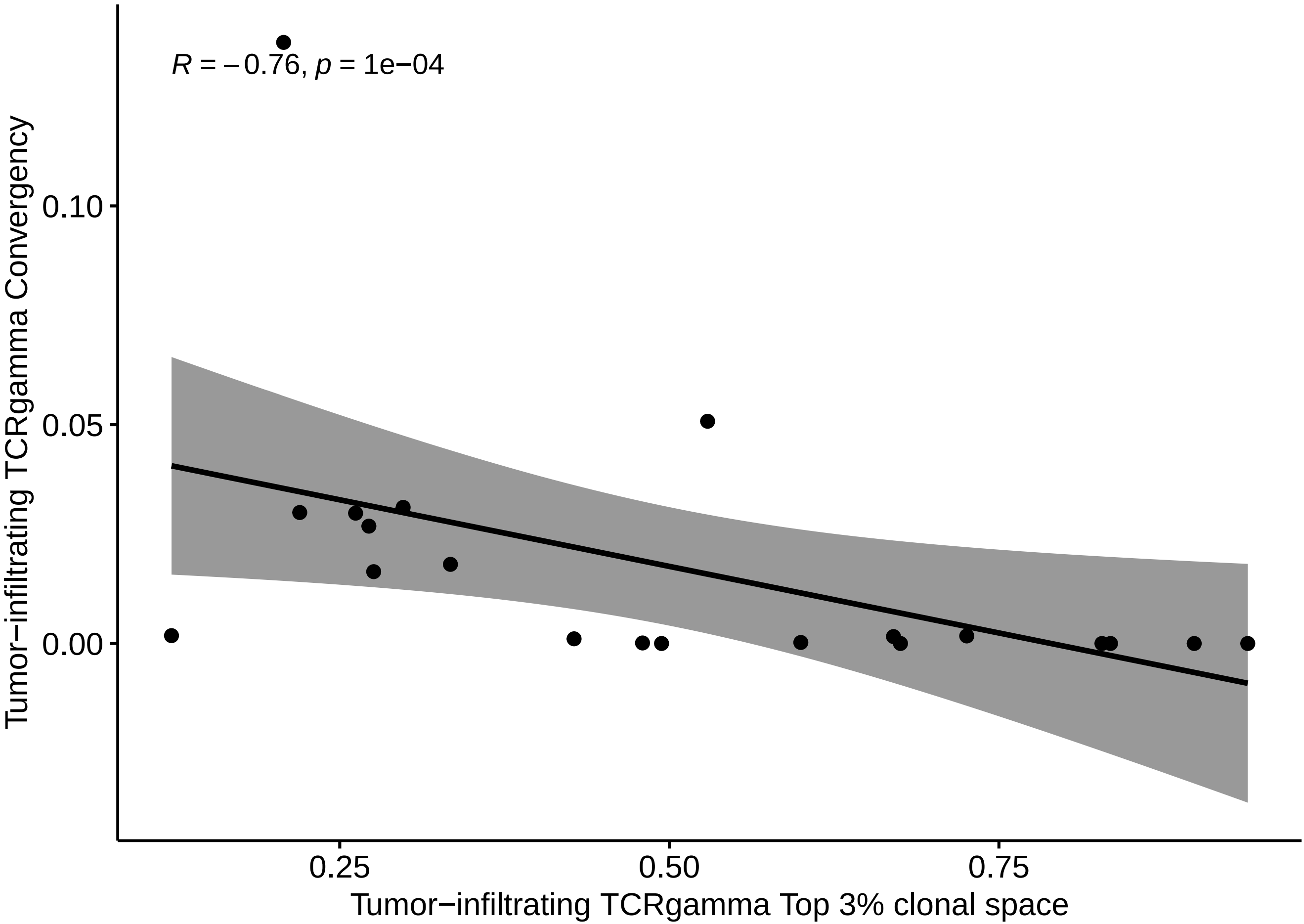

R = −0.76, p = 1e−04
Tumor−infiltrating TCRgamma Convergency
0.10
0.05
0.00
0.25
0.50
0.75
Tumor−infiltrating TCRgamma Top 3% clonal space

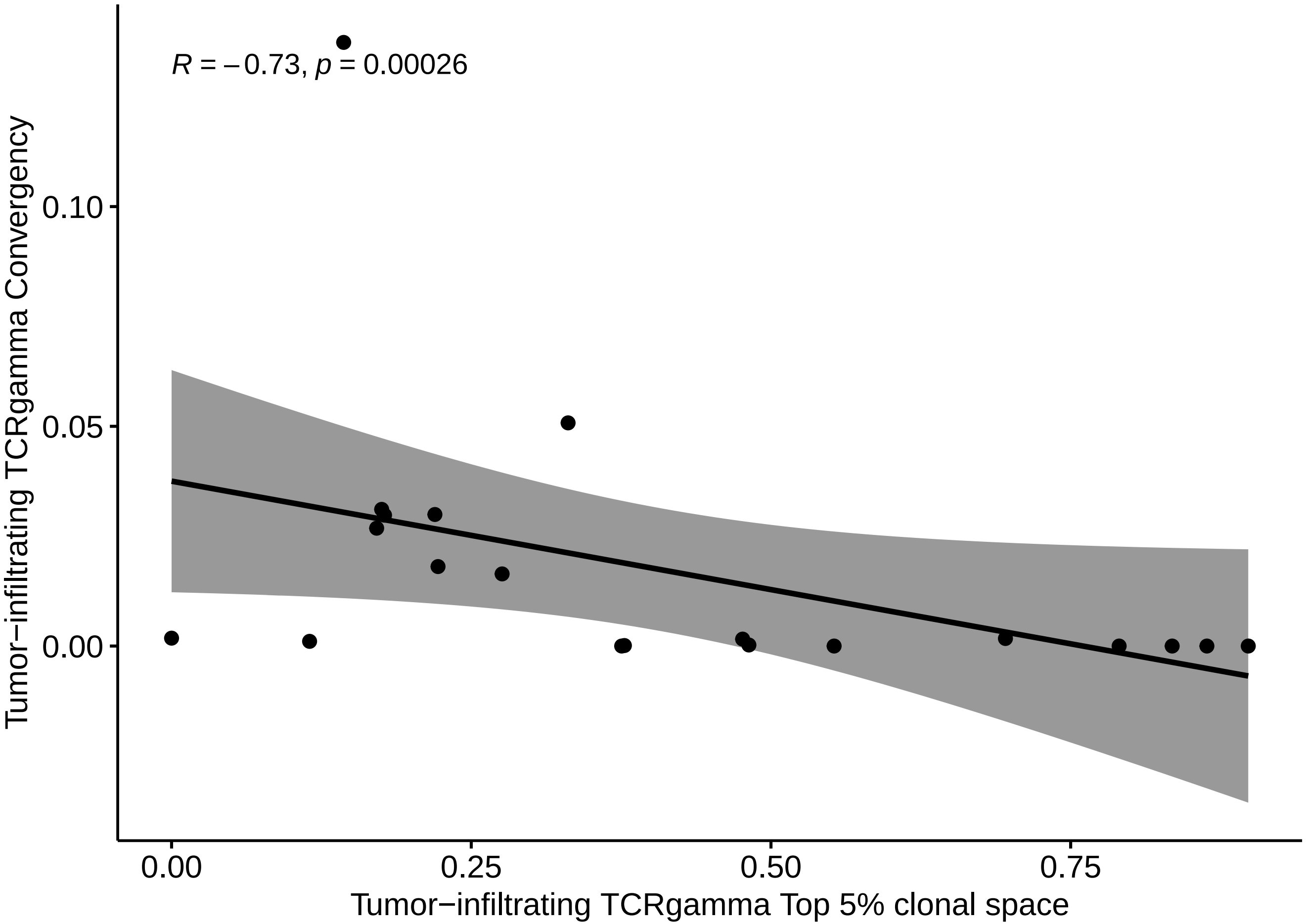

R = −0.73, p = 0.00026
0.10
0.05
0.00
Tumor−infiltrating TCRgamma Convergency
0.00
0.25
0.50
0.75
Tumor−infiltrating TCRgamma Top 5% clonal space
